# Partial ORF1ab Gene Target Failure with Omicron BA.2.12.1

**DOI:** 10.1101/2022.04.25.22274187

**Authors:** Kyle G. Rodino, David R. Peaper, Brendan J. Kelly, Frederic Bushman, Andrew Marques, Hriju Adhikari, Zheng Jin Tu, Rebecca Marrero Rolon, Lars F. Westblade, Daniel A. Green, Gregory J. Berry, Fann Wu, Medini K. Annavajhala, Anne-Catrin Uhlemann, Bijal A. Parikh, Tracy McMillen, Krupa Jani, N. Esther Babady, Anne M. Hahn, Robert T. Koch, Nathan D. Grubaugh, Yale SARS-CoV-2 Genomic Surveillance Initiative, Daniel D. Rhoads

## Abstract

Mutations in the viral genome of SARS-CoV-2 can impact the performance of molecular diagnostic assays. In some cases, such as S gene target failure, the impact can serve as a unique indicator of a particular SARS-CoV-2 variant and provide a method for rapid detection. Here we describe partial ORF1ab gene target failure (pOGTF) on the cobas^®^ SARS-CoV-2 assays, defined by a ≥2 thermocycles delay in detection of the ORF1ab gene compared to the E gene. We demonstrate that pOGTF is 97% sensitive and 99% specific for SARS-CoV-2 lineage BA.2.12.1, an emerging variant in the United States with spike L452Q and S704L mutations that may impact transmission, infectivity, and/or immune evasion. Increasing rates of pOGTF closely mirrored rates of BA.2.12.1 sequences uploaded to public databases, and, importantly increasing local rates of pOGTF also mirrored increasing overall test positivity. Use of pOGTF as a proxy for BA.2.12.1 provides faster tracking of the variant than whole-genome sequencing and can benefit laboratories without sequencing capabilities.

## Introduction

Mutations in the primer/probe binding sites of the SARS-CoV-2 genome can impact oligonucleotide binding and molecular test performance. Throughout the pandemic a number of such mutations have been described, resulting in partial or complete PCR target failure (1). Mutations that impair diagnostic detection is a criterion that the United States (US) Center for Disease Control and Prevention (CDC) considers when classifying a novel SARS-CoV-2 lineage as a Variant of Concern (2). Most nucleic acid amplification tests (NAAT) used for *in vitro* diagnostic (IVD) clinical testing and which have received emergency use authorization (EUA) from the US Food and Drug Administration (FDA) employ multi-target assay design. This limits the diagnostic impact of a SARS-CoV-2 mutation that causes single-target failure, as the additional primer/probe target sequences remain unaltered and adequately detect the presence of viral nucleic acid.

Viral mutations can negatively impact assay performance, but these phenomena have proven useful in certain situations. Whole-genome sequencing (WGS) of SARS-CoV-2 remains inadequately implemented or unavailable in many medical centers across the US due to logistical, cost, and time constraints. Moreover, full genomic characterization of circulating variants often takes multiple weeks from sample collection to data reporting to public health agencies. Specific mutations that yield unique NAAT performance characteristics can provide broader and more rapid assessment of circulating lineages than WGS. For example, the well characterized six base pair deletion in the spike gene resulting in the absence of amino acids H69/V70 (69-70del) results in complete loss of detection of the spike gene target on the Thermo Fisher Scientific TaqPath™ COVID-19 assay (Thermo Fisher Scientific, Waltham, MA), commonly termed <u>S</u>-<u>G</u>ene <u>T</u>arget <u>F</u>ailure (SGTF). SGTF was a hallmark of the Alpha (B.1.1.7) variant allowing for sequence-free estimation of Alpha’s emergence and prevalence in early 2021 (3). Utility in tracking SGTF was observed in late 2021 with the introduction of the Omicron (B.1.1.529.1 [BA.1]) variant, which contained the same characteristic 69-70del. Rapid estimation of Omicron emergence was possible as the preceding dominant variant, Delta, did not harbor the 69-70del and was therefore <u>S</u>-<u>G</u>ene <u>T</u>arget <u>P</u>ositive (SGTP) (4). Finally, in early 2022, SGTF tracking proved useful in monitoring the transition from BA.1 Omicron to BA.2 Omicron (B.1.1.529.2), as the latter lacked the 69-70del and was SGTP, which served as a reliable proxy for identifying BA.2 (5).

While the impact of the 69-70del was dramatic, resulting in complete SGTF, other genomic mutations may result in subtler changes including abnormal variance between cycle threshold (C_*T*_) values of multi-target SASR-CoV-2 assays (6-10). Here, we describe <u>p</u>artial <u>O</u>RF1ab <u>G</u>ene <u>T</u>arget <u>F</u>ailure (pOGTF) as detected by abnormal <u>d</u>elta of the C_*T*_ values from the <u>E</u> and <u>O</u>RF1ab (dEO) targets of the cobas^®^ SARS-CoV-2 assay and cobas^®^ SARS-CoV-2 & Influenza A/B assay (Roche Molecular Systems, Inc.; Branchburg, NJ) linked to BA.2.12.1, which has rapidly emerged in the US in March-April 2022 and has become the dominant SARS-CoV-2 lineage in central New York state (11).

## Materials and Methods

### Study Population

Samples used in this study included upper respiratory specimens (e.g., nasopharyngeal, anterior nares, nasal swabs, and saliva) collected in a transport medium validated by the individual testing laboratory, obtained from patients undergoing standard of care testing for COVID-19 between March 6, 2022 and April 16, 2022, a period of time corresponding to MMWR epidemiologic weeks 10 to 15 (12). Samples included in the dEO analysis were tested for the presence of SARS-CoV-2 on cobas^®^ SARS-CoV-2 assay or cobas^®^ SARS-CoV-2 & Influenza A/B assays at the originating laboratory. Samples sent for SARS-CoV-2 WGS may have been tested on a variety of other FDA EUA-approved SARS-CoV-2 diagnostic platforms, and not a cobas^®^ SARS-CoV-2 assay, depending on workflows and testing protocols of the originating laboratory. Laboratories contributing to this study and appropriate IRB determination include the Cleveland Clinic Foundation (CCF) Clinical Microbiology Laboratory (IRB #18-318), the Clinical Microbiology Laboratory of Columbia University Irving Medical Center (CUIMC) (IRB #AAT0123), the Clinical Microbiology and SARS-CoV-2 Molecular Testing Laboratories at the Hospital of the University of Pennsylvania (HUP) (IRB #848605), the Clinical Microbiology Service at Memorial Sloan Kettering (MSK) (IRB #18-491), the Clinical Microbiology Laboratory of Weill Cornell Medical Center (WCMC) (IRB #20-03021671), the Barnes-Jewish Hospital Molecular Infectious Disease Laboratory and Washington University in St. Louis (WUSTL) (IRB #20211131), and Clinical Virology Laboratory at Yale-New Haven Hospital (YNHH) and Yale School of Public Health (IRB #2000031374).

### cobas^®^ SARS-CoV-2 assays

The cobas^®^ SARS-CoV-2 and cobas^®^ SARS-CoV-2 & Influenza A/B assays are available on the Roche cobas^®^ 6800 and 8800 analyzers (Roche Molecular Systems, Inc.). Both assays perform qualitative detection of SARS-CoV-2 using two genome targets; the ORF1ab gene that is specific to SARS-CoV-2 and the pan-Sarbecovirus envelope (E) gene. Details on the oligonucleotide sequences and the specific genomic regions targeted by the NAAT are not publicly available. MS2 bacteriophage is used as an internal RNA processing control. RNA extraction, reverse transcription, target amplification, and result analysis all occur on the instrument. Positive results are determined by amplification curves that cross a predetermined threshold thus generating C_*T*_ values for these loci. These C_*T*_ values are available to the laboratory, but are not included in patient reports by any of the performing laboratories. Only qualitative interpretations (SARS-CoV-2 detected, not detected, or presumptive positive) of the results are reported, which is congruent with the EUA instructions for use. Samples do not have to be positive for both gene targets to be called positive by the assay. However, only results positive for both targets were used in this study to allow for C_*T*_ value comparison. C_*T*_ values used in this study as well as dates of sample collection and/or testing were either directly obtained from instruments or extracted from the laboratory information system depending on the testing laboratory.

### Identification of samples as dEO outliers

Initial recognition of dEO was determined by manual review of C_*T*_ values, noticing ≥1 or ≥2 cycle difference, between the ORF1ab and E C_*T*_ values, with ORF1ab C_*T*_ value > E gene C_*T*_ value, depending on the institution. At HUP we modeled the expected difference in C_*T*_ values between two parallel targets as a function of the minimum observed C_*T*_ value in the pair, in order to accommodate the observation that the expected difference in C_*T*_ values across targets increases at higher C_*T*_ values. The expected variation in the C_*T*_ value difference was likewise allowed to vary with the minimum observed C_*T*_ value in the pair. Bayesian mixed-effect linear regression models, incorporating random effects (slope and intercept) for sequencing platform (i.e., amplicon-target pair), were fit using Stan Hamiltonian Monte Carlo via the brms package in R (13, 14). Model parameters were established using C_*T*_ value data collected across seven platforms from March to June 2021, and the parameterized model was used to monitor for dEO (and other target pair) outliers, with outliers identified as differences beyond the 99% posterior credible interval of the expected C_*T*_ value difference for the minimum C_*T*_ value observed in the pair. Model code and sample reproducible report are available at: https://github.com/bjklab/SARS-CoV-2_Ct_report.

### SARS-CoV-2 Whole-Genome Sequencing

At CCF, NGS sequencing libraries were prepared with Illumina COVIDSeq Test kit according to the manufacture’s recommendation. These libraries were sequenced paired end with read length 151 bases using a NextSeq550 instrument (Illumina, San Diego, CA). Data analysis was performed with an in-house developed bioinformatics pipeline. Sequence reads were mapped to reference genome Wuhan-Hu-1 (NC_045512.2) using BWA (version 0.7.15), variant calling was performed with both Freebayes (version 1.3.4) and LoFreq (version 2.1.5). Average coverage is x6567 and minimum coverage for mutation is x10. Samples below this minimum coverage at the S gene codon 452 were considered unreliable, and these genotypes were not considered in this study. Manual review of mutations was performed with Integrative Genomics Viewer (IGV) to remove any artifacts. Variant classification was performed with the Pangolin program (https://pangolin.cog-uk.io/ version v4.0.5, lineages version 2022-04-09). Derived genomes with related information were deposited into GISAID database. Samples from (CUIMC) and (WCMC) campuses were sequenced using the Oxford Nanopore (Oxford Nanopore Technologies, Oxford, UK) Midnight protocol targeting 1200bp tiled amplicons across the length of the genome, as previously described (15). Samples were sequenced on an Oxford Nanopore GridION using R9.4.1 flow cells, with negative controls included on each run. Variant calling and consensus genome generation was performed using the Oxford Nextflow ARTIC pipeline. Viral lineage classification, identification of mutations, and phylogenetic analyses were performed using Pangolin v4.0.5 and Nextclade v1.11.0. Genomic data from CUIMC is routinely uploaded to GISAID and to GenBank (under NCBI BioProject PRJNA751551). At Yale, sequencing was performed as previously described, but Pangolin v4.0.5 was used (16). All other SARS-CoV-2 WGS and lineage assignment was performed as previously described (17-19).

### Data Analysis

All charts and analyses were generated and performed with GraphPad Prism v8.2.1. Non-linear regression of BA.2.12.1 and non-BA.2.12.1 was performed and best fit straight lines using the least squares method and *y*-intercepts were compared for differences. Receiver operating characteristic (ROC) curve to determine the performance characteristics of different dEO cut-off values was generated by classifying samples with lineage information available as either “BA.2.12.1” or “Other”. Positive and negative predictive values of pOGTF for the detection of BA.2.12.1 were calculated using samples with lineage information classified as either “BA.2.12.1” or “Other” where pOGTF was considered a dEO of ≥2 with an E-gene C_*T*_ value ≤30.

### Data availability

All SARS-CoV-2 viral genomes have been deposited in GISAID and/or NCBI Genbank with accession numbers or viral names listed in Table S1.

## Results

Difference in C*T* values for the E gene target compared to the ORF1ab gene target (dEO) from March 6, 2022 to April 16, 2022 were plotted (Figure 1, Table S1). Using an E gene C_*T*_ value cutoff of ≤30 thermocycles to avoid non-specific variation between values near the limit of detection of the assay, an outlier group with abnormal difference between the two C_*T*_ values was identified. These data indicate delayed detection of the ORF1ab target as compared to the E gene target. In total, 428 samples, run on either the classic or combo SARS-CoV-2 assay, with pOGTF were detected during the study period.

**Figure 1.**
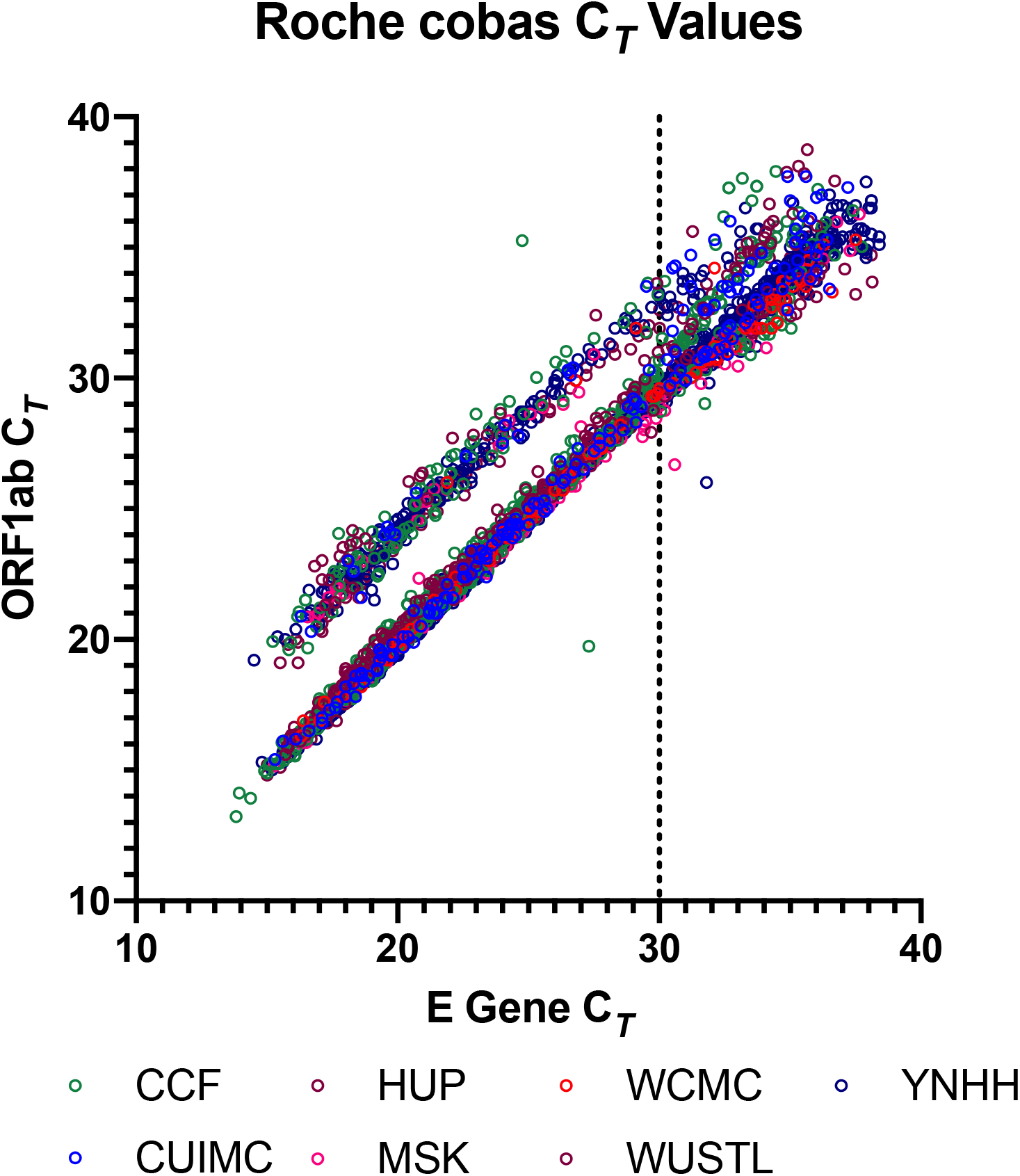
Plot of ORF1ab and E-gene C_*T*_ values generated using the cobas^®^ SARS-CoV-2 or cobas^®^ SARS-CoV-2 & Influenza A/B assays. ORF1ab and E loci C_*T*_ values from the indicated institutions for all positive results from March 6, 2022 to April 16, 2022. Vertical line indicates an E-gene C_*T*_ value of 30 that was used as a cut-off for subsequent analyses. Samples with only a single gene detected were not included and are not shown. A total of 3,471 unique samples are included. Abbreviations: Cleveland Clinic Foundation (CCF), Columbia University Irving Medical Center (CUIMC), Hospital of the University of Pennsylvania (HUP), Memorial Sloan Kettering (MSK), Weill-Cornell Medical Center (WCMC), Washington University in St. Louis (WUSTL), and Yale-New Haven Hospital (YNHH).

Following institution-specific workflows for SARS-CoV-2 surveillance sequencing, a subset of samples from the study period underwent WGS and lineage determination, including 71 samples showing pOGTF (Table S1). Overwhelmingly, pOGTF samples were determined to be BA.2.12.1 (70/71), with the first WGS-confirmed detection of BA.2.12.1 among study participants occurring in the first week of March in the YNHH cohort and remaining study sites also confirming BA.2.12.1 from pOGTF specimens in late-March. The emergence of samples with pOGTF coincided with increases in BA.2.12.1, and preliminary evidence suggested that pOGTF may be a marker for BA.2.12.1. To explore this ORF1ab and E gene C_*T*_ values were plotted for samples confirmed as BA.2.12.1 or another lineage by sequencing. BA.2.12.1 samples also clustered as dEO outliers (Figure 2). Best-fit straight lines demonstrated these to be different populations with significantly different y-intercepts (p < 0.0001). Further analysis of sequencing data demonstrated that BA.2.12.1, a sub-lineage of BA.2, contains five characteristic mutations of interest to this study, compared to the parent strain. These include two missense mutations altering amino acids in the spike gene, g.22917 T>G (S:L452Q) and g.23673 C>T (S:S704L), and three synonymous mutations g.11674 C>T (ORF1ab), g.15009 T>C (ORF1ab), and g.21721 C>T (S). Likely, one of the two ORF1ab synonymous mutations causes reduced efficiency of amplification and/or detection of the target in the cobas^®^ SARS-CoV-2 and cobas^®^ SARS-CoV-2 & Influenza A/B assays.

**Figure 2.**
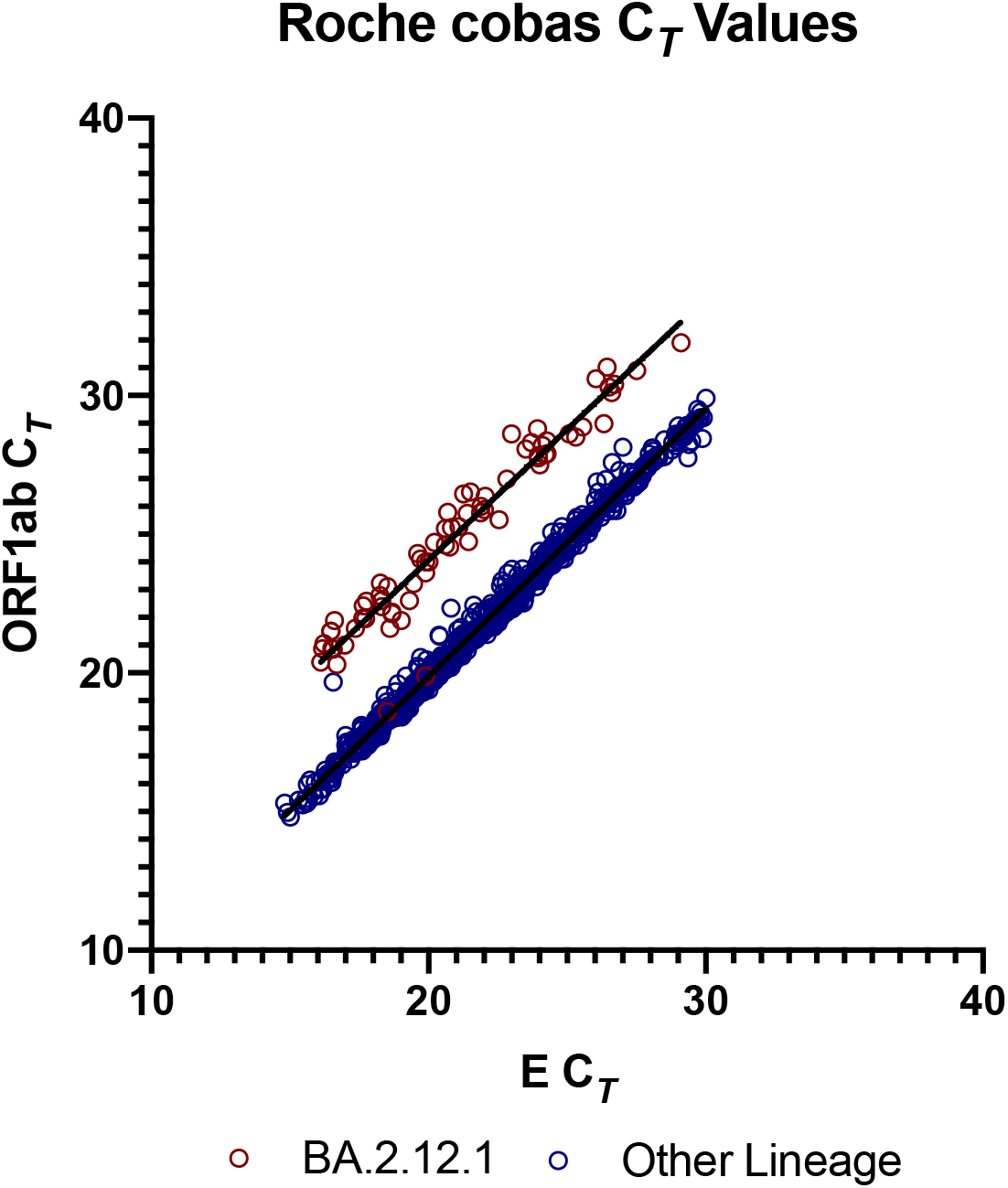
Plot of ORF1ab and E-gene C_*T*_ values generated using the cobas^®^ SARS-CoV-2 or cobas^®^ SARS-CoV-2 & Influenza A/B assays for samples with lineage results. ORF1ab and E loci C_*T*_ values from all institutions for samples with lineage results from March 6, 2022 to April 16, 2022 are shown. All lineages other than BA.2.12.1 were consolidated into a single category. A total of 72 BA.2.12.1 and 766 other lineage samples were included. Linear regression was performed, and best fit curves for BA.2.12.1 and all other lineages were had significantly different *y*-intercepts (p < 0.0001). Only samples with E-gene C_*T*_ values ≤30 were used for regression analysis.

We used the subset of samples with lineage information to generate an ROC curve to determine the performance characteristics of different cut-off values for dEO (Figure 3). At a dEO cut-off of 2.1, the sensitivity of was 97.2% while the specificity was 99.9% generated during the ROC analysis. After review of the primary data, there was no difference in performance using a cutoff of 2.0 vs. 2.1, thus dEO ≥ 2.0 was selected for simplicity of use in identifying samples with pOGTF (Table 1). When this criterion was applied to our population of samples with lineage information with E gene C_*T*_ values ≤30 the positive and negative predictive values were 98.6% and 99.7%, respectively. Using the thresholds if E gene C_*T*_ values ≤30 and dEO ≥2 to define pOGTF, we reanalyzed our initial cobas SARS-CoV-2 results, finding that the pOGTF group showed a mean dEO of 4.02 (95% CI 3.95 to 4.09) cycles as compared to -0.13 (95% CI - 0.14 to -0.11) cycles in the non-pOGTF group (Table 2).

**Table 1.**
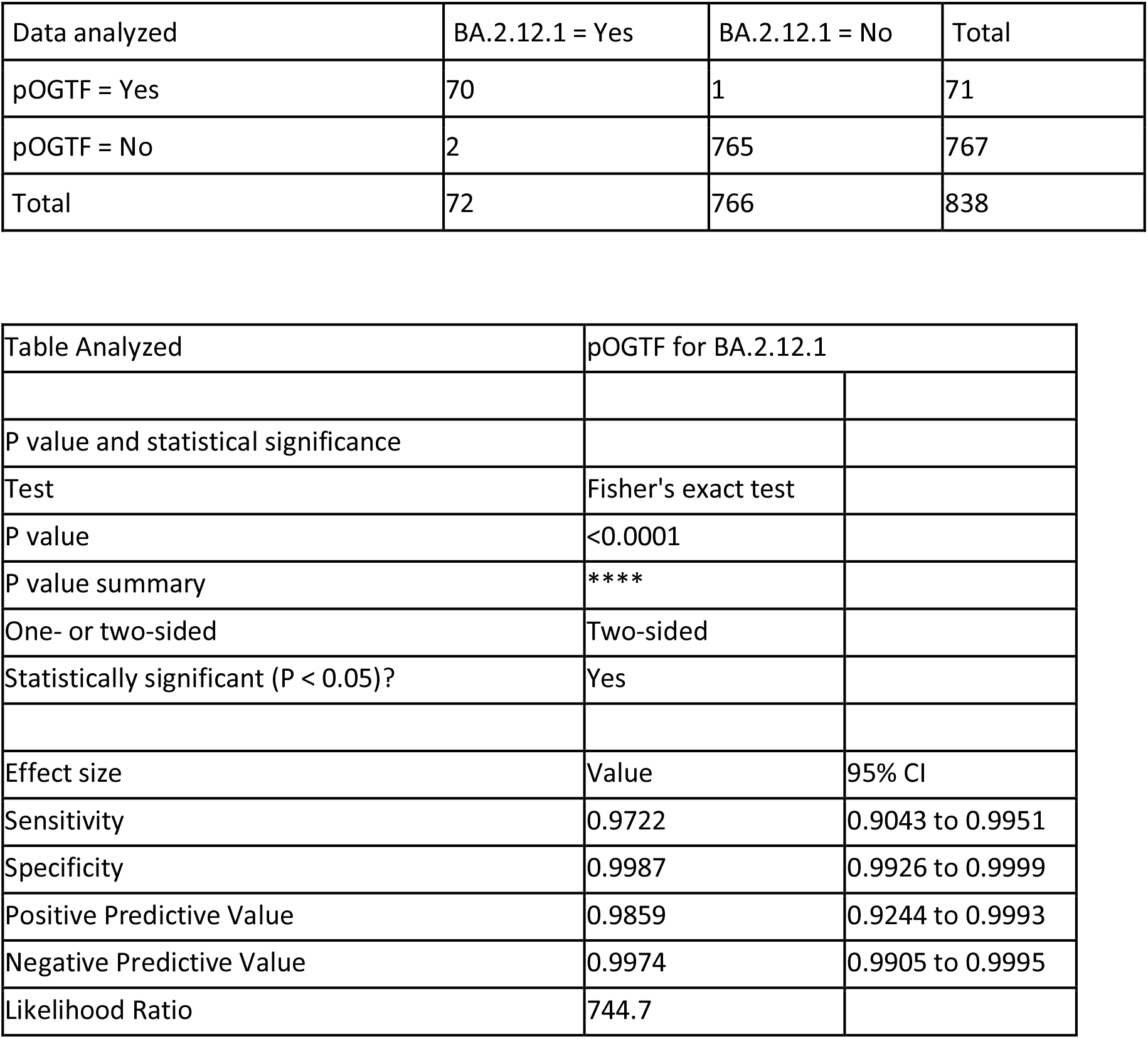
Contingency table and performance analysis using dEO threshold ≥2 for samples with E-gene C_*T*_ ≤30 as a marker of BA.2.12.1.

| Data analyzed | BA.2.12.1 = Yes | BA.2.12.1 = No | Total |
| --- | --- | --- | --- |
| pOGTF = Yes | 70 | 1 | 71 |
| pOGTF = No | 2 | 765 | 767 |
| Total | 72 | 766 | 838 |

| Table Analyzed | pOGTF for BA.2.12.1 |  |
| --- | --- | --- |
| P value and statistical significance |  |  |
| Test | Fisher's exact test |  |
| P value | <0.0001 |  |
| P value summary | **** |  |
| One- or two-sided | Two-sided |  |
| Statistically significant ($P < 0.05$)? | Yes | |
| Effect size | Value | 95% CI |
| Sensitivity | 0.9722 | 0.9043 to 0.9951 |
| Specificity | 0.9987 | 0.9926 to 0.9999 |
| Positive Predictive Value | 0.9859 | 0.9244 to 0.9993 |
| Negative Predictive Value | 0.9974 | 0.9905 to 0.9995 |
| Likelihood Ratio | 744.7 |  |

**Table 2.**
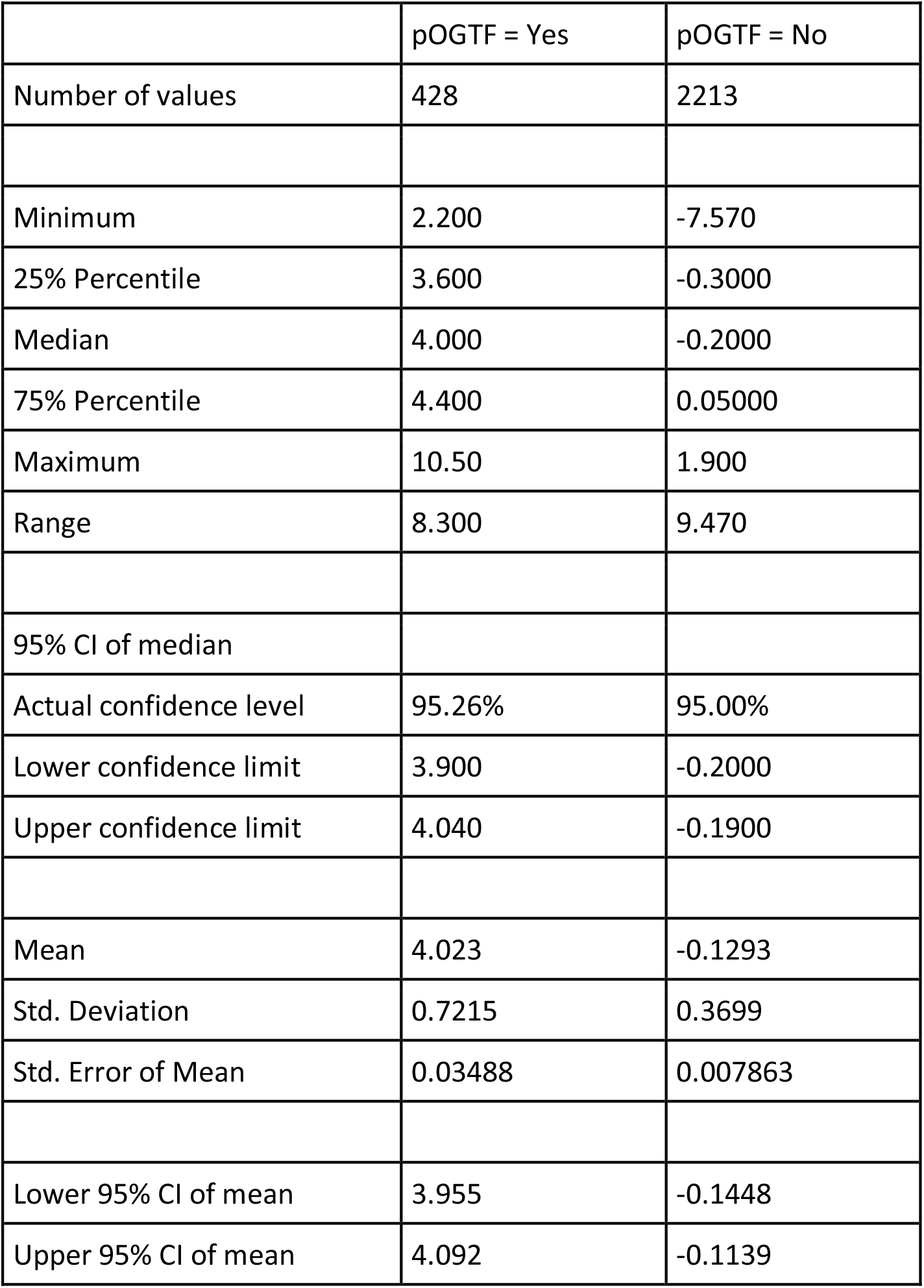
Descriptive stats for entire data set for samples with E gene C_*T*_ ≤30 and dEO ≥2 (ie. pOGTF).

**Figure 3.**
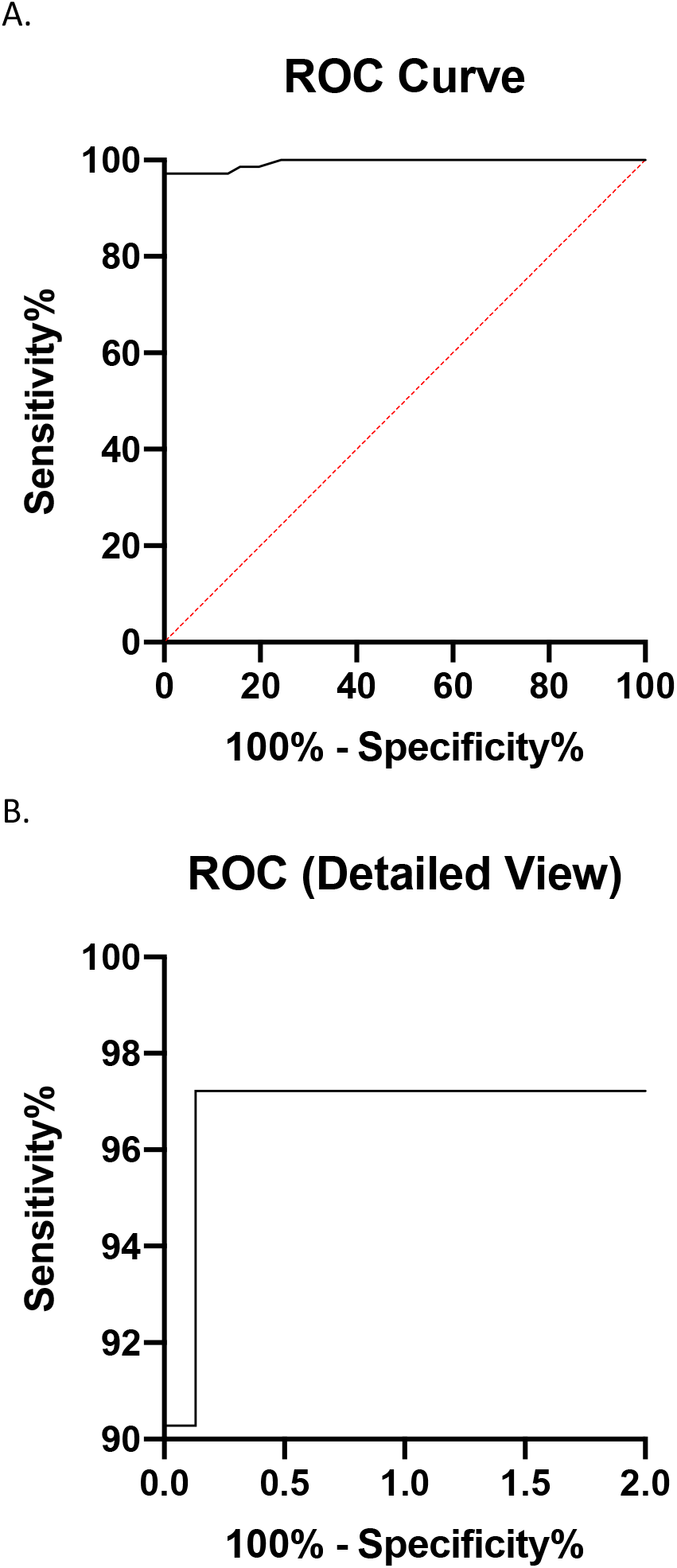
ROC curve analysis for dEO as a marker for BA.2.12.1. For samples with available sequencing data, dEO was calculated, and samples were classified as “BA.2.12.1” or “Not BA.2.12.1.” A total of 72 BA.2.12.1 and 766 other lineage samples were included. ROC analysis was performed in GraphPad Prism and full ROC Curve (A) and a detailed view (B) are shown. Only samples with E-gene C_*T*_ values ≤30 were used.

To better understand the relationship between pOGTF, the emergence of BA.2.12.1, and overall test positivity, we reviewed publicly available GISAID data to identify the number of all sequences and BA.2.12.1 sequences uploaded from North America for the same time period as our study. Over a five-week period, the number of sequences submitted was stable, but the proportion of BA.2.12.1 samples increased over time to approximately 13% of all sequences (Figure 4A). Our total testing data using the pOGTF threshold also revealed, similar rise in samples demonstrating pOGTF, with the proportion of positive samples with pOGTF steadily increasing over the study period, peaking in the final week of the study (April 10-16) with a range from 11.1% to 39.8 % among the seven study sites (Figure 4B). This rise is continuing. Importantly, the rise in pOGTF at our institutions has also corresponded to an increase in overall test positivity, consistent with concerns of increased transmissibility of the BA.2.12.1 variant.

**Figure 4.**
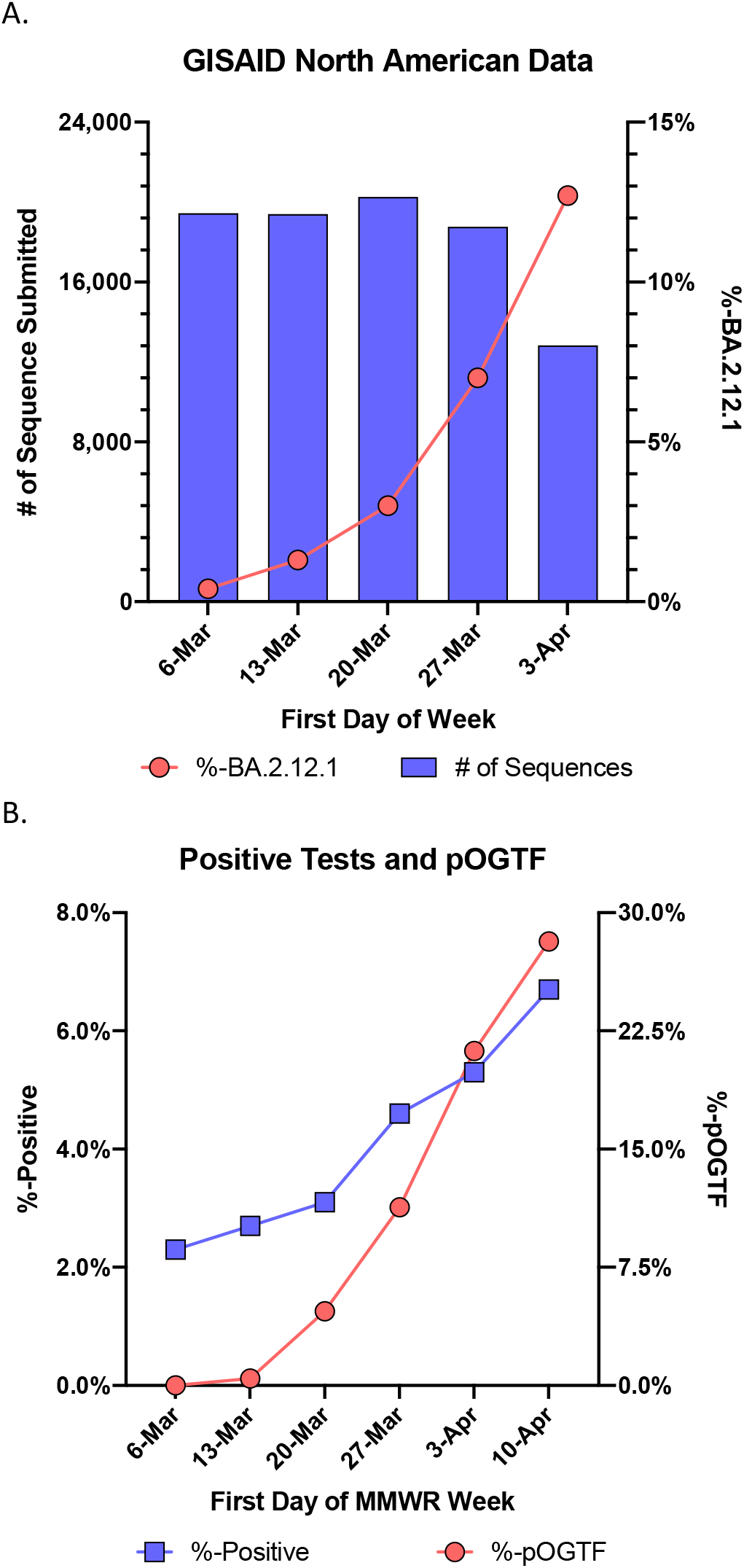
Increasing BA.2.12.1 prevalence relates to pOGTF and percent positivity. (A) All North American samples submitted to GISAID from the indicated period of time were extracted on 4/21/22, and the percentage of all sequences classified as BA.2.12.1 was calculated. (B) Percent positivity and percent pOGTF for all samples tested by cobas**^®^** SARS-CoV-2 or cobas**^®^** SARS-CoV-2 & Influenza A/B assays at participating institutions for the indicated weeks. For CCF, percent positive data included additional samples tested by an alternative platform. All samples regardless of E-gene C_*T*_ value were included. Percent pOGTF was calculated for all samples with E gene C_*T*_ values ≤30 with or without a dEO ≥2 cycles.

## Discussion

The role of C_*T*_ values in clinical decision-making has been hotly debated throughout the pandemic; however, studies have demonstrated their association with clinical outcomes and thus their potential to inform clinical and logistic decisions (20-30). Nevertheless, multiple sources of pre-analytic and analytic variability, along with the qualitative nature of SARS-CoV-2 assays, can complicate interpretation (31-33). However, with enough data points a normal dispersion between C_*T*_ values of multi-target assays can be constructed and monitored for evidence of abnormal differences in C_*T*_ values between two targets (Figure 1) (6, 7). Abnormal divergence between C_*T*_ values may indicate polymorphisms that impact detection of individual target genes, particularly when the values are well above the limit of detection (6-10). Outliers showing abnormally large differences between targets or delayed detection of a given target relative to other targets may indicate mutations impacting the primer/probe binding sites and warrant further characterization by WGS.

Tracking unique assay performance characteristics as a marker of a specific variant has demonstrated value through more rapid assessment than WGS, as has been readily apparent with SGTF in the Alpha and Omicron (BA.1 and BA.2) waves of the COVID-19 pandemic. Here we describe the use of pOGTF on the cobas^®^ SARS-CoV-2 and cobas^®^ SARS-CoV-2 & Influenza A/B assays to rapidly identify the emergence of BA.2.12.1. pOGTF is a highly reliable proxy for BA.2.12.1, particularly for samples with C_*T*_ values ≤30 thermocycles. This phenomenon is useful for rapid recognition, particularly for laboratories that lack sequencing capabilities. The CDC Nowcast variant proportion projection for the week ending 4/16/22 estimates BA.2.12.1 at 19.0% (13.4-26.0%) nationally, with higher proportions in the Northeast of the US (34). Our data are congruent with these projections, showing highest concentration in the US Northeast with spread into the US Midwest through mid-April, and we demonstrate that pOGTF can be used for real-time BA.2.12.1 tracking. Notably, estimates of the BA.2.12.1 variant fraction using pOGTF are higher than WGS-derived projections. This increased fraction may indicate that BA.2.12.1 is spreading more quickly than has been estimated.

The emergence of BA.2.12.1 at our study sites has coincided with increased positivity rates (Figure 4), suggesting this variant is at least partially responsible for increased community transmission. Increased transmissibility is likely attributable in part to the spike L452Q mutation present in BA.2.12.1, which is not found in the parent strain but was present in Lambda (C.37). A similar mutation, L452R was observed with Delta (B.1.617.2). L452Q has been implicated as an important driver of human transmission, enhancing infectivity and receptor binding while reducing vaccine-derived immunity (35).

For routine diagnostic laboratories, identifying mutations responsible for altering commercial assay performance is hindered by a lack of public information regarding the primer/probe target regions of commercially available FDA EUA assays. As stated during the July 26, 2021 CDC Clinical Laboratory COVID-19 Response Call, “…ideally genomic regions targeted by IVDs would be made publicly available by manufacturers, so prospective investigation of polymorphisms occurring within target regions could be identified” (36). The authors support disclosure of primer/probe target regions for FDA EUA assays to facilitate monitoring for mutations that could impact diagnostic assay performance. As in the case of pOGTF with BA.2.12.1, two putative mutations (g.11674 C>T and g.15009 T>C) have been identified, but the responsible mutation impacting the assay performance has not yet been confirmed. If the former, this supports Wang et al.’s conclusion that cytidines in assay target regions are particularly susceptible to mutations for SARS-CoV-2 (37). Additional challenges for sentinel laboratories in identifying assay-impacting mutations are the frequency of the observation and quantity of testing. Sentinel laboratories may struggle to identify an emerging pattern when restricted to local data. It may therefore be easier to identify target failure using larger data sets or by retrospectively retesting samples with mutations identified by WGS, if the genomic regions targeted by the NAAT are publicly available.

Currently, mutations impacting primer/probe binding are self-monitored by manufacturers, which may pose a conflict of interest (38). Monitoring therefore relies on publicly generated and openly shared genomic data (GISAID and Genbank). The lag in availability of WGS data may delay detection of new variants that impact assay performance by weeks. Additionally, if a variant emerges in a locale with low sequencing surveillance, recognition may be further delayed. As demonstrated herein, monitoring of assay performance through real-time evaluation of C_*T*_ values has utility in SARS-CoV-2 variant surveillance, and could inform clinical and logistical decision making. Widespread real-time uploading of SARS-CoV-2 NAAT C_*T*_ values to an FDA or manufacturer-sponsored database, similar to the data curation available for BioFire^®^ Syndromic Trends Epidemiology Tool, could permit rapid detection, and thus response, to emerging variants (39).

This study has some limitations. Not all SARS-CoV-2 tests were performed on the cobas^®^ SARS-CoV-2 assays as multiple NAAT platforms are used at each participating site, and operational workflow may have unintentionally introduced biases (e.g., directing samples from certain patient populations or need for rapid result to particular platforms) to the study set. All analyses were performed on a per-sample and not a per-subject basis, and a single subject’s samples could have been included multiple times within the data set. However, the impact of this possibility is diminished by the multi-center nature of this study and large data. While pOGTF is currently specific to BA.2.12.1, the specificity, sensitivity, and predictive values may change over time as SARS-CoV-2 continues to evolve and the lineages prevalent in the population changes. Although pOGTF can be a rapid and useful tool for monitoring BA.2.12.1, WGS remains important in confirming the lineage, assessing for additional mutations, and detecting new variants that decrease the specificity of the association of pOGTF with BA.2.12.1.

## Data Availability

All SARS-CoV-2 viral genomes have been deposited in GISAID and/or NCBI Genbank with accession numbers or viral names listed in Table S1.
dEO outlier model code and sample reproducible report are available at: https://github.com/bjklab/SARS-CoV-2_Ct_report.

## Acknowledgements

All authors would like to acknowledge the ASM ClinMicroNet Listserv for providing a platform for communication and collaboration between clinical microbiology laboratory directors, which allowed this group of authors to connect on this shared observation. KGR, BJK, FB, AM acknowledge funding provided by a contract award from the Centers for Disease Control and Prevention (CDC BAA 200-2021-10986 and 75D30121C11102/000HCVL1-2021-55232), philanthropic donations to the Penn Center for Research on Coronaviruses and Other Emerging Pathogens, and in part by NIH grant R61/33-HL137063 and AI140442 -supplement for SARS-CoV-2. Additional assistance was provided by the Penn Center for AIDS Research (P30-AI045008) and in part with Federal funds from the National Institute of Allergy and Infectious Diseases, National Institutes of Health, Department of Health and Human Services, under Contract No. 75N93021C00015. BJK is also supported by NIH K23 AI121485. They also acknowledge assistance with data collection and analysis by Daniel Danoski, Michael Kosenski, Amee Aghera, Irina Petlakh and the Penn Medicine Data Analytics Center. DAG, GJB, FW, MKA, ACU acknowledge in part NIH funding (U01 DA053949) and the Columbia University Biobank team. DDR and TZ acknowledge Ashley Figula who first recognized the pOGTF phenomenon, which prompted this study; and we thank the Ohio Department of Health for its support of Cleveland Clinic’s SARS-CoV-2 genomic surveillance. TM, KJ, NEB acknowledge funding in part through the National Institute of Heath/National Cancer Institute Cancer Center Support (Grant P30 CA008748) and Philanthropic Funds from the Burns Family. NDG acknowledges funding provided by the Centers for Disease Control and Prevention Broad Agency Announcement (75D30120C09570). LFW and RMR acknowledge Kathy Fauntleroy and Selma Salter from New York-Presbyterian/Weill Cornell Medical Center, and Melissa Cushing from Weill Cornell Medicine. We gratefully acknowledge both the originating and submitting laboratories for the sequence data in GISAID EpiCoV on which the SARS-CoV-2 variant prevalence data are partially based.

The Yale SARS-CoV-2 Genomic Surveillance Initiative consists of Kendall Billig, Mallery Breban, Chantal Vogels, Kien Pham, Nicholas Chen, Chrispin Chaguza, Irina Tikhonova, Christopher Castaldi, Shrikant Mane, Bony De Kumar, David Ferguson, Nicholas Kerantzas, Marie Landry, Wade Schulz.

