## Supplemental Table 1 for "Partial ORF1ab Gene Target Failure with Omicron BA.2.12.1"

| Unique ID | MMWR Week | E Ct | ORF1AB Ct | GISAID ID |
|---|---|---|---|---|
| CCF-0004 | 2022;10 | 18.19 | 18.36 | EPI_ISL_10942121 |
| CCF-0860 | 2022;10 | 19.23 | 18.82 | EPI_ISL_10942135 |
| CCF-0861 | 2022;10 | 22.79 | 22.18 | EPI_ISL_10942136 |
| CCF-0862 | 2022;10 | 19.07 | 19.05 | EPI_ISL_10942137 |
| CCF-0863 | 2022;10 | 19.1 | 18.82 | EPI_ISL_10942138 |
| CCF-0003 | 2022;10 | 20.94 | 21.08 | EPI_ISL_10942173 |
| CCF-0855 | 2022;10 | 20.05 | 19.68 | EPI_ISL_10942174 |
| CCF-0857 | 2022;10 | 19.45 | 19.54 | EPI_ISL_10942175 |
| CCF-0005 | 2022;10 | 21.53 | 21.42 | EPI_ISL_10942191 |
| CCF-0851 | 2022;10 | 19.42 | 19.07 | EPI_ISL_10942192 |
| CCF-0856 | 2022;10 | 19.08 | 18.89 | EPI_ISL_10942193 |
| CCF-0853 | 2022;10 | 24.78 | 24.63 | EPI_ISL_10942196 |
| CCF-0021 | 2022;10 | 17.01 | 17.74 | EPI_ISL_11169926 |
| CCF-0022 | 2022;10 | 17.04 | 17.5 | EPI_ISL_11169927 |
| CCF-0027 | 2022;10 | 20.73 | 20.47 | EPI_ISL_11169928 |
| CCF-0029 | 2022;10 | 18.47 | 18.5 | EPI_ISL_11169929 |
| CCF-0031 | 2022;10 | 20.46 | 20.63 | EPI_ISL_11169930 |
| CCF-0049 | 2022;10 | 24.56 | 24.74 | EPI_ISL_11169931 |
| CCF-0042 | 2022;10 | 24.98 | 25.01 | EPI_ISL_11169931 |
| CCF-0841 | 2022;10 | 20.6 | 20.19 | EPI_ISL_11169935 |
| CCF-0842 | 2022;10 | 24.99 | 24.37 | EPI_ISL_11169936 |
| CCF-0835 | 2022;10 | 19.93 | 19.68 | EPI_ISL_11169937 |
| CCF-0829 | 2022;10 | 22.69 | 22.19 | EPI_ISL_11169938 |
| CCF-0039 | 2022;10 | 15.73 | 16.14 | EPI_ISL_11169940 |
| CCF-0050 | 2022;10 | 24.43 | 24.55 | EPI_ISL_11169941 |
| CCF-0041 | 2022;10 | 25.57 | 25.44 | EPI_ISL_11169941 |
| CCF-0019 | 2022;10 | 23.03 | 23.16 | EPI_ISL_11169944 |
| CCF-0020 | 2022;10 | 21.26 | 21.42 | EPI_ISL_11169945 |
| CCF-0023 | 2022;10 | 21.32 | 21.35 | EPI_ISL_11169946 |
| CCF-0024 | 2022;10 | 18.21 | 18.37 | EPI_ISL_11169947 |
| CCF-0025 | 2022;10 | 19.77 | 19.55 | EPI_ISL_11169948 |
| CCF-0030 | 2022;10 | 19.79 | 19.89 | EPI_ISL_11169949 |
| CCF-0037 | 2022;10 | 22.64 | 23.33 | EPI_ISL_11169950 |
| CCF-0047 | 2022;10 | 22.89 | 23.02 | EPI_ISL_11169951 |
| CCF-0044 | 2022;10 | 23.33 | 23.43 | EPI_ISL_11169951 |
| CCF-0048 | 2022;10 | 18.8 | 19.32 | EPI_ISL_11169952 |
| CCF-0043 | 2022;10 | 19.66 | 20.16 | EPI_ISL_11169952 |
| CCF-0051 | 2022;10 | 19.63 | 19.39 | EPI_ISL_11169953 |
| CCF-0053 | 2022;10 | 18.9 | 19.61 | EPI_ISL_11169954 |
| CCF-0054 | 2022;10 | 17.54 | 17.41 | EPI_ISL_11169955 |
| CCF-0057 | 2022;10 | 20.42 | 20.38 | EPI_ISL_11169956 |
| CCF-0062 | 2022;10 | 19.65 | 19.71 | EPI_ISL_11169957 |

| | | | | |
|---|---|---|---|---|
| CCF-0063 | 2022;10 | 17.01 | 17.4 | EPI_ISL_11169958 |
| CCF-0066 | 2022;11 | 24.74 | 25.09 | EPI_ISL_11169959 |
| CCF-0068 | 2022;11 | 24.15 | 24.09 | EPI_ISL_11169960 |
| CCF-0070 | 2022;11 | 18.12 | 18.25 | EPI_ISL_11169961 |
| CCF-0071 | 2022;11 | 22.71 | 22.91 | EPI_ISL_11169962 |
| CCF-0845 | 2022;10 | 19.41 | 19.12 | EPI_ISL_11169976 |
| CCF-0846 | 2022;10 | 18.95 | 18.44 | EPI_ISL_11169977 |
| CCF-0839 | 2022;10 | 23.95 | 23.37 | EPI_ISL_11169978 |
| CCF-0833 | 2022;10 | 23.81 | 23.35 | EPI_ISL_11169979 |
| CCF-0830 | 2022;10 | 20.69 | 20.51 | EPI_ISL_11169980 |
| CCF-0828 | 2022;10 | 22.99 | 22.55 | EPI_ISL_11169981 |
| CCF-0827 | 2022;10 | 20.21 | 20.01 | EPI_ISL_11169982 |
| CCF-0824 | 2022;11 | 23.67 | 23.26 | EPI_ISL_11169983 |
| CCF-0065 | 2022;10 | 21.62 | 21.92 | EPI_ISL_11169985 |
| CCF-0034 | 2022;10 | 24.19 | 24.09 | EPI_ISL_11169988 |
| CCF-0064 | 2022;10 | 17.92 | 18.12 | EPI_ISL_11169991 |
| CCF-0060 | 2022;10 | 26.01 | 26.22 | EPI_ISL_11169993 |
| CCF-0074 | 2022;12 | 24.25 | 24.24 | EPI_ISL_11378423 |
| CCF-0077 | 2022;12 | 19.19 | 19.3 | EPI_ISL_11378424 |
| CCF-0139 | 2022;11 | 17.45 | 17.21 | EPI_ISL_11378425 |
| CCF-0821 | 2022;11 | 18.79 | 18.64 | EPI_ISL_11378426 |
| CCF-0793 | 2022;12 | 22.43 | 22.08 | EPI_ISL_11378427 |
| CCF-0128 | 2022;11 | 16.55 | 16.52 | EPI_ISL_11378428 |
| CCF-0123 | 2022;11 | 22.59 | 23.15 | EPI_ISL_11378429 |
| CCF-0124 | 2022;11 | 24.44 | 25.08 | EPI_ISL_11378430 |
| CCF-0115 | 2022;11 | 18.49 | 18.95 | EPI_ISL_11378432 |
| CCF-0116 | 2022;11 | 20.03 | 19.94 | EPI_ISL_11378433 |
| CCF-0118 | 2022;11 | 23.91 | 23.86 | EPI_ISL_11378434 |
| CCF-0119 | 2022;11 | 17.29 | 17.14 | EPI_ISL_11378435 |
| CCF-0112 | 2022;11 | 18.8 | 18.75 | EPI_ISL_11378436 |
| CCF-0102 | 2022;11 | 15.62 | 15.97 | EPI_ISL_11378438 |
| CCF-0093 | 2022;11 | 25.54 | 25.71 | EPI_ISL_11378439 |
| CCF-0084 | 2022;11 | 18.79 | 18.8 | EPI_ISL_11378442 |
| CCF-0797 | 2022;12 | 19.69 | 19.63 | EPI_ISL_11378443 |
| CCF-0804 | 2022;11 | 19.48 | 19.06 | EPI_ISL_11378444 |
| CCF-0805 | 2022;11 | 19.77 | 19.42 | EPI_ISL_11378445 |
| CCF-0806 | 2022;11 | 19.02 | 19 | EPI_ISL_11378446 |
| CCF-0813 | 2022;11 | 21.2 | 20.94 | EPI_ISL_11378447 |
| CCF-0814 | 2022;11 | 16.59 | 16.37 | EPI_ISL_11378448 |
| CCF-0141 | 2022;11 | 21.39 | 21.45 | EPI_ISL_11378466 |
| CCF-0819 | 2022;11 | 17.95 | 17.81 | EPI_ISL_11378467 |
| CCF-0126 | 2022;11 | 20.35 | 20.49 | EPI_ISL_11378471 |
| CCF-0114 | 2022;11 | 16.17 | 16.22 | EPI_ISL_11378474 |

| | | | | |
|---|---|---|---|---|
| CCF-0117 | 2022;11 | 19.57 | 19.72 | EPI_ISL_11378475 |
| CCF-0111 | 2022;11 | 19.7 | 20.56 | EPI_ISL_11378477 |
| CCF-0107 | 2022;11 | 21.17 | 21.64 | EPI_ISL_11378478 |
| CCF-0099 | 2022;11 | 19.93 | 20.47 | EPI_ISL_11378481 |
| CCF-0095 | 2022;11 | 23.97 | 24.09 | EPI_ISL_11378482 |
| CCF-0090 | 2022;11 | 19.66 | 19.54 | EPI_ISL_11378484 |
| CCF-0081 | 2022;11 | 19.54 | 19.54 | EPI_ISL_11378485 |
| CCF-0085 | 2022;11 | 20.45 | 20.28 | EPI_ISL_11378488 |
| CCF-0079 | 2022;12 | 20.39 | 21.33 | EPI_ISL_11378489 |
| CCF-0800 | 2022;11 | 19.03 | 18.41 | EPI_ISL_11378490 |
| CCF-0803 | 2022;11 | 21.18 | 21.02 | EPI_ISL_11378491 |
| CCF-0807 | 2022;11 | 21.92 | 21.61 | EPI_ISL_11378492 |
| CCF-0812 | 2022;11 | 24.66 | 24.21 | EPI_ISL_11378495 |
| CCF-0104 | 2022;11 | 19.83 | 19.92 | EPI_ISL_11378499 |
| CCF-0091 | 2022;11 | 24.74 | 24.65 | EPI_ISL_11378501 |
| CCF-0140 | 2022;11 | 28.29 | 27.74 | EPI_ISL_11378504 |
| YNHH-0033 | 2022;10 | 23.4 | 23.1 | EPI_ISL_11503890 |
| YNHH-0035 | 2022;10 | 18.3 | 18.1 | EPI_ISL_11503891 |
| YNHH-0049 | 2022;10 | 25.1 | 24.9 | EPI_ISL_11503898 |
| YNHH-0034 | 2022;10 | 21.4 | 21.1 | EPI_ISL_11503899 |
| YNHH-0031 | 2022;10 | 31 | 30.4 | EPI_ISL_11503914 |
| YNHH-0040 | 2022;10 | 27.8 | 27.4 | EPI_ISL_11503915 |
| YNHH-0056 | 2022;10 | 20.5 | 20.3 | EPI_ISL_11576284 |
| YNHH-0055 | 2022;10 | 24 | 24.4 | EPI_ISL_11576285 |
| YNHH-0061 | 2022;10 | 20.9 | 20.6 | EPI_ISL_11576286 |
| YNHH-0060 | 2022;10 | 18.6 | 18.7 | EPI_ISL_11576287 |
| YNHH-0043 | 2022;10 | 29.3 | 28.9 | EPI_ISL_11576288 |
| YNHH-0046 | 2022;10 | 20 | 19.9 | EPI_ISL_11576289 |
| YNHH-0059 | 2022;10 | 19.8 | 19.6 | EPI_ISL_11576290 |
| YNHH-0052 | 2022;10 | 27.4 | 26.7 | EPI_ISL_11576295 |
| YNHH-0077 | 2022;10 | 21.1 | 20.9 | EPI_ISL_11576302 |
| YNHH-0069 | 2022;10 | 20.4 | 20.3 | EPI_ISL_11576303 |
| YNHH-0076 | 2022;10 | 28.1 | 28 | EPI_ISL_11576304 |
| YNHH-0070 | 2022;10 | 25 | 24.7 | EPI_ISL_11576305 |
| YNHH-0089 | 2022;10 | 18.6 | 18.4 | EPI_ISL_11576317 |
| YNHH-0114 | 2022;11 | 26.2 | 25.6 | EPI_ISL_11576318 |
| YNHH-0090 | 2022;10 | 17.9 | 17.5 | EPI_ISL_11576320 |
| YNHH-0095 | 2022;10 | 23.9 | 23.9 | EPI_ISL_11576321 |
| YNHH-0122 | 2022;11 | 24.5 | 24.2 | EPI_ISL_11576322 |
| YNHH-0110 | 2022;10 | 20.4 | 20.4 | EPI_ISL_11576324 |
| YNHH-0098 | 2022;10 | 21.8 | 21.5 | EPI_ISL_11576327 |
| YNHH-0111 | 2022;11 | 22.3 | 21.7 | EPI_ISL_11576329 |
| YNHH-0121 | 2022;11 | 20 | 19.6 | EPI_ISL_11576331 |

| | | | | |
|---|---|---|---|---|
| YNHH-0117 | 2022;11 | 19.6 | 19.3 | EPI_ISL_11576333 |
| YNHH-0106 | 2022;10 | 20.5 | 20.2 | EPI_ISL_11576337 |
| YNHH-0116 | 2022;11 | 19.1 | 18.5 | EPI_ISL_11576338 |
| YNHH-0115 | 2022;11 | 29.3 | 28.6 | EPI_ISL_11576340 |
| YNHH-0088 | 2022;10 | 18.6 | 18.4 | EPI_ISL_11576341 |
| YNHH-0084 | 2022;10 | 19 | 18.8 | EPI_ISL_11576343 |
| YNHH-0085 | 2022;10 | 21.6 | 21.4 | EPI_ISL_11576344 |
| YNHH-0125 | 2022;11 | 30.6 | 30.1 | EPI_ISL_11576345 |
| YNHH-0079 | 2022;10 | 20.2 | 19.8 | EPI_ISL_11576347 |
| YNHH-0082 | 2022;10 | 19.3 | 18.7 | EPI_ISL_11576348 |
| YNHH-0087 | 2022;10 | 22.2 | 21.8 | EPI_ISL_11576349 |
| YNHH-0096 | 2022;10 | 28.2 | 27.8 | EPI_ISL_11576350 |
| YNHH-0099 | 2022;10 | 21.8 | 21.5 | EPI_ISL_11576352 |
| YNHH-0128 | 2022;11 | 30.3 | 29.3 | EPI_ISL_11576353 |
| YNHH-0105 | 2022;10 | 20.3 | 20 | EPI_ISL_11576355 |
| YNHH-0119 | 2022;11 | 22.9 | 22.3 | EPI_ISL_11576356 |
| YNHH-0072 | 2022;10 | 29.6 | 29.1 | EPI_ISL_11576357 |
| YNHH-0100 | 2022;10 | 20.8 | 20.2 | EPI_ISL_11576358 |
| YNHH-0097 | 2022;10 | 18.8 | 18.5 | EPI_ISL_11576359 |
| YNHH-0109 | 2022;10 | 27.9 | 27.5 | EPI_ISL_11576360 |
| YNHH-0083 | 2022;10 | 29.3 | 28.9 | EPI_ISL_11576361 |
| YNHH-0086 | 2022;10 | 30.7 | 30.1 | EPI_ISL_11576362 |
| YNHH-0152 | 2022;11 | 23.8 | 23.4 | EPI_ISL_11576363 |
| YNHH-0133 | 2022;11 | 22.5 | 21.9 | EPI_ISL_11576364 |
| YNHH-0112 | 2022;11 | 23.1 | 22.8 | EPI_ISL_11576365 |
| YNHH-0137 | 2022;11 | 19.8 | 19.6 | EPI_ISL_11576366 |
| YNHH-0130 | 2022;11 | 28.2 | 27.8 | EPI_ISL_11576367 |
| YNHH-0135 | 2022;11 | 20.5 | 20.6 | EPI_ISL_11576368 |
| YNHH-0150 | 2022;11 | 22.4 | 22.2 | EPI_ISL_11576371 |
| YNHH-0148 | 2022;11 | 20.1 | 19.7 | EPI_ISL_11576372 |
| YNHH-0123 | 2022;11 | 29.2 | 28.6 | EPI_ISL_11576373 |
| YNHH-0129 | 2022;11 | 20.9 | 20.5 | EPI_ISL_11576374 |
| YNHH-0131 | 2022;11 | 19.5 | 19.2 | EPI_ISL_11576375 |
| YNHH-0143 | 2022;11 | 23.9 | 23.1 | EPI_ISL_11576376 |
| YNHH-0132 | 2022;11 | 19.3 | 18.9 | EPI_ISL_11576377 |
| YNHH-0127 | 2022;11 | 16.1 | 15.8 | EPI_ISL_11576385 |
| YNHH-0160 | 2022;11 | 21.8 | 21.6 | EPI_ISL_11576388 |
| YNHH-0154 | 2022;11 | 25.1 | 24.5 | EPI_ISL_11576389 |
| YNHH-0163 | 2022;11 | 21.5 | 21.4 | EPI_ISL_11576390 |
| YNHH-0155 | 2022;11 | 18.7 | 18.6 | EPI_ISL_11576391 |
| YNHH-0165 | 2022;11 | 22.8 | 22.6 | EPI_ISL_11576392 |
| YNHH-0171 | 2022;11 | 24.7 | 24.2 | EPI_ISL_11576394 |
| YNHH-0158 | 2022;11 | 20.4 | 20.1 | EPI_ISL_11576395 |

| | | | | |
|---|---|---|---|---|
| YNHH-0145 | 2022;11 | 20.7 | 20.4 | EPI_ISL_11576396 |
| YNHH-0166 | 2022;11 | 27.8 | 27.3 | EPI_ISL_11576397 |
| YNHH-0161 | 2022;11 | 17.7 | 17.7 | EPI_ISL_11576398 |
| YNHH-0170 | 2022;11 | 31.4 | 30.3 | EPI_ISL_11576399 |
| YNHH-0159 | 2022;11 | 23.9 | 23.6 | EPI_ISL_11576401 |
| YNHH-0156 | 2022;11 | 19.4 | 19.3 | EPI_ISL_11576404 |
| YNHH-0173 | 2022;11 | 24.8 | 24.6 | EPI_ISL_11576405 |
| YNHH-0157 | 2022;11 | 19.6 | 19.4 | EPI_ISL_11576407 |
| YNHH-0146 | 2022;11 | 24.2 | 24.1 | EPI_ISL_11576409 |
| YNHH-0149 | 2022;11 | 24.8 | 24.4 | EPI_ISL_11576410 |
| YNHH-0140 | 2022;11 | 23.6 | 23.4 | EPI_ISL_11576411 |
| YNHH-0168 | 2022;11 | 18.8 | 18.8 | EPI_ISL_11576416 |
| YNHH-0169 | 2022;11 | 22.2 | 21.9 | EPI_ISL_11576417 |
| Columbia 54 | 2022;12 | 21.3 | 21.3 | EPI_ISL_11680561 |
| Columbia 61 | 2022;12 | 18.4 | 18.6 | EPI_ISL_11680562 |
| CCF-0794 | 2022;12 | 24.07 | 23.76 | EPI_ISL_11695304 |
| CCF-0144 | 2022;12 | 20.87 | 21.01 | EPI_ISL_11695305 |
| CCF-0145 | 2022;12 | 22.31 | 22.22 | EPI_ISL_11695306 |
| CCF-0147 | 2022;12 | 21.14 | 21.14 | EPI_ISL_11695308 |
| CCF-0148 | 2022;12 | 19.65 | 19.37 | EPI_ISL_11695309 |
| CCF-0153 | 2022;12 | 19.19 | 19.89 | EPI_ISL_11695310 |
| CCF-0155 | 2022;12 | 26.29 | 26.33 | EPI_ISL_11695311 |
| CCF-0156 | 2022;12 | 22.28 | 22.37 | EPI_ISL_11695312 |
| CCF-0159 | 2022;12 | 26.07 | 26.9 | EPI_ISL_11695313 |
| CCF-0161 | 2022;12 | 21.42 | 21.47 | EPI_ISL_11695314 |
| CCF-0163 | 2022;12 | 17.48 | 17.6 | EPI_ISL_11695315 |
| CCF-0164 | 2022;12 | 16.7 | 16.76 | EPI_ISL_11695316 |
| CCF-0167 | 2022;12 | 22.79 | 23.23 | EPI_ISL_11695317 |
| CCF-0168 | 2022;12 | 22.86 | 23.59 | EPI_ISL_11695318 |
| CCF-0181 | 2022;12 | 26.52 | 26.3 | EPI_ISL_11695319 |
| CCF-0183 | 2022;12 | 27.27 | 27.24 | EPI_ISL_11695320 |
| CCF-0184 | 2022;12 | 24.41 | 24.44 | EPI_ISL_11695321 |
| CCF-0186 | 2022;12 | 25 | 25.07 | EPI_ISL_11695322 |
| CCF-0190 | 2022;12 | 17.2 | 17.36 | EPI_ISL_11695323 |
| CCF-0192 | 2022;12 | 15.91 | 16.05 | EPI_ISL_11695324 |
| CCF-0193 | 2022;12 | 17.75 | 22.59 | EPI_ISL_11695325 |
| CCF-0194 | 2022;12 | 24.3 | 24.4 | EPI_ISL_11695326 |
| CCF-0195 | 2022;12 | 16.52 | 16.17 | EPI_ISL_11695327 |
| CCF-0198 | 2022;13 | 26.62 | 27.59 | EPI_ISL_11695328 |
| CCF-0199 | 2022;13 | 22.65 | 22.61 | EPI_ISL_11695329 |
| CCF-0203 | 2022;13 | 19.69 | 19.78 | EPI_ISL_11695330 |
| CCF-0782 | 2022;12 | 28.77 | 28.04 | EPI_ISL_11695331 |
| CCF-0786 | 2022;12 | 17.86 | 17.74 | EPI_ISL_11695332 |

| | | | | |
|---|---|---|---|---|
| CCF-0787 | 2022;12 | 21.58 | 21.15 | EPI_ISL_11695333 |
| CCF-0777 | 2022;12 | 22.9 | 22.58 | EPI_ISL_11695334 |
| CCF-0775 | 2022;12 | 21.41 | 21.03 | EPI_ISL_11695335 |
| CCF-0769 | 2022;12 | 28.32 | 27.88 | EPI_ISL_11695336 |
| CCF-0766 | 2022;12 | 15.65 | 15.37 | EPI_ISL_11695337 |
| CCF-0760 | 2022;13 | 23.65 | 23.46 | EPI_ISL_11695338 |
| CCF-0208 | 2022;13 | 24.23 | 24.36 | EPI_ISL_11695350 |
| CCF-0753 | 2022;13 | 20.34 | 19.86 | EPI_ISL_11695351 |
| CCF-0143 | 2022;12 | 21.9 | 22.11 | EPI_ISL_11695352 |
| CCF-0149 | 2022;12 | 20.13 | 20.34 | EPI_ISL_11695353 |
| CCF-0166 | 2022;12 | 24.06 | 24.17 | EPI_ISL_11695356 |
| CCF-0188 | 2022;12 | 18.45 | 18.3 | EPI_ISL_11695359 |
| CCF-0189 | 2022;12 | 20.48 | 20.23 | EPI_ISL_11695360 |
| CCF-0191 | 2022;12 | 18.57 | 18.48 | EPI_ISL_11695361 |
| CCF-0202 | 2022;13 | 21.53 | 21.44 | EPI_ISL_11695363 |
| CCF-0783 | 2022;12 | 18 | 17.71 | EPI_ISL_11695364 |
| CCF-0774 | 2022;12 | 17.95 | 18.03 | EPI_ISL_11695367 |
| CCF-0773 | 2022;12 | 19.97 | 19.67 | EPI_ISL_11695368 |
| CCF-0767 | 2022;12 | 23.46 | 22.62 | EPI_ISL_11695369 |
| CCF-0768 | 2022;12 | 20.22 | 19.83 | EPI_ISL_11695370 |
| CCF-0205 | 2022;13 | 21.97 | 22.22 | EPI_ISL_11695377 |
| CCF-0150 | 2022;12 | 16.09 | 16.08 | EPI_ISL_11695378 |
| CCF-0179 | 2022;12 | 20.83 | 20.66 | EPI_ISL_11695380 |
| CCF-0776 | 2022;12 | 23.23 | 22.68 | EPI_ISL_11695381 |
| CCF-0763 | 2022;12 | 23.7 | 23.26 | EPI_ISL_11695382 |
| YNHH-0273 | 2022;12 | 23.8 | 23.6 | EPI_ISL_11812115 |
| YNHH-0284 | 2022;12 | 23.3 | 22.6 | EPI_ISL_11812116 |
| YNHH-0264 | 2022;12 | 15.6 | 15.3 | EPI_ISL_11812120 |
| YNHH-0244 | 2022;11 | 23.8 | 23.3 | EPI_ISL_11812126 |
| YNHH-0188 | 2022;11 | 24.6 | 24.7 | EPI_ISL_11812129 |
| YNHH-0189 | 2022;11 | 25.6 | 25.2 | EPI_ISL_11812132 |
| YNHH-0232 | 2022;11 | 26.4 | 25.8 | EPI_ISL_11812133 |
| YNHH-0253 | 2022;11 | 25.8 | 25.3 | EPI_ISL_11812138 |
| YNHH-0231 | 2022;11 | 21.8 | 21.3 | EPI_ISL_11812143 |
| YNHH-0265 | 2022;12 | 25.5 | 25 | EPI_ISL_11812145 |
| YNHH-0248 | 2022;11 | 20 | 19.8 | EPI_ISL_11812146 |
| YNHH-0305 | 2022;12 | 19.4 | 19.2 | EPI_ISL_11812147 |
| YNHH-0300 | 2022;12 | 21.5 | 21.4 | EPI_ISL_11812150 |
| YNHH-0229 | 2022;11 | 21.7 | 21.2 | EPI_ISL_11812156 |
| YNHH-0274 | 2022;12 | 18.3 | 18 | EPI_ISL_11812157 |
| YNHH-0254 | 2022;11 | 21.3 | 20.9 | EPI_ISL_11812158 |
| YNHH-0344 | 2022;12 | 25.4 | 24.6 | EPI_ISL_11812159 |
| YNHH-0297 | 2022;12 | 20.8 | 20.4 | EPI_ISL_11812160 |

| | | | | |
|---|---|---|---|---|
| YNHH-0252 | 2022;11 | 19.9 | 19.4 | EPI_ISL_11812161 |
| YNHH-0261 | 2022;11 | 22 | 21.8 | EPI_ISL_11812162 |
| YNHH-0320 | 2022;12 | 21.1 | 20.8 | EPI_ISL_11812164 |
| YNHH-0270 | 2022;12 | 17.7 | 17.3 | EPI_ISL_11812166 |
| YNHH-0246 | 2022;11 | 20.4 | 20.1 | EPI_ISL_11812167 |
| YNHH-0224 | 2022;11 | 18.2 | 18 | EPI_ISL_11812169 |
| YNHH-0219 | 2022;11 | 22.7 | 22.4 | EPI_ISL_11812173 |
| YNHH-0193 | 2022;11 | 17.8 | 17.8 | EPI_ISL_11812174 |
| YNHH-0218 | 2022;11 | 25.7 | 25.5 | EPI_ISL_11812178 |
| YNHH-0182 | 2022;11 | 18.3 | 18 | EPI_ISL_11812180 |
| YNHH-0288 | 2022;12 | 24.2 | 23.9 | EPI_ISL_11812181 |
| YNHH-0290 | 2022;12 | 24.2 | 23.9 | EPI_ISL_11812182 |
| YNHH-0242 | 2022;11 | 18.9 | 18.7 | EPI_ISL_11812183 |
| YNHH-0306 | 2022;12 | 20.7 | 20.4 | EPI_ISL_11812184 |
| YNHH-0294 | 2022;12 | 20.2 | 20.1 | EPI_ISL_11812185 |
| YNHH-0295 | 2022;12 | 22.1 | 21.6 | EPI_ISL_11812186 |
| YNHH-0312 | 2022;12 | 24.1 | 23.8 | EPI_ISL_11812188 |
| YNHH-0225 | 2022;11 | 17.6 | 17.2 | EPI_ISL_11812191 |
| YNHH-0286 | 2022;12 | 23.5 | 23.3 | EPI_ISL_11812192 |
| YNHH-0282 | 2022;12 | 17.5 | 17.6 | EPI_ISL_11812193 |
| YNHH-0335 | 2022;12 | 21.9 | 21.5 | EPI_ISL_11812194 |
| YNHH-0302 | 2022;12 | 16.9 | 16.7 | EPI_ISL_11812195 |
| YNHH-0342 | 2022;12 | 17.2 | 16.9 | EPI_ISL_11812197 |
| YNHH-0215 | 2022;11 | 22 | 21.5 | EPI_ISL_11812198 |
| YNHH-0291 | 2022;12 | 23.1 | 22.8 | EPI_ISL_11812199 |
| YNHH-0277 | 2022;12 | 23 | 22.7 | EPI_ISL_11812201 |
| YNHH-0259 | 2022;11 | 20 | 19.4 | EPI_ISL_11812203 |
| YNHH-0200 | 2022;11 | 21.2 | 20.7 | EPI_ISL_11812209 |
| YNHH-0202 | 2022;11 | 29.8 | 29.4 | EPI_ISL_11812212 |
| YNHH-0237 | 2022;11 | 21.1 | 21.1 | EPI_ISL_11812216 |
| YNHH-0266 | 2022;12 | 18 | 17.6 | EPI_ISL_11812217 |
| YNHH-0263 | 2022;11 | 19.4 | 19.2 | EPI_ISL_11812219 |
| YNHH-0247 | 2022;11 | 24.9 | 24.1 | EPI_ISL_11812220 |
| YNHH-0001 | 2022;11 | 23.4 | 23.1 | EPI_ISL_11812221 |
| YNHH-0238 | 2022;11 | 26.7 | 26.1 | EPI_ISL_11812222 |
| YNHH-0256 | 2022;11 | 20.5 | 20.5 | EPI_ISL_11812223 |
| YNHH-0236 | 2022;11 | 20.3 | 20.1 | EPI_ISL_11812224 |
| YNHH-0239 | 2022;11 | 18.4 | 18.5 | EPI_ISL_11812225 |
| YNHH-0260 | 2022;11 | 23.4 | 22.9 | EPI_ISL_11812226 |
| YNHH-0308 | 2022;12 | 21.4 | 20.8 | EPI_ISL_11812227 |
| YNHH-0314 | 2022;12 | 23.2 | 22.8 | EPI_ISL_11812228 |
| YNHH-0220 | 2022;11 | 19.8 | 19.6 | EPI_ISL_11812229 |
| YNHH-0186 | 2022;11 | 20.6 | 20.2 | EPI_ISL_11812231 |

| | | | | |
|---|---|---|---|---|
| YNHH-0181 | 2022;11 | 25.9 | 25.7 | EPI_ISL_11812232 |
| YNHH-0296 | 2022;12 | 20.5 | 20.5 | EPI_ISL_11812234 |
| YNHH-0339 | 2022;12 | 18.6 | 18.6 | EPI_ISL_11812235 |
| YNHH-0276 | 2022;12 | 21.9 | 21.4 | EPI_ISL_11812236 |
| YNHH-0310 | 2022;12 | 31.8 | 26 | EPI_ISL_11812237 |
| YNHH-0207 | 2022;11 | 23.1 | 23.2 | EPI_ISL_11812238 |
| YNHH-0283 | 2022;12 | 30.9 | 30.5 | EPI_ISL_11812239 |
| YNHH-0311 | 2022;12 | 31 | 30.4 | EPI_ISL_11812241 |
| YNHH-0279 | 2022;12 | 29.6 | 29.1 | EPI_ISL_11812243 |
| YNHH-0304 | 2022;12 | 30.1 | 29.6 | EPI_ISL_11812244 |
| YNHH-0208 | 2022;11 | 26.8 | 26.5 | EPI_ISL_11812245 |
| YNHH-0272 | 2022;12 | 29.3 | 28.7 | EPI_ISL_11812247 |
| YNHH-0333 | 2022;12 | 27.8 | 27.5 | EPI_ISL_11812249 |
| YNHH-0192 | 2022;11 | 28.5 | 27.8 | EPI_ISL_11812251 |
| YNHH-0258 | 2022;11 | 26.4 | 26.2 | EPI_ISL_11812254 |
| YNHH-0262 | 2022;11 | 22 | 21.8 | EPI_ISL_11812255 |
| YNHH-0187 | 2022;11 | 27.4 | 26.9 | EPI_ISL_11812256 |
| YNHH-0289 | 2022;12 | 27.7 | 27.2 | EPI_ISL_11812257 |
| YNHH-0180 | 2022;11 | 29.7 | 29.1 | EPI_ISL_11812259 |
| YNHH-0195 | 2022;11 | 23.1 | 22.6 | EPI_ISL_11812260 |
| YNHH-0228 | 2022;11 | 24.6 | 24.3 | EPI_ISL_11812262 |
| YNHH-0278 | 2022;12 | 27.5 | 27.2 | EPI_ISL_11812264 |
| YNHH-0275 | 2022;12 | 27.1 | 26.7 | EPI_ISL_11812269 |
| YNHH-0326 | 2022;12 | 29.5 | 29 | EPI_ISL_11812270 |
| YNHH-0216 | 2022;11 | 30.6 | 29.9 | EPI_ISL_11812274 |
| YNHH-0183 | 2022;11 | 29.3 | 28.7 | EPI_ISL_11812275 |
| YNHH-0303 | 2022;12 | 29.2 | 28.5 | EPI_ISL_11812278 |
| YNHH-0269 | 2022;12 | 27.7 | 27.1 | EPI_ISL_11812279 |
| YNHH-0223 | 2022;11 | 30.2 | 29.5 | EPI_ISL_11812280 |
| YNHH-0185 | 2022;11 | 27.9 | 27.4 | EPI_ISL_11812282 |
| YNHH-0190 | 2022;11 | 26.5 | 26.1 | EPI_ISL_11812283 |
| YNHH-0285 | 2022;12 | 29.9 | 29.2 | EPI_ISL_11812284 |
| YNHH-0243 | 2022;11 | 30.9 | 29.8 | EPI_ISL_11812286 |
| YNHH-0234 | 2022;11 | 29.1 | 28.6 | EPI_ISL_11812289 |
| YNHH-0175 | 2022;11 | 24.7 | 24.3 | EPI_ISL_11812291 |
| YNHH-0217 | 2022;11 | 25.5 | 25 | EPI_ISL_11812295 |
| CCF-0227 | 2022;13 | 26.5 | 26.51 | EPI_ISL_11889657 |
| CCF-0263 | 2022;14 | 25.34 | 25.61 | EPI_ISL_11889667 |
| CCF-0264 | 2022;14 | 26.04 | 30.6 | EPI_ISL_11889668 |
| CCF-0265 | 2022;14 | 17.57 | 18.11 | EPI_ISL_11889669 |
| CCF-0266 | 2022;14 | 23.96 | 27.84 | EPI_ISL_11889670 |
| CCF-0268 | 2022;14 | 25.61 | 25.22 | EPI_ISL_11889671 |
| CCF-0261 | 2022;13 | 23.16 | 23.13 | EPI_ISL_11889672 |

| | | | | |
|---|---|---|---|---|
| CCF-0259 | 2022;13 | 16.56 | 19.67 | EPI_ISL_11889673 |
| CCF-0260 | 2022;13 | 17.08 | 17.43 | EPI_ISL_11889674 |
| CCF-0258 | 2022;13 | 23.01 | 23.74 | EPI_ISL_11889675 |
| CCF-0255 | 2022;13 | 20.1 | 20.15 | EPI_ISL_11889676 |
| CCF-0256 | 2022;13 | 18.1 | 18.34 | EPI_ISL_11889677 |
| CCF-0257 | 2022;13 | 20 | 24.01 | EPI_ISL_11889678 |
| CCF-0250 | 2022;13 | 23.5 | 28.08 | EPI_ISL_11889679 |
| CCF-0251 | 2022;13 | 20.02 | 19.94 | EPI_ISL_11889680 |
| CCF-0252 | 2022;13 | 21.72 | 22.2 | EPI_ISL_11889681 |
| CCF-0253 | 2022;13 | 17.11 | 17.26 | EPI_ISL_11889682 |
| CCF-0254 | 2022;13 | 26.44 | 31.02 | EPI_ISL_11889683 |
| CCF-0248 | 2022;13 | 18.25 | 23.24 | EPI_ISL_11889684 |
| CCF-0244 | 2022;13 | 24.81 | 25.27 | EPI_ISL_11889685 |
| CCF-0245 | 2022;13 | 21.34 | 21.46 | EPI_ISL_11889686 |
| CCF-0246 | 2022;13 | 26.39 | 26.98 | EPI_ISL_11889687 |
| CCF-0247 | 2022;13 | 21.84 | 21.71 | EPI_ISL_11889688 |
| CCF-0239 | 2022;13 | 26.13 | 26.53 | EPI_ISL_11889689 |
| CCF-0241 | 2022;13 | 16.36 | 16.35 | EPI_ISL_11889690 |
| CCF-0242 | 2022;13 | 20.37 | 21.36 | EPI_ISL_11889691 |
| CCF-0243 | 2022;13 | 19.67 | 19.72 | EPI_ISL_11889692 |
| CCF-0233 | 2022;13 | 22.04 | 26.38 | EPI_ISL_11889693 |
| CCF-0234 | 2022;13 | 18.43 | 19.19 | EPI_ISL_11889694 |
| CCF-0236 | 2022;13 | 17.34 | 17.37 | EPI_ISL_11889695 |
| CCF-0231 | 2022;13 | 23.93 | 28.81 | EPI_ISL_11889696 |
| CCF-0232 | 2022;13 | 18.13 | 18.23 | EPI_ISL_11889697 |
| CCF-0220 | 2022;13 | 18.38 | 18.15 | EPI_ISL_11889698 |
| CCF-0221 | 2022;13 | 18.12 | 17.94 | EPI_ISL_11889699 |
| CCF-0223 | 2022;13 | 20.29 | 20.14 | EPI_ISL_11889700 |
| CCF-0224 | 2022;13 | 26.27 | 26.22 | EPI_ISL_11889701 |
| CCF-0225 | 2022;13 | 23.55 | 23.39 | EPI_ISL_11889702 |
| CCF-0226 | 2022;13 | 23.7 | 28.31 | EPI_ISL_11889703 |
| CCF-0228 | 2022;13 | 22.19 | 22.23 | EPI_ISL_11889704 |
| CCF-0218 | 2022;13 | 27.07 | 26.82 | EPI_ISL_11889706 |
| CCF-0219 | 2022;13 | 17.65 | 22.43 | EPI_ISL_11889707 |
| CCF-0215 | 2022;13 | 18.1 | 18.35 | EPI_ISL_11889708 |
| CCF-0212 | 2022;13 | 27.96 | 27.89 | EPI_ISL_11889709 |
| CCF-0209 | 2022;13 | 19.28 | 19.37 | EPI_ISL_11889710 |
| CCF-0207 | 2022;13 | 28.08 | 28.11 | EPI_ISL_11889711 |
| CCF-0210 | 2022;13 | 28.08 | 28.11 | EPI_ISL_11889711 |
| CCF-0719 | 2022;13 | 26.88 | 26.34 | EPI_ISL_11889712 |
| CCF-0720 | 2022;13 | 23 | 22.64 | EPI_ISL_11889713 |
| CCF-0746 | 2022;13 | 22.17 | 21.62 | EPI_ISL_11889714 |
| CCF-0722 | 2022;13 | 15.47 | 15.25 | EPI_ISL_11889715 |

| | | | | |
|---|---|---|---|---|
| CCF-0723 | 2022;13 | 20.06 | 19.75 | EPI_ISL_11889716 |
| CCF-0713 | 2022;13 | 23.54 | 23.17 | EPI_ISL_11889717 |
| CCF-0714 | 2022;13 | 17.62 | 17.6 | EPI_ISL_11889718 |
| CCF-0715 | 2022;13 | 18.22 | 17.88 | EPI_ISL_11889719 |
| CCF-0725 | 2022;13 | 18.28 | 17.8 | EPI_ISL_11889721 |
| CCF-0733 | 2022;13 | 21.44 | 24.74 | EPI_ISL_11889722 |
| CCF-0730 | 2022;13 | 19.68 | 19.46 | EPI_ISL_11889723 |
| CCF-0735 | 2022;13 | 18.53 | 18.4 | EPI_ISL_11889724 |
| CCF-0736 | 2022;13 | 17.74 | 17.68 | EPI_ISL_11889725 |
| CCF-0737 | 2022;13 | 18.75 | 18.55 | EPI_ISL_11889726 |
| CCF-0738 | 2022;13 | 17.38 | 17.16 | EPI_ISL_11889727 |
| CCF-0739 | 2022;13 | 20.76 | 20.36 | EPI_ISL_11889728 |
| CCF-0747 | 2022;13 | 20.15 | 19.8 | EPI_ISL_11889729 |
| CCF-0741 | 2022;13 | 26.69 | 26.11 | EPI_ISL_11889730 |
| CCF-0743 | 2022;13 | 24.48 | 24.19 | EPI_ISL_11889731 |
| CCF-0771 | 2022;12 | 16.05 | 15.58 | EPI_ISL_11889732 |
| CCF-0230 | 2022;13 | 18.69 | 18.77 | EPI_ISL_11889736 |
| CCF-0727 | 2022;13 | 24.24 | 24.04 | EPI_ISL_11889739 |
| CCF-0732 | 2022;13 | 19.48 | 19.57 | EPI_ISL_11889740 |
| CCF-0742 | 2022;13 | 19.04 | 18.75 | EPI_ISL_11889741 |
| CCF-0745 | 2022;13 | 23.94 | 23.56 | EPI_ISL_11889742 |
| Penn1 | 2022;10 | 24.3 | 23.8 | EPI_ISL_12001561 |
| Penn2 | 2022;10 | 23 | 22.6 | EPI_ISL_12001562 |
| Penn4 | 2022;10 | 27.2 | 26.9 | EPI_ISL_12001563 |
| Penn5 | 2022;10 | 26.2 | 25.9 | EPI_ISL_12001564 |
| Penn8 | 2022;10 | 20.3 | 20 | EPI_ISL_12001565 |
| Penn9 | 2022;10 | 18.6 | 18.3 | EPI_ISL_12001566 |
| Penn10 | 2022;10 | 22.7 | 22.4 | EPI_ISL_12001567 |
| Penn12 | 2022;10 | 25.3 | 25 | EPI_ISL_12001568 |
| Penn18 | 2022;10 | 21 | 21.2 | EPI_ISL_12001575 |
| Penn20 | 2022;10 | 20.3 | 20 | EPI_ISL_12001576 |
| Penn21 | 2022;10 | 24.1 | 23.8 | EPI_ISL_12001578 |
| Penn22 | 2022;10 | 17.9 | 17.7 | EPI_ISL_12001580 |
| Penn23 | 2022;10 | 22.1 | 21.8 | EPI_ISL_12001581 |
| Penn31 | 2022;10 | 25.3 | 25.5 | EPI_ISL_12001583 |
| Penn29 | 2022;10 | 20 | 20.1 | EPI_ISL_12001584 |
| Penn32 | 2022;10 | 24.2 | 23.7 | EPI_ISL_12001585 |
| Penn34 | 2022;10 | 19.9 | 19.7 | EPI_ISL_12001586 |
| Penn38 | 2022;11 | 21.8 | 21.4 | EPI_ISL_12001592 |
| Penn39 | 2022;11 | 26.3 | 26 | EPI_ISL_12001606 |
| Penn40 | 2022;11 | 21.9 | 22.1 | EPI_ISL_12001609 |
| Penn46 | 2022;11 | 21.2 | 20.9 | EPI_ISL_12001636 |
| Penn47 | 2022;11 | 23.1 | 23.2 | EPI_ISL_12001637 |

| | | | | |
|---|---|---|---|---|
| Penn48 | 2022;11 | 18.8 | 18.4 | EPI_ISL_12001638 |
| Penn51 | 2022;11 | 19.8 | 20 | EPI_ISL_12001646 |
| Penn53 | 2022;11 | 19.2 | 18.9 | EPI_ISL_12001647 |
| Penn56 | 2022;11 | 27.4 | 27.1 | EPI_ISL_12001653 |
| Penn57 | 2022;11 | 22.4 | 22.2 | EPI_ISL_12001654 |
| Penn62 | 2022;11 | 16.4 | 16.4 | EPI_ISL_12001656 |
| Penn63 | 2022;11 | 20.8 | 20.3 | EPI_ISL_12001657 |
| Penn65 | 2022;11 | 27.5 | 26.8 | EPI_ISL_12001658 |
| Penn68 | 2022;11 | 25.7 | 25.6 | EPI_ISL_12001659 |
| Penn71 | 2022;12 | 15 | 14.8 | EPI_ISL_12001660 |
| Penn70 | 2022;12 | 21.5 | 22 | EPI_ISL_12001661 |
| Penn72 | 2022;12 | 22.54 | 25.53 | EPI_ISL_12001666 |
| Penn73 | 2022;12 | 18.68 | 22.15 | EPI_ISL_12001670 |
| Penn76 | 2022;12 | 22.4 | 22 | EPI_ISL_12001673 |
| Penn79 | 2022;12 | 18.68 | 22.19 | EPI_ISL_12001677 |
| Penn80 | 2022;12 | 23.3 | 23.2 | EPI_ISL_12001682 |
| Penn82 | 2022;12 | 17.9 | 17.8 | EPI_ISL_12001684 |
| Penn84 | 2022;12 | 19.45 | 23.22 | EPI_ISL_12001691 |
| Penn85 | 2022;12 | 22.9 | 23 | EPI_ISL_12001695 |
| Penn94 | 2022;12 | 26.7 | 26.3 | EPI_ISL_12001703 |
| YNHH-0346 | 2022;12 | 28.9 | 28.6 | EPI_ISL_12006439 |
| YNHH-0363 | 2022;12 | 27.1 | 26.9 | EPI_ISL_12006440 |
| YNHH-0365 | 2022;12 | 21.6 | 21.5 | EPI_ISL_12006441 |
| YNHH-0375 | 2022;12 | 19.5 | 19.2 | EPI_ISL_12006442 |
| YNHH-0385 | 2022;12 | 24 | 23.8 | EPI_ISL_12006443 |
| YNHH-0453 | 2022;13 | 21.5 | 21.1 | EPI_ISL_12006444 |
| YNHH-0343 | 2022;12 | 20.6 | 20.1 | EPI_ISL_12006447 |
| YNHH-0351 | 2022;12 | 30.5 | 30 | EPI_ISL_12006448 |
| YNHH-0373 | 2022;12 | 30.2 | 29.5 | EPI_ISL_12006449 |
| YNHH-0380 | 2022;12 | 25.5 | 25.4 | EPI_ISL_12006450 |
| YNHH-0417 | 2022;12 | 21.8 | 21.5 | EPI_ISL_12006451 |
| YNHH-0445 | 2022;13 | 23.3 | 23.3 | EPI_ISL_12006452 |
| YNHH-0347 | 2022;12 | 23 | 22.8 | EPI_ISL_12006454 |
| YNHH-0420 | 2022;12 | 24.8 | 24.6 | EPI_ISL_12006457 |
| YNHH-0443 | 2022;13 | 26.6 | 26.4 | EPI_ISL_12006458 |
| YNHH-0427 | 2022;12 | 26.9 | 26.7 | EPI_ISL_12006459 |
| YNHH-0398 | 2022;12 | 19.1 | 19 | EPI_ISL_12006460 |
| YNHH-0404 | 2022;12 | 22.6 | 22 | EPI_ISL_12006465 |
| YNHH-0440 | 2022;13 | 26.8 | 26.6 | EPI_ISL_12006466 |
| YNHH-0424 | 2022;12 | 21.4 | 21.1 | EPI_ISL_12006467 |
| YNHH-0411 | 2022;12 | 29.3 | 28.9 | EPI_ISL_12006468 |
| YNHH-0364 | 2022;12 | 17.9 | 17.9 | EPI_ISL_12006472 |
| YNHH-0402 | 2022;12 | 22.3 | 22.1 | EPI_ISL_12006473 |

| | | | | |
|---|---|---|---|---|
| YNHH-0377 | 2022;12 | 30.2 | 29.8 | EPI_ISL_12006474 |
| YNHH-0456 | 2022;13 | 22.9 | 22.5 | EPI_ISL_12006475 |
| YNHH-0442 | 2022;13 | 27.2 | 27.2 | EPI_ISL_12006476 |
| YNHH-0361 | 2022;12 | 28.5 | 28.4 | EPI_ISL_12006478 |
| YNHH-0376 | 2022;12 | 25.3 | 25.3 | EPI_ISL_12006481 |
| YNHH-0409 | 2022;12 | 27.1 | 26.5 | EPI_ISL_12006482 |
| YNHH-0437 | 2022;13 | 22.1 | 21.4 | EPI_ISL_12006483 |
| YNHH-0450 | 2022;13 | 25.7 | 25.5 | EPI_ISL_12006484 |
| YNHH-0407 | 2022;12 | 22.5 | 22.3 | EPI_ISL_12006489 |
| YNHH-0382 | 2022;12 | 23.2 | 23.5 | EPI_ISL_12006490 |
| YNHH-0452 | 2022;13 | 20.3 | 19.9 | EPI_ISL_12006491 |
| YNHH-0369 | 2022;12 | 23.5 | 23.4 | EPI_ISL_12006492 |
| YNHH-0384 | 2022;12 | 21.4 | 21.2 | EPI_ISL_12006497 |
| YNHH-0422 | 2022;12 | 23.5 | 23.1 | EPI_ISL_12006498 |
| YNHH-0416 | 2022;12 | 29.7 | 29.5 | EPI_ISL_12006499 |
| YNHH-0408 | 2022;12 | 27.4 | 26.8 | EPI_ISL_12006500 |
| YNHH-0447 | 2022;13 | 19 | 18.8 | EPI_ISL_12006505 |
| YNHH-0400 | 2022;12 | 22.8 | 22.4 | EPI_ISL_12006506 |
| YNHH-0439 | 2022;13 | 26 | 25.6 | EPI_ISL_12006507 |
| YNHH-0448 | 2022;13 | 28.8 | 28.3 | EPI_ISL_12006508 |
| YNHH-0457 | 2022;13 | 19 | 18.5 | EPI_ISL_12006513 |
| YNHH-0403 | 2022;12 | 26.6 | 25.9 | EPI_ISL_12006514 |
| YNHH-0446 | 2022;13 | 29.8 | 29.2 | EPI_ISL_12006515 |
| YNHH-0397 | 2022;12 | 18.8 | 18.5 | EPI_ISL_12006516 |
| YNHH-0360 | 2022;12 | 23.1 | 22.4 | EPI_ISL_12006517 |
| YNHH-0428 | 2022;12 | 21.2 | 21.1 | EPI_ISL_12006521 |
| YNHH-0414 | 2022;12 | 25.6 | 25.2 | EPI_ISL_12006522 |
| YNHH-0454 | 2022;13 | 20 | 19.8 | EPI_ISL_12006523 |
| YNHH-0371 | 2022;12 | 18.8 | 18.5 | EPI_ISL_12006524 |
| YNHH-0357 | 2022;12 | 23.1 | 23 | EPI_ISL_12006526 |
| YNHH-0362 | 2022;12 | 30.5 | 30.3 | EPI_ISL_12006528 |
| YNHH-0429 | 2022;12 | 29.6 | 29.1 | EPI_ISL_12006529 |
| YNHH-0378 | 2022;12 | 22.8 | 22.4 | EPI_ISL_12006531 |
| YNHH-0496 | 2022;13 | 20.5 | 20.2 | EPI_ISL_12006532 |
| YNHH-0461 | 2022;13 | 16.5 | 20.9 | EPI_ISL_12006534 |
| YNHH-0467 | 2022;13 | 22.5 | 22 | EPI_ISL_12006535 |
| YNHH-0481 | 2022;13 | 29.5 | 28.9 | EPI_ISL_12006536 |
| YNHH-0379 | 2022;12 | 22.7 | 22.6 | EPI_ISL_12006538 |
| YNHH-0389 | 2022;12 | 21.1 | 20.7 | EPI_ISL_12006539 |
| YNHH-0489 | 2022;13 | 16.6 | 16.4 | EPI_ISL_12006540 |
| YNHH-0470 | 2022;13 | 30.8 | 30.4 | EPI_ISL_12006541 |
| YNHH-0459 | 2022;13 | 23.7 | 23.4 | EPI_ISL_12006542 |
| YNHH-0482 | 2022;13 | 22.7 | 22.7 | EPI_ISL_12006543 |

| | | | | |
|---|---|---|---|---|
| YNHH-0488 | 2022;13 | 25.9 | 25.3 | EPI_ISL_12006544 |
| YNHH-0390 | 2022;12 | 26.8 | 26.5 | EPI_ISL_12006547 |
| YNHH-0490 | 2022;13 | 19 | 18.7 | EPI_ISL_12006550 |
| YNHH-0487 | 2022;13 | 20.5 | 20.4 | EPI_ISL_12006551 |
| YNHH-0480 | 2022;13 | 29.3 | 28.9 | EPI_ISL_12006552 |
| YNHH-0388 | 2022;12 | 18.1 | 18 | EPI_ISL_12006554 |
| YNHH-0477 | 2022;13 | 18.7 | 18.4 | EPI_ISL_12006555 |
| YNHH-0464 | 2022;13 | 22.1 | 22.3 | EPI_ISL_12006556 |
| YNHH-0502 | 2022;13 | 19 | 18.7 | EPI_ISL_12006557 |
| YNHH-0463 | 2022;13 | 29.5 | 28.9 | EPI_ISL_12006558 |
| YNHH-0374 | 2022;12 | 21.7 | 21.6 | EPI_ISL_12006562 |
| YNHH-0466 | 2022;13 | 21.2 | 20.6 | EPI_ISL_12006563 |
| YNHH-0468 | 2022;13 | 29.1 | 28.6 | EPI_ISL_12006565 |
| YNHH-0476 | 2022;13 | 22.5 | 21.8 | EPI_ISL_12006566 |
| YNHH-0386 | 2022;12 | 22.4 | 22.1 | EPI_ISL_12006570 |
| YNHH-0465 | 2022;13 | 23.1 | 22.8 | EPI_ISL_12006573 |
| YNHH-0522 | 2022;13 | 19 | 18.6 | EPI_ISL_12006574 |
| YNHH-0518 | 2022;13 | 19.2 | 19.2 | EPI_ISL_12006575 |
| YNHH-0393 | 2022;12 | 26.8 | 26.5 | EPI_ISL_12006578 |
| YNHH-0517 | 2022;13 | 23.3 | 23 | EPI_ISL_12006580 |
| YNHH-0494 | 2022;13 | 24.2 | 23.8 | EPI_ISL_12006581 |
| YNHH-0491 | 2022;13 | 29.6 | 29 | EPI_ISL_12006582 |
| YNHH-0545 | 2022;13 | 19.9 | 23.6 | EPI_ISL_12006584 |
| YNHH-0399 | 2022;12 | 29.1 | 28.6 | EPI_ISL_12006585 |
| YNHH-0391 | 2022;12 | 22.8 | 22.6 | EPI_ISL_12006586 |
| YNHH-0432 | 2022;13 | 24.5 | 24.2 | EPI_ISL_12006587 |
| YNHH-0519 | 2022;13 | 29 | 28.6 | EPI_ISL_12006588 |
| YNHH-0484 | 2022;13 | 24.7 | 24.4 | EPI_ISL_12006589 |
| YNHH-0516 | 2022;13 | 26.8 | 26.5 | EPI_ISL_12006591 |
| YNHH-0381 | 2022;12 | 28.9 | 28.2 | EPI_ISL_12006592 |
| YNHH-0505 | 2022;13 | 20.8 | 20.5 | EPI_ISL_12006595 |
| YNHH-0458 | 2022;13 | 27.6 | 26.9 | EPI_ISL_12006596 |
| YNHH-0483 | 2022;13 | 29 | 28.9 | EPI_ISL_12006597 |
| YNHH-0383 | 2022;12 | 27.4 | 27.1 | EPI_ISL_12006600 |
| YNHH-0471 | 2022;13 | 23.2 | 22.6 | EPI_ISL_12006602 |
| YNHH-0475 | 2022;13 | 19.4 | 19.2 | EPI_ISL_12006603 |
| YNHH-0492 | 2022;13 | 22.8 | 22.5 | EPI_ISL_12006604 |
| YNHH-0526 | 2022;13 | 19.8 | 19.7 | EPI_ISL_12006605 |
| YNHH-0534 | 2022;13 | 22.4 | 22 | EPI_ISL_12006606 |
| YNHH-0550 | 2022;13 | 21.9 | 21.4 | EPI_ISL_12006607 |
| YNHH-0418 | 2022;12 | 19.5 | 19.3 | EPI_ISL_12006608 |
| YNHH-0493 | 2022;13 | 22.8 | 22.3 | EPI_ISL_12006609 |
| YNHH-0503 | 2022;13 | 19.3 | 18.8 | EPI_ISL_12006611 |

| | | | | |
|---|---|---|---|---|
| YNHH-0485 | 2022;13 | 25.4 | 25.2 | EPI_ISL_12006612 |
| YNHH-0500 | 2022;13 | 25.2 | 24.8 | EPI_ISL_12006613 |
| YNHH-0552 | 2022;13 | 20.8 | 20.5 | EPI_ISL_12006614 |
| YNHH-0524 | 2022;13 | 29.5 | 29 | EPI_ISL_12006615 |
| YNHH-0425 | 2022;12 | 17.7 | 17.5 | EPI_ISL_12006616 |
| YNHH-0472 | 2022;13 | 30.8 | 30.2 | EPI_ISL_12006618 |
| YNHH-0474 | 2022;13 | 23 | 22.4 | EPI_ISL_12006619 |
| YNHH-0460 | 2022;13 | 23 | 22.3 | EPI_ISL_12006620 |
| CCF-0684 | 2022;14 | 19.11 | 19.03 | EPI_ISL_12079922 |
| CCF-0335 | 2022;14 | 19.47 | 19.59 | EPI_ISL_12079923 |
| CCF-0697 | 2022;14 | 18.26 | 17.72 | EPI_ISL_12079924 |
| CCF-0315 | 2022;14 | 16.7 | 16.62 | EPI_ISL_12079925 |
| CCF-0282 | 2022;14 | 18.24 | 22.79 | EPI_ISL_12079926 |
| CCF-0288 | 2022;14 | 18.01 | 18.05 | EPI_ISL_12079927 |
| CCF-0292 | 2022;14 | 19.61 | 20.24 | EPI_ISL_12079928 |
| CCF-0294 | 2022;14 | 21.6 | 21.88 | EPI_ISL_12079929 |
| CCF-0700 | 2022;14 | 22.14 | 21.66 | EPI_ISL_12079930 |
| CCF-0693 | 2022;14 | 19.26 | 18.74 | EPI_ISL_12079931 |
| CCF-0695 | 2022;14 | 20.91 | 20.62 | EPI_ISL_12079932 |
| CCF-0691 | 2022;14 | 21.25 | 20.88 | EPI_ISL_12079933 |
| CCF-0692 | 2022;14 | 21.55 | 21.2 | EPI_ISL_12079934 |
| CCF-0678 | 2022;14 | 16.31 | 16.11 | EPI_ISL_12079935 |
| CCF-0680 | 2022;14 | 21.48 | 21.23 | EPI_ISL_12079936 |
| CCF-0682 | 2022;14 | 18.91 | 18.68 | EPI_ISL_12079937 |
| CCF-0675 | 2022;14 | 22.9 | 22.43 | EPI_ISL_12079938 |
| CCF-0306 | 2022;14 | 15.57 | 15.48 | EPI_ISL_12079939 |
| CCF-0307 | 2022;14 | 18.52 | 23.11 | EPI_ISL_12079940 |
| CCF-0309 | 2022;14 | 20.69 | 25.79 | EPI_ISL_12079941 |
| CCF-0672 | 2022;14 | 18.44 | 18.2 | EPI_ISL_12079942 |
| CCF-0671 | 2022;14 | 17.38 | 17.16 | EPI_ISL_12079943 |
| CCF-0319 | 2022;14 | 17.58 | 17.94 | EPI_ISL_12079948 |
| CCF-0322 | 2022;14 | 18.52 | 18.81 | EPI_ISL_12079949 |
| CCF-0324 | 2022;14 | 17.15 | 17.53 | EPI_ISL_12079950 |
| CCF-0666 | 2022;14 | 20.71 | 20.57 | EPI_ISL_12079951 |
| CCF-0668 | 2022;14 | 15.88 | 15.56 | EPI_ISL_12079952 |
| CCF-0326 | 2022;14 | 21.62 | 22.44 | EPI_ISL_12079953 |
| CCF-0330 | 2022;14 | 14.91 | 14.98 | EPI_ISL_12079954 |
| CCF-0334 | 2022;14 | 16.6 | 16.79 | EPI_ISL_12079955 |
| CCF-0341 | 2022;14 | 18.12 | 18.26 | EPI_ISL_12079956 |
| CCF-0661 | 2022;14 | 17.43 | 17.28 | EPI_ISL_12079957 |
| CCF-0662 | 2022;14 | 19.07 | 18.78 | EPI_ISL_12079958 |
| CCF-0346 | 2022;14 | 18.57 | 18.67 | EPI_ISL_12079959 |
| CCF-0373 | 2022;15 | 21.25 | 26.46 | EPI_ISL_12079960 |

| | | | | |
|---|---|---|---|---|
| CCF-0374 | 2022;15 | 18.61 | 18.6 | EPI_ISL_12079961 |
| CCF-0375 | 2022;15 | 16.27 | 16.49 | EPI_ISL_12079962 |
| CCF-0376 | 2022;15 | 19.9 | 20.05 | EPI_ISL_12079963 |
| CCF-0377 | 2022;15 | 21.5 | 26.53 | EPI_ISL_12079964 |
| CCF-0378 | 2022;15 | 19.44 | 19.61 | EPI_ISL_12079965 |
| CCF-0379 | 2022;15 | 16.17 | 20.88 | EPI_ISL_12079966 |
| CCF-0380 | 2022;15 | 18.33 | 18.57 | EPI_ISL_12079967 |
| CCF-0381 | 2022;15 | 17.03 | 17.11 | EPI_ISL_12079968 |
| CCF-0361 | 2022;14 | 16.22 | 21.06 | EPI_ISL_12079969 |
| CCF-0643 | 2022;15 | 18.44 | 18.31 | EPI_ISL_12079970 |
| CCF-0649 | 2022;15 | 19.44 | 19.1 | EPI_ISL_12079971 |
| CCF-0650 | 2022;14 | 16.49 | 16.37 | EPI_ISL_12079972 |
| CCF-0323 | 2022;14 | 17.62 | 18.04 | EPI_ISL_12079983 |
| CCF-0368 | 2022;15 | 18.13 | 17.89 | EPI_ISL_12079984 |
| CCF-0296 | 2022;14 | 16.48 | 21.51 | EPI_ISL_12079985 |
| CCF-0298 | 2022;14 | 16.48 | 21.51 | EPI_ISL_12079985 |
| CCF-0702 | 2022;14 | 15.45 | 15.26 | EPI_ISL_12079986 |
| CCF-0384 | 2022;15 | 20.15 | 20.25 | EPI_ISL_12080000 |
| CCF-0385 | 2022;15 | 22.99 | 28.62 | EPI_ISL_12080001 |
| CCF-0331 | 2022;14 | 18.82 | 19.06 | EPI_ISL_12080002 |
| CCF-0677 | 2022;14 | 23.6 | 23.57 | EPI_ISL_12080003 |
| CCF-0348 | 2022;14 | 21.06 | 21.55 | EPI_ISL_12080004 |
| CCF-0333 | 2022;14 | 22.26 | 22.5 | EPI_ISL_12080005 |
| CCF-0328 | 2022;14 | 21.14 | 21.33 | EPI_ISL_12080006 |
| CCF-0686 | 2022;14 | 21.09 | 20.85 | EPI_ISL_12080008 |
| CCF-0311 | 2022;14 | 22.83 | 26.99 | EPI_ISL_12080009 |
| CCF-0354 | 2022;14 | 21.87 | 25.78 | EPI_ISL_12080010 |
| CCF-0318 | 2022;14 | 19.83 | 19.99 | EPI_ISL_12080011 |
| CCF-0313 | 2022;14 | 18.27 | 18.72 | EPI_ISL_12080014 |
| CCF-0339 | 2022;14 | 23.39 | 23.76 | EPI_ISL_12080015 |
| CCF-0640 | 2022;15 | 24.2 | 27.88 | EPI_ISL_12080031 |
| YNHH-0536 | 2022;13 | 22 | 21.6 | EPI_ISL_12109111 |
| YNHH-0575 | 2022;13 | 20 | 19.9 | EPI_ISL_12109112 |
| YNHH-0537 | 2022;13 | 19.7 | 19.3 | EPI_ISL_12109113 |
| YNHH-0543 | 2022;13 | 18.8 | 18.4 | EPI_ISL_12109114 |
| YNHH-0585 | 2022;13 | 18.3 | 18.3 | EPI_ISL_12109115 |
| YNHH-0812 | 2022;14 | 24 | 23.7 | EPI_ISL_12109116 |
| YNHH-0565 | 2022;13 | 20.8 | 20.2 | EPI_ISL_12109118 |
| YNHH-0589 | 2022;13 | 30.9 | 30.5 | EPI_ISL_12109119 |
| YNHH-0611 | 2022;13 | 25.6 | 25.2 | EPI_ISL_12109120 |
| YNHH-0626 | 2022;13 | 24.5 | 24.3 | EPI_ISL_12109121 |
| YNHH-0629 | 2022;13 | 25.3 | 28.5 | EPI_ISL_12109122 |
| YNHH-0714 | 2022;13 | 18.9 | 18.7 | EPI_ISL_12109123 |

| | | | | |
|---|---|---|---|---|
| YNHH-0563 | 2022;13 | 21.4 | 21.2 | EPI_ISL_12109125 |
| YNHH-0593 | 2022;13 | 25.2 | 24.9 | EPI_ISL_12109126 |
| YNHH-0649 | 2022;13 | 24.4 | 24.3 | EPI_ISL_12109127 |
| YNHH-0638 | 2022;13 | 19.5 | 19.2 | EPI_ISL_12109128 |
| YNHH-0734 | 2022;13 | 18.8 | 18.5 | EPI_ISL_12109131 |
| YNHH-0590 | 2022;13 | 19.3 | 22.6 | EPI_ISL_12109133 |
| YNHH-0557 | 2022;13 | 23.9 | 23.5 | EPI_ISL_12109134 |
| YNHH-0586 | 2022;13 | 22.2 | 21.9 | EPI_ISL_12109135 |
| YNHH-0740 | 2022;13 | 18.7 | 18.7 | EPI_ISL_12109136 |
| YNHH-0579 | 2022;13 | 17.8 | 17.4 | EPI_ISL_12109139 |
| YNHH-0608 | 2022;13 | 18.5 | 18.4 | EPI_ISL_12109140 |
| YNHH-0744 | 2022;13 | 25.8 | 25.6 | EPI_ISL_12109141 |
| YNHH-0747 | 2022;13 | 23.7 | 23.3 | EPI_ISL_12109144 |
| YNHH-0648 | 2022;13 | 20.6 | 25.2 | EPI_ISL_12109148 |
| YNHH-0772 | 2022;14 | 20.6 | 24.6 | EPI_ISL_12109149 |
| YNHH-0644 | 2022;13 | 17.9 | 17.6 | EPI_ISL_12109150 |
| YNHH-0665 | 2022;13 | 25 | 24.6 | EPI_ISL_12109151 |
| YNHH-0588 | 2022;13 | 18.8 | 18.7 | EPI_ISL_12109152 |
| YNHH-0724 | 2022;13 | 16.8 | 16.8 | EPI_ISL_12109153 |
| YNHH-0600 | 2022;13 | 20.3 | 20.1 | EPI_ISL_12109156 |
| YNHH-0760 | 2022;14 | 16.2 | 15.8 | EPI_ISL_12109157 |
| YNHH-0583 | 2022;13 | 26.5 | 26.2 | EPI_ISL_12109158 |
| YNHH-0604 | 2022;13 | 17.4 | 17.4 | EPI_ISL_12109159 |
| YNHH-0630 | 2022;13 | 17.3 | 17.6 | EPI_ISL_12109160 |
| YNHH-0640 | 2022;13 | 20.6 | 20.2 | EPI_ISL_12109161 |
| YNHH-0716 | 2022;13 | 15.8 | 15.7 | EPI_ISL_12109162 |
| YNHH-0718 | 2022;13 | 18.3 | 18.1 | EPI_ISL_12109163 |
| YNHH-0556 | 2022;13 | 17.8 | 17.6 | EPI_ISL_12109164 |
| YNHH-0627 | 2022;13 | 21.1 | 21 | EPI_ISL_12109165 |
| YNHH-0634 | 2022;13 | 21 | 20.9 | EPI_ISL_12109166 |
| YNHH-0774 | 2022;14 | 14.8 | 15.3 | EPI_ISL_12109167 |
| YNHH-0622 | 2022;13 | 20.7 | 20.2 | EPI_ISL_12109168 |
| YNHH-0717 | 2022;13 | 22.4 | 22.1 | EPI_ISL_12109170 |
| YNHH-0646 | 2022;13 | 20.4 | 20.2 | EPI_ISL_12109171 |
| YNHH-0681 | 2022;13 | 26 | 25.9 | EPI_ISL_12109172 |
| YNHH-0659 | 2022;13 | 23.9 | 23.7 | EPI_ISL_12109173 |
| YNHH-0840 | 2022;14 | 18.3 | 22.4 | EPI_ISL_12109175 |
| YNHH-0731 | 2022;13 | 23.1 | 23 | EPI_ISL_12109176 |
| YNHH-0713 | 2022;13 | 24.2 | 23.8 | EPI_ISL_12109177 |
| YNHH-0689 | 2022;13 | 19.8 | 19.3 | EPI_ISL_12109178 |
| YNHH-0706 | 2022;13 | 27.4 | 27.1 | EPI_ISL_12109179 |
| YNHH-0653 | 2022;13 | 19.1 | 18.8 | EPI_ISL_12109180 |
| YNHH-0736 | 2022;13 | 21.8 | 21.6 | EPI_ISL_12109182 |

| | | | | |
|---|---|---|---|---|
| YNHH-0752 | 2022;14 | 22 | 25.9 | EPI_ISL_12109183 |
| YNHH-0692 | 2022;13 | 27.1 | 26.4 | EPI_ISL_12109184 |
| YNHH-0696 | 2022;13 | 20.4 | 20.1 | EPI_ISL_12109185 |
| YNHH-0845 | 2022;14 | 20.1 | 19.7 | EPI_ISL_12109187 |
| YNHH-0746 | 2022;13 | 25.3 | 25.2 | EPI_ISL_12109188 |
| YNHH-0776 | 2022;14 | 24.7 | 24.4 | EPI_ISL_12109189 |
| YNHH-0688 | 2022;13 | 20.4 | 19.9 | EPI_ISL_12109190 |
| YNHH-0697 | 2022;13 | 17.3 | 17.1 | EPI_ISL_12109191 |
| YNHH-0730 | 2022;13 | 25.3 | 25.1 | EPI_ISL_12109192 |
| YNHH-0687 | 2022;13 | 28 | 27.7 | EPI_ISL_12109193 |
| YNHH-0815 | 2022;14 | 19 | 21.9 | EPI_ISL_12109194 |
| YNHH-0810 | 2022;14 | 22.3 | 22.2 | EPI_ISL_12109195 |
| YNHH-0666 | 2022;13 | 18.5 | 18.6 | EPI_ISL_12109196 |
| YNHH-0754 | 2022;14 | 21.4 | 21.4 | EPI_ISL_12109197 |
| YNHH-0725 | 2022;13 | 21.6 | 21.3 | EPI_ISL_12109198 |
| YNHH-0707 | 2022;13 | 28.5 | 27.8 | EPI_ISL_12109199 |
| YNHH-0742 | 2022;13 | 22.7 | 22.7 | EPI_ISL_12109200 |
| YNHH-0677 | 2022;13 | 20.7 | 20.5 | EPI_ISL_12109201 |
| YNHH-0705 | 2022;13 | 22.6 | 22 | EPI_ISL_12109202 |
| YNHH-0782 | 2022;14 | 18.1 | 18 | EPI_ISL_12109204 |
| YNHH-0745 | 2022;13 | 25.2 | 24.6 | EPI_ISL_12109205 |
| YNHH-0670 | 2022;13 | 22.8 | 22.7 | EPI_ISL_12109206 |
| YNHH-0701 | 2022;13 | 23.1 | 23 | EPI_ISL_12109207 |
| YNHH-0789 | 2022;14 | 18.5 | 18.3 | EPI_ISL_12109209 |
| YNHH-0856 | 2022;14 | 25.5 | 25.1 | EPI_ISL_12109210 |
| YNHH-0700 | 2022;13 | 20.6 | 20.2 | EPI_ISL_12109211 |
| YNHH-0809 | 2022;14 | 24 | 23.7 | EPI_ISL_12109212 |
| YNHH-0726 | 2022;13 | 23.5 | 23.4 | EPI_ISL_12109213 |
| YNHH-0674 | 2022;13 | 20.4 | 20.4 | EPI_ISL_12109214 |
| YNHH-0678 | 2022;13 | 25.5 | 24.9 | EPI_ISL_12109215 |
| YNHH-0735 | 2022;13 | 20 | 20 | EPI_ISL_12109216 |
| YNHH-0880 | 2022;14 | 18.4 | 18.2 | EPI_ISL_12109218 |
| YNHH-0763 | 2022;14 | 18.1 | 18.2 | EPI_ISL_12109219 |
| YNHH-0749 | 2022;14 | 19.8 | 19.4 | EPI_ISL_12109220 |
| YNHH-0657 | 2022;13 | 21.9 | 21.7 | EPI_ISL_12109221 |
| YNHH-0596 | 2022;13 | 19.4 | 19.2 | EPI_ISL_12109222 |
| YNHH-0598 | 2022;13 | 19.9 | 19.7 | EPI_ISL_12109224 |
| YNHH-0636 | 2022;13 | 25.9 | 25.5 | EPI_ISL_12109225 |
| YNHH-0542 | 2022;13 | 30.8 | 29.9 | EPI_ISL_12109227 |
| YNHH-0673 | 2022;13 | 18.4 | 18.3 | EPI_ISL_12109228 |
| YNHH-0691 | 2022;13 | 22.6 | 22.2 | EPI_ISL_12109229 |
| YNHH-0683 | 2022;13 | 22.5 | 22.5 | EPI_ISL_12109231 |
| YNHH-0712 | 2022;13 | 26.5 | 26.2 | EPI_ISL_12109232 |

| | | | | |
|---|---|---|---|---|
| YNHH-0739 | 2022;13 | 20.9 | 20.7 | EPI_ISL_12109234 |
| YNHH-0584 | 2022;13 | 23.1 | 22.9 | EPI_ISL_12109235 |
| YNHH-0684 | 2022;13 | 18.6 | 18.4 | EPI_ISL_12109238 |
| YNHH-0671 | 2022;13 | 27.5 | 27.2 | EPI_ISL_12109239 |
| YNHH-0574 | 2022;13 | 29 | 28.5 | EPI_ISL_12109243 |
| YNHH-0628 | 2022;13 | 22 | 21.8 | EPI_ISL_12109244 |
| YNHH-0624 | 2022;13 | 24.7 | 24.5 | EPI_ISL_12109245 |
| YNHH-0613 | 2022;13 | 25.9 | 25.7 | EPI_ISL_12109247 |
| YNHH-0631 | 2022;13 | 17.6 | 17.6 | EPI_ISL_12109248 |
| YNHH-0639 | 2022;13 | 30 | 29.9 | EPI_ISL_12109249 |
| YNHH-0587 | 2022;13 | 25.3 | 25 | EPI_ISL_12109250 |
| YNHH-0577 | 2022;13 | 29 | 28.4 | EPI_ISL_12109251 |
| YNHH-0618 | 2022;13 | 27.5 | 27 | EPI_ISL_12109252 |
| YNHH-0606 | 2022;13 | 23.4 | 23.2 | EPI_ISL_12109253 |
| YNHH-0582 | 2022;13 | 20.8 | 20.9 | EPI_ISL_12109254 |
| YNHH-0573 | 2022;13 | 29.3 | 28.7 | EPI_ISL_12109255 |
| YNHH-0625 | 2022;13 | 17.8 | 18 | EPI_ISL_12109256 |
| YNHH-0562 | 2022;13 | 30.8 | 29.8 | EPI_ISL_12109258 |
| YNHH-0623 | 2022;13 | 23.1 | 22.8 | EPI_ISL_12109259 |
| YNHH-0612 | 2022;13 | 18.1 | 17.9 | EPI_ISL_12109263 |
| YNHH-0728 | 2022;13 | 21.2 | 20.9 | EPI_ISL_12109264 |
| YNHH-0651 | 2022;13 | 24.8 | 24.5 | EPI_ISL_12109266 |
| YNHH-0719 | 2022;13 | 16.7 | 16.7 | EPI_ISL_12109267 |
| YNHH-0710 | 2022;13 | 20 | 19.8 | EPI_ISL_12109268 |
| YNHH-0836 | 2022;14 | 19.6 | 19.4 | EPI_ISL_12109269 |
| YNHH-0715 | 2022;13 | 22.3 | 21.9 | EPI_ISL_12109270 |
| YNHH-0702 | 2022;13 | 19 | 18.8 | EPI_ISL_12109271 |
| YNHH-0664 | 2022;13 | 19.2 | 18.9 | EPI_ISL_12109272 |
| YNHH-0778 | 2022;14 | 17.5 | 17.2 | EPI_ISL_12109273 |
| YNHH-0876 | 2022;14 | 21.9 | 21.7 | EPI_ISL_12109274 |
| YNHH-0757 | 2022;14 | 20.2 | 24.7 | EPI_ISL_12109275 |
| YNHH-0685 | 2022;13 | 21 | 20.7 | EPI_ISL_12109276 |
| YNHH-0675 | 2022;13 | 21.8 | 21.6 | EPI_ISL_12109277 |
| YNHH-0835 | 2022;14 | 19.9 | 19.9 | EPI_ISL_12109278 |
| YNHH-0872 | 2022;14 | 17.6 | 17.4 | EPI_ISL_12109279 |
| YNHH-0662 | 2022;13 | 22.9 | 22.4 | EPI_ISL_12109280 |
| YNHH-0761 | 2022;14 | 24.2 | 24 | EPI_ISL_12109281 |
| YNHH-0878 | 2022;14 | 18 | 18 | EPI_ISL_12109282 |
| YNHH-0767 | 2022;14 | 17.3 | 17.1 | EPI_ISL_12109283 |
| YNHH-0711 | 2022;13 | 19.4 | 19.3 | EPI_ISL_12109285 |
| YNHH-0768 | 2022;14 | 23.9 | 23.9 | EPI_ISL_12109286 |
| YNHH-0650 | 2022;13 | 16.1 | 20.4 | EPI_ISL_12109287 |
| YNHH-0777 | 2022;14 | 18.2 | 18.1 | EPI_ISL_12109288 |

| | | | | |
|---|---|---|---|---|
| YNHH-0758 | 2022;14 | 24.2 | 24.1 | EPI_ISL_12109289 |
| YNHH-0817 | 2022;14 | 16.6 | 21.9 | EPI_ISL_12109290 |
| YNHH-0720 | 2022;13 | 25.2 | 25.1 | EPI_ISL_12109291 |
| YNHH-0655 | 2022;13 | 20.8 | 20.5 | EPI_ISL_12109292 |
| YNHH-0818 | 2022;14 | 16.7 | 16.7 | EPI_ISL_12109293 |
| YNHH-0766 | 2022;14 | 21.2 | 21 | EPI_ISL_12109294 |
| YNHH-0693 | 2022;13 | 18.9 | 18.7 | EPI_ISL_12109295 |
| MSK 1 | 2022;13 | 17.73 | 21.96 | EPI_ISL_12148421 |
| MSK 2 | 2022;13 | 25.56 | 28.87 | EPI_ISL_12148434 |
| MSK 3 | 2022;13 | 21.39 | 25.75 | EPI_ISL_12148451 |
| MSK 4 | 2022;13 | 24.25 | 28.37 | EPI_ISL_12148453 |
| MSK 5 | 2022;14 | 26.33 | 28.99 | EPI_ISL_12148472 |
| MSK 6 | 2022;14 | 27.51 | 30.91 | EPI_ISL_12148475 |
| MSK 7 | 2022;14 | 20.75 | 24.54 | EPI_ISL_12148497 |
| MSK 8 | 2022;14 | 16.97 | 21.01 | EPI_ISL_12148508 |
| MSK 9 | 2022;14 | 16.55 | 20.86 | EPI_ISL_12148511 |
| MSK 10 | 2022;14 | 24.28 | 27.9 | EPI_ISL_12148513 |
| MSK 11 | 2022;14 | 23.96 | 27.76 | EPI_ISL_12148519 |
| MSK 12 | 2022;14 | 17.63 | 21.97 | EPI_ISL_12148520 |
| MSK 13 | 2022;14 | 17.35 | 21.6 | EPI_ISL_12148521 |
| MSK 14 | 2022;14 | 21.09 | 25.27 | EPI_ISL_12148522 |
| MSK 15 | 2022;14 | 25.08 | 28.62 | EPI_ISL_12148523 |
| MSK 16 | 2022;14 | 20.82 | 25.24 | EPI_ISL_12148525 |
| Columbia 11 | 2022;10 | 25.7 | 25 | EPI_ISL_12191742 |
| Columbia 41 | 2022;11 | 22.1 | 21.6 | EPI_ISL_12191744 |
| Columbia 43 | 2022;12 | 25 | 25 | EPI_ISL_12191745 |
| Columbia 51 | 2022;12 | 30.3 | 30.7 | EPI_ISL_12191746 |
| Columbia 71 | 2022;12 | 26.8 | 26.7 | EPI_ISL_12191747 |
| Columbia 79 | 2022;13 | 18.6 | 18.7 | EPI_ISL_12191750 |
| Columbia 91 | 2022;13 | 19.8 | 19.4 | EPI_ISL_12191752 |
| Columbia 114 | 2022;14 | 22.8 | 22.5 | EPI_ISL_12191760 |
| Columbia 129 | 2022;14 | 27.7 | 27.4 | EPI_ISL_12191761 |
| Columbia 140 | 2022;14 | 18.6 | 21.6 | EPI_ISL_12191762 |
| Columbia 144 | 2022;14 | 24.4 | 23.9 | EPI_ISL_12191764 |
| Columbia 134 | 2022;14 | 21.7 | 21.3 | EPI_ISL_12191766 |
| Columbia 139 | 2022;14 | 19.4 | 19.4 | EPI_ISL_12191767 |
| Columbia 167 | 2022;15 | 15.3 | 15.4 | EPI_ISL_12191770 |
| Columbia 162 | 2022;15 | 25.4 | 25.2 | EPI_ISL_12191772 |
| Columbia 163 | 2022;15 | 20.2 | 19.7 | EPI_ISL_12191774 |
| Columbia 185 | 2022;15 | 28.1 | 27.6 | EPI_ISL_12191775 |
| Columbia 195 | 2022;15 | 32.7 | 33.3 | EPI_ISL_12191778 |
| Columbia 53 | 2022;12 | 26.9 | 27.3 | EPI_ISL_12198483 |
| Columbia 182 | 2022;15 | 30.6 | 34.3 | Pending |

| | | | | |
|---|---|---|---|---|
| Columbia 84 | 2022;13 | 26.7 | 30.4 | Pending |
| Columbia 141 | 2022;14 | 19.6 | 24.3 | Pending |
| Columbia 138 | 2022;14 | 24 | 27.5 | Pending |
| Columbia 165 | 2022;15 | 19.9 | 24 | Pending |
| Columbia 191 | 2022;15 | 18.3 | 22.6 | Pending |
| Columbia 189 | 2022;15 | 26.6 | 30.1 | Pending |
| Columbia 156 | 2022;14 | 24.1 | 28.2 | Pending |
| Columbia 187 | 2022;15 | 19.7 | 24.1 | Pending |
| Columbia 181 | 2022;15 | 16.7 | 20.3 | Pending |
| Columbia 176 | 2022;15 | 26.5 | 30.3 | Pending |
| WCMC-0115 | 2022;14 | 29.1 | 31.9 | Pending |
| WCMC-0116 | 2022;15 | 21.9 | 26 | Pending |
| MSK 37 | 2022;11 | 24.01 | 23.32 | pending |
| MSK 27 | 2022;10 | 30.26 | 29.54 | pending |
| MSK 35 | 2022;11 | 36.27 | 34.56 | pending |
| MSK 38 | 2022;12 | 18.31 | 17.9 | pending |
| MSK 66 | 2022;14 | 19.56 | 19.14 | pending |
| MSK 60 | 2022;14 | 22.09 | 21.84 | pending |
| MSK 28 | 2022;10 | 24.48 | 23.89 | pending |
| MSK 23 | 2022;10 | 25.78 | 25.06 | pending |
| MSK 69 | 2022;14 | 27.16 | 26.67 | pending |
| MSK 30 | 2022;11 | 29.01 | 28.32 | pending |
| MSK 75 | 2022;14 | 29.35 | 27.75 | pending |
| MSK 32 | 2022;11 | 29.88 | 28.46 | pending |
| MSK 31 | 2022;11 | 30.27 | 29.37 | pending |
| MSK 65 | 2022;14 | 30.59 | 26.69 | pending |
| MSK 29 | 2022;11 | 30.71 | 29.74 | pending |
| MSK 64 | 2022;14 | 31.66 | 30.5 | pending |
| MSK 24 | 2022;10 | 32.24 | 31.43 | pending |
| MSK 62 | 2022;14 | 16.47 | 16.22 | pending |
| MSK 67 | 2022;14 | 16.51 | 16.06 | pending |
| MSK 68 | 2022;14 | 18.01 | 17.67 | pending |
| MSK 54 | 2022;13 | 19.05 | 18.56 | pending |
| MSK 45 | 2022;13 | 20.27 | 20.02 | pending |
| MSK 58 | 2022;13 | 20.8 | 22.34 | pending |
| MSK 52 | 2022;13 | 20.96 | 20.46 | pending |
| MSK 40 | 2022;12 | 23.43 | 22.53 | pending |
| MSK 55 | 2022;13 | 23.51 | 22.89 | pending |
| MSK 70 | 2022;14 | 24.12 | 23.58 | pending |
| MSK 59 | 2022;13 | 24.78 | 24.11 | pending |
| MSK 72 | 2022;14 | 25.43 | 25.18 | pending |
| MSK 48 | 2022;13 | 25.67 | 25.35 | pending |
| MSK 51 | 2022;13 | 25.83 | 25.24 | pending |

| | | | | |
|---|---|---|---|---|
| MSK 71 | 2022;14 | 26.64 | 25.85 | pending |
| MSK 49 | 2022;13 | 26.79 | 25.86 | pending |
| MSK 25 | 2022;10 | 27.01 | 28.14 | pending |
| MSK 33 | 2022;11 | 27.51 | 26.8 | pending |
| MSK 39 | 2022;12 | 29.35 | 28.92 | pending |
| MSK 26 | 2022;10 | 29.39 | 28.24 | pending |
| MSK 57 | 2022;13 | 29.49 | 28.36 | pending |
| MSK 34 | 2022;11 | 30.01 | 28.94 | pending |
| MSK 41 | 2022;12 | 32.51 | 30.54 | pending |
| MSK 73 | 2022;14 | 20.59 | 20.51 | pending |
| MSK 43 | 2022;12 | 34.09 | 31.16 | pending |
| MSK 50 | 2022;13 | 17.65 | 17.78 | pending |
| MSK 61 | 2022;14 | 22.35 | 21.66 | pending |
| MSK 97 | 2022;15 | 15.27 | 15.16 | |
| MSK 78 | 2022;15 | 16.27 | 16.02 | |
| MSK 17 | 2022;15 | 16.84 | 20.87 | |
| MSK 20 | 2022;15 | 17.98 | 21.76 | |
| MSK 91 | 2022;15 | 18.19 | 17.83 | |
| MSK 21 | 2022;15 | 18.66 | 23 | |
| MSK 95 | 2022;15 | 19.57 | 19.11 | |
| MSK 19 | 2022;15 | 19.94 | 23.71 | |
| MSK 86 | 2022;15 | 20.65 | 20.24 | |
| MSK 93 | 2022;15 | 21.45 | 21.26 | |
| MSK 18 | 2022;15 | 21.87 | 25.74 | |
| MSK 79 | 2022;15 | 22.18 | 21.76 | |
| MSK 89 | 2022;15 | 23.88 | 27.41 | |
| MSK 88 | 2022;15 | 24.73 | 24.13 | |
| MSK 98 | 2022;15 | 24.98 | 24.34 | |
| MSK 81 | 2022;15 | 25.49 | 25.4 | |
| MSK 94 | 2022;15 | 26.18 | 25.42 | |
| MSK 92 | 2022;15 | 26.67 | 26.06 | |
| MSK 22 | 2022;15 | 26.92 | 29.45 | |
| MSK 87 | 2022;15 | 26.95 | 26.24 | |
| MSK 82 | 2022;15 | 27.94 | 27.01 | |
| MSK 77 | 2022;15 | 28.54 | 27.69 | |
| MSK 85 | 2022;15 | 29.42 | 28.02 | |
| MSK 80 | 2022;15 | 30.03 | 29.25 | |
| MSK 84 | 2022;15 | 30.21 | 28.75 | |
| MSK 96 | 2022;15 | 31.59 | 29.79 | |
| MSK 90 | 2022;15 | 32.98 | 31.12 | |
| MSK 76 | 2022;14 | 33.05 | 31.66 | |
| MSK 83 | 2022;15 | 33.61 | 31.89 | |
| CCF-0564 | 2022;15 | 13.82 | 13.23 | |

| | | | |
|---|---|---|---|
| CCF-0440 | 2022;15 | 13.95 | 14.13 |
| CCF-0598 | 2022;15 | 14.37 | 13.93 |
| CCF-0790 | 2022;12 | 15 | 14.89 |
| CCF-0395 | 2022;15 | 15.08 | 15.21 |
| CCF-0459 | 2022;15 | 15.13 | 15.26 |
| CCF-0488 | 2022;15 | 15.22 | 19.93 |
| CCF-0132 | 2022;11 | 15.36 | 15.24 |
| CCF-0537 | 2022;15 | 15.57 | 16.05 |
| CCF-0586 | 2022;15 | 15.84 | 19.61 |
| CCF-0627 | 2022;15 | 15.91 | 19.9 |
| CCF-0016 | 2022;10 | 16 | 16.27 |
| CCF-0626 | 2022;15 | 16.07 | 15.54 |
| CCF-0582 | 2022;15 | 16.11 | 15.81 |
| CCF-0565 | 2022;15 | 16.32 | 16.28 |
| CCF-0478 | 2022;15 | 16.49 | 16.79 |
| CCF-0618 | 2022;15 | 16.57 | 16.6 |
| CCF-0338 | 2022;14 | 16.67 | 17.06 |
| CCF-0345 | 2022;14 | 16.82 | 16.68 |
| CCF-0059 | 2022;10 | 16.86 | 17.07 |
| CCF-0583 | 2022;15 | 16.9 | 20.49 |
| CCF-0608 | 2022;15 | 16.91 | 16.75 |
| CCF-0528 | 2022;15 | 16.92 | 17.02 |
| CCF-0630 | 2022;15 | 16.93 | 16.88 |
| CCF-0710 | 2022;14 | 16.95 | 16.59 |
| CCF-0472 | 2022;15 | 17 | 17.06 |
| CCF-0426 | 2022;15 | 17.03 | 21.21 |
| CCF-0480 | 2022;15 | 17.05 | 16.93 |
| CCF-0532 | 2022;15 | 17.08 | 17.65 |
| CCF-0637 | 2022;15 | 17.12 | 17.01 |
| CCF-0481 | 2022;15 | 17.21 | 17.35 |
| CCF-0501 | 2022;15 | 17.23 | 17.25 |
| CCF-0572 | 2022;15 | 17.27 | 17.12 |
| CCF-0403 | 2022;15 | 17.28 | 17.66 |
| CCF-0674 | 2022;14 | 17.34 | 17.13 |
| CCF-0538 | 2022;15 | 17.41 | 18.06 |
| CCF-0370 | 2022;15 | 17.49 | 17.69 |
| CCF-0603 | 2022;15 | 17.49 | 21.04 |
| CCF-0512 | 2022;15 | 17.5 | 17.55 |
| CCF-0605 | 2022;15 | 17.51 | 17.26 |
| CCF-0523 | 2022;15 | 17.57 | 22.6 |
| CCF-0628 | 2022;15 | 17.61 | 17.52 |
| CCF-0397 | 2022;15 | 17.74 | 24.05 |
| CCF-0122 | 2022;11 | 17.75 | 17.78 |

| | | | |
|---|---|---|---|
| CCF-0547 | 2022;15 | 17.77 | 22.06 |
| CCF-0508 | 2022;15 | 17.82 | 18.18 |
| CCF-0560 | 2022;15 | 17.82 | 21.67 |
| CCF-0423 | 2022;15 | 17.85 | 17.96 |
| CCF-0555 | 2022;15 | 17.85 | 18.03 |
| CCF-0442 | 2022;15 | 17.86 | 18.06 |
| CCF-0749 | 2022;13 | 17.89 | 17.78 |
| CCF-0625 | 2022;15 | 17.89 | 17.85 |
| CCF-0575 | 2022;15 | 17.9 | 17.92 |
| CCF-0396 | 2022;15 | 17.91 | 18.12 |
| CCF-0452 | 2022;15 | 17.98 | 22.94 |
| CCF-0673 | 2022;14 | 18.06 | 17.76 |
| CCF-0443 | 2022;15 | 18.23 | 18.41 |
| CCF-0550 | 2022;15 | 18.23 | 18.63 |
| CCF-0544 | 2022;15 | 18.25 | 23.75 |
| CCF-0588 | 2022;15 | 18.27 | 18.12 |
| CCF-0138 | 2022;11 | 18.27 | 18.25 |
| CCF-0465 | 2022;15 | 18.34 | 18.56 |
| CCF-0080 | 2022;11 | 18.38 | 18.36 |
| CCF-0493 | 2022;15 | 18.39 | 22.72 |
| CCF-0579 | 2022;15 | 18.45 | 21.97 |
| CCF-0428 | 2022;15 | 18.45 | 23.17 |
| CCF-0278 | 2022;14 | 18.46 | 18.4 |
| CCF-0633 | 2022;15 | 18.47 | 18.35 |
| CCF-0530 | 2022;15 | 18.49 | 18.53 |
| CCF-0448 | 2022;15 | 18.5 | 24.06 |
| CCF-0383 | 2022;15 | 18.53 | 19.14 |
| CCF-0632 | 2022;15 | 18.56 | 18.69 |
| CCF-0519 | 2022;15 | 18.56 | 18.8 |
| CCF-0417 | 2022;15 | 18.56 | 19.08 |
| CCF-0412 | 2022;15 | 18.63 | 22.8 |
| CCF-0513 | 2022;15 | 18.75 | 19.01 |
| CCF-0587 | 2022;15 | 18.76 | 18.6 |
| CCF-0389 | 2022;15 | 18.78 | 18.85 |
| CCF-0491 | 2022;15 | 18.8 | 19.03 |
| CCF-0658 | 2022;14 | 18.83 | 18.53 |
| CCF-0566 | 2022;15 | 18.94 | 18.8 |
| CCF-0552 | 2022;15 | 18.94 | 23.6 |
| CCF-0551 | 2022;15 | 19 | 19.52 |
| CCF-0479 | 2022;15 | 19.03 | 24.13 |
| CCF-0270 | 2022;14 | 19.05 | 19.29 |
| CCF-0489 | 2022;15 | 19.06 | 19.13 |
| CCF-0599 | 2022;15 | 19.1 | 23.11 |

| | | | |
|---|---|---|---|
| CCF-0542 | 2022;15 | 19.13 | 19.92 |
| CCF-0486 | 2022;15 | 19.16 | 19.39 |
| CCF-0517 | 2022;15 | 19.22 | 19.8 |
| CCF-0634 | 2022;15 | 19.23 | 22.42 |
| CCF-0659 | 2022;14 | 19.24 | 19.11 |
| CCF-0462 | 2022;15 | 19.24 | 19.35 |
| CCF-0622 | 2022;15 | 19.27 | 22.51 |
| CCF-0206 | 2022;13 | 19.28 | 19.37 |
| CCF-0439 | 2022;15 | 19.28 | 19.54 |
| CCF-0651 | 2022;14 | 19.31 | 19.13 |
| CCF-0696 | 2022;14 | 19.32 | 19.06 |
| CCF-0610 | 2022;15 | 19.36 | 18.94 |
| CCF-0464 | 2022;15 | 19.4 | 20.31 |
| CCF-0404 | 2022;15 | 19.4 | 23.94 |
| CCF-0485 | 2022;15 | 19.42 | 19.41 |
| CCF-0441 | 2022;15 | 19.45 | 19.51 |
| CCF-0606 | 2022;15 | 19.48 | 23.21 |
| CCF-0470 | 2022;15 | 19.49 | 20.03 |
| CCF-0471 | 2022;15 | 19.5 | 24.69 |
| CCF-0127 | 2022;11 | 19.51 | 19.86 |
| CCF-0436 | 2022;15 | 19.51 | 19.96 |
| CCF-0342 | 2022;14 | 19.56 | 19.66 |
| CCF-0312 | 2022;14 | 19.57 | 20.12 |
| CCF-0631 | 2022;15 | 19.59 | 19.31 |
| CCF-0429 | 2022;15 | 19.59 | 20.52 |
| CCF-0496 | 2022;15 | 19.67 | 19.82 |
| CCF-0590 | 2022;15 | 19.67 | 23.61 |
| CCF-0271 | 2022;14 | 19.68 | 19.32 |
| CCF-0355 | 2022;14 | 19.69 | 20.17 |
| CCF-0504 | 2022;15 | 19.69 | 24.11 |
| CCF-0415 | 2022;15 | 19.7 | 19.68 |
| CCF-0437 | 2022;15 | 19.79 | 20.01 |
| CCF-0602 | 2022;15 | 19.86 | 23.51 |
| CCF-0520 | 2022;15 | 19.88 | 19.83 |
| CCF-0852 | 2022;10 | 19.89 | 19.62 |
| CCF-0808 | 2022;11 | 19.93 | 19.81 |
| CCF-0490 | 2022;15 | 19.94 | 20.24 |
| CCF-0641 | 2022;15 | 19.95 | 23.63 |
| CCF-0456 | 2022;15 | 20 | 24.08 |
| CCF-0305 | 2022;14 | 20.03 | 19.86 |
| CCF-0283 | 2022;14 | 20.05 | 20.06 |
| CCF-0609 | 2022;15 | 20.08 | 19.71 |
| CCF-0536 | 2022;15 | 20.08 | 20.29 |

| | | | |
|---|---|---|---|
| CCF-0638 | 2022;15 | 20.1 | 23.97 |
| CCF-0273 | 2022;14 | 20.18 | 20.41 |
| CCF-0015 | 2022;10 | 20.22 | 20.58 |
| CCF-0495 | 2022;15 | 20.26 | 24.23 |
| CCF-0613 | 2022;15 | 20.27 | 19.91 |
| CCF-0477 | 2022;15 | 20.29 | 20.14 |
| CCF-0425 | 2022;15 | 20.3 | 20.75 |
| CCF-0646 | 2022;15 | 20.32 | 19.96 |
| CCF-0577 | 2022;15 | 20.34 | 20.09 |
| CCF-0386 | 2022;15 | 20.34 | 20.48 |
| CCF-0527 | 2022;15 | 20.35 | 20.42 |
| CCF-0573 | 2022;15 | 20.36 | 20.02 |
| CCF-0142 | 2022;12 | 20.38 | 20.43 |
| CCF-0012 | 2022;10 | 20.39 | 20.77 |
| CCF-0410 | 2022;15 | 20.44 | 24.31 |
| CCF-0482 | 2022;15 | 20.46 | 20.64 |
| CCF-0620 | 2022;15 | 20.48 | 20.06 |
| CCF-0505 | 2022;15 | 20.5 | 21.66 |
| CCF-0600 | 2022;15 | 20.54 | 24.05 |
| CCF-0711 | 2022;14 | 20.56 | 20.15 |
| CCF-0604 | 2022;15 | 20.58 | 20.18 |
| CCF-0304 | 2022;14 | 20.64 | 20.91 |
| CCF-0607 | 2022;15 | 20.65 | 20.2 |
| CCF-0317 | 2022;14 | 20.66 | 25.12 |
| CCF-0506 | 2022;15 | 20.68 | 25.19 |
| CCF-0461 | 2022;15 | 20.7 | 20.94 |
| CCF-0406 | 2022;15 | 20.71 | 20.67 |
| CCF-0543 | 2022;15 | 20.72 | 20.71 |
| CCF-0174 | 2022;13 | 20.76 | 20.91 |
| CCF-0430 | 2022;15 | 20.76 | 21.17 |
| CCF-0645 | 2022;15 | 20.83 | 20.67 |
| CCF-0580 | 2022;15 | 20.88 | 24.54 |
| CCF-0222 | 2022;13 | 20.88 | 25.77 |
| CCF-0540 | 2022;15 | 20.93 | 21.41 |
| CCF-0013 | 2022;10 | 20.95 | 20.89 |
| CCF-0357 | 2022;14 | 21.04 | 21.09 |
| CCF-0007 | 2022;10 | 21.04 | 21.2 |
| CCF-0358 | 2022;14 | 21.04 | 21.27 |
| CCF-0522 | 2022;15 | 21.07 | 21.17 |
| CCF-0160 | 2022;12 | 21.1 | 20.69 |
| CCF-0130 | 2022;11 | 21.15 | 21.64 |
| CCF-0455 | 2022;15 | 21.19 | 26.01 |
| CCF-0176 | 2022;13 | 21.22 | 21.35 |

| | | | |
|---|---|---|---|
| CCF-0454 | 2022;15 | 21.22 | 21.37 |
| CCF-0497 | 2022;15 | 21.22 | 21.94 |
| CCF-0173 | 2022;13 | 21.23 | 21.21 |
| CCF-0534 | 2022;15 | 21.26 | 21.33 |
| CCF-0516 | 2022;15 | 21.27 | 21.38 |
| CCF-0511 | 2022;15 | 21.37 | 21.5 |
| CCF-0642 | 2022;15 | 21.41 | 24.86 |
| CCF-0351 | 2022;14 | 21.44 | 25.34 |
| CCF-0549 | 2022;15 | 21.5 | 21.53 |
| CCF-0446 | 2022;15 | 21.56 | 21.65 |
| CCF-0453 | 2022;15 | 21.67 | 22.18 |
| CCF-0303 | 2022;14 | 21.7 | 22.46 |
| CCF-0591 | 2022;15 | 21.71 | 21.51 |
| CCF-0286 | 2022;14 | 21.72 | 21.56 |
| CCF-0568 | 2022;15 | 21.74 | 21.58 |
| CCF-0382 | 2022;15 | 21.74 | 21.77 |
| CCF-0621 | 2022;15 | 21.76 | 21.38 |
| CCF-0352 | 2022;14 | 21.78 | 21.92 |
| CCF-0750 | 2022;13 | 21.8 | 21.39 |
| CCF-0548 | 2022;15 | 21.8 | 21.99 |
| CCF-0817 | 2022;11 | 21.81 | 21.31 |
| CCF-0559 | 2022;15 | 21.82 | 21.64 |
| CCF-0623 | 2022;15 | 21.89 | 21.5 |
| CCF-0526 | 2022;15 | 21.94 | 22.13 |
| CCF-0364 | 2022;14 | 22.01 | 26.33 |
| CCF-0593 | 2022;15 | 22.02 | 21.68 |
| CCF-0353 | 2022;14 | 22.03 | 22.13 |
| CCF-0499 | 2022;15 | 22.06 | 27.03 |
| CCF-0619 | 2022;15 | 22.07 | 21.7 |
| CCF-0413 | 2022;15 | 22.08 | 22.24 |
| CCF-0670 | 2022;14 | 22.09 | 21.8 |
| CCF-0445 | 2022;15 | 22.12 | 22.29 |
| CCF-0366 | 2022;14 | 22.13 | 25.56 |
| CCF-0531 | 2022;15 | 22.17 | 23.3 |
| CCF-0407 | 2022;15 | 22.18 | 22.05 |
| CCF-0811 | 2022;11 | 22.19 | 21.95 |
| CCF-0748 | 2022;13 | 22.2 | 21.76 |
| CCF-0337 | 2022;14 | 22.21 | 27.09 |
| CCF-0408 | 2022;15 | 22.23 | 22.05 |
| CCF-0654 | 2022;14 | 22.25 | 21.96 |
| CCF-0301 | 2022;14 | 22.32 | 22.39 |
| CCF-0416 | 2022;15 | 22.37 | 22.42 |
| CCF-0402 | 2022;15 | 22.37 | 22.59 |

| | | | |
|---|---|---|---|
| CCF-0596 | 2022;15 | 22.39 | 21.97 |
| CCF-0363 | 2022;14 | 22.43 | 22.44 |
| CCF-0597 | 2022;15 | 22.44 | 21.92 |
| CCF-0401 | 2022;15 | 22.44 | 22.85 |
| CCF-0109 | 2022;11 | 22.45 | 22.57 |
| CCF-0332 | 2022;14 | 22.45 | 22.58 |
| CCF-0663 | 2022;14 | 22.51 | 22.09 |
| CCF-0279 | 2022;14 | 22.51 | 22.59 |
| CCF-0685 | 2022;14 | 22.51 | 25.96 |
| CCF-0372 | 2022;15 | 22.52 | 22.63 |
| CCF-0509 | 2022;15 | 22.57 | 22.7 |
| CCF-0574 | 2022;15 | 22.64 | 22.27 |
| CCF-0648 | 2022;15 | 22.73 | 22.18 |
| CCF-0581 | 2022;15 | 22.78 | 23.17 |
| CCF-0275 | 2022;14 | 22.79 | 22.86 |
| CCF-0484 | 2022;15 | 22.79 | 27.51 |
| CCF-0514 | 2022;15 | 22.8 | 23.04 |
| CCF-0699 | 2022;14 | 22.85 | 22.42 |
| CCF-0398 | 2022;15 | 22.91 | 27.58 |
| CCF-0272 | 2022;14 | 22.93 | 23.09 |
| CCF-0411 | 2022;15 | 23.01 | 23.69 |
| CCF-0758 | 2022;13 | 23.04 | 22.87 |
| CCF-0281 | 2022;14 | 23.06 | 23.1 |
| CCF-0291 | 2022;14 | 23.12 | 23.34 |
| CCF-0601 | 2022;15 | 23.19 | 22.79 |
| CCF-0679 | 2022;14 | 23.21 | 22.76 |
| CCF-0541 | 2022;15 | 23.27 | 23.89 |
| CCF-0463 | 2022;15 | 23.31 | 23.45 |
| CCF-0036 | 2022;10 | 23.43 | 23.61 |
| CCF-0635 | 2022;15 | 23.45 | 23.25 |
| CCF-0137 | 2022;11 | 23.49 | 23.44 |
| CCF-0681 | 2022;14 | 23.51 | 23.22 |
| CCF-0314 | 2022;14 | 23.52 | 24.03 |
| CCF-0008 | 2022;10 | 23.59 | 23.53 |
| CCF-0082 | 2022;11 | 23.6 | 23.42 |
| CCF-0360 | 2022;14 | 23.6 | 23.64 |
| CCF-0350 | 2022;14 | 23.6 | 23.74 |
| CCF-0624 | 2022;15 | 23.6 | 27.02 |
| CCF-0075 | 2022;12 | 23.63 | 23.65 |
| CCF-0420 | 2022;15 | 23.66 | 23.75 |
| CCF-0316 | 2022;14 | 23.67 | 23.86 |
| CCF-0660 | 2022;14 | 23.71 | 23.4 |
| CCF-0460 | 2022;15 | 23.71 | 23.97 |

| | | | |
|---|---|---|---|
| CCF-0435 | 2022;15 | 23.73 | 23.79 |
| CCF-0287 | 2022;14 | 23.75 | 24.26 |
| CCF-0369 | 2022;15 | 23.77 | 23.9 |
| CCF-0152 | 2022;12 | 23.81 | 23.8 |
| CCF-0269 | 2022;14 | 23.83 | 23.79 |
| CCF-0474 | 2022;15 | 24.03 | 23.98 |
| CCF-0570 | 2022;15 | 24.03 | 27.33 |
| CCF-0052 | 2022;10 | 24.06 | 24.14 |
| CCF-0503 | 2022;15 | 24.07 | 24.78 |
| CCF-0076 | 2022;12 | 24.15 | 24 |
| CCF-0146 | 2022;12 | 24.25 | 24.15 |
| CCF-0284 | 2022;14 | 24.28 | 29.1 |
| CCF-0320 | 2022;14 | 24.29 | 24.84 |
| CCF-0703 | 2022;14 | 24.32 | 24.01 |
| CCF-0365 | 2022;14 | 24.33 | 24.15 |
| CCF-0421 | 2022;15 | 24.34 | 24.25 |
| CCF-0424 | 2022;15 | 24.42 | 24.43 |
| CCF-0850 | 2022;10 | 24.62 | 23.93 |
| CCF-0083 | 2022;11 | 24.62 | 24.77 |
| CCF-0325 | 2022;14 | 24.65 | 25.02 |
| CCF-0105 | 2022;11 | 24.68 | 24.96 |
| CCF-0500 | 2022;15 | 24.68 | 25.48 |
| CCF-0121 | 2022;11 | 24.72 | 24.99 |
| CCF-0158 | 2022;12 | 24.73 | 24.88 |
| CCF-0392 | 2022;15 | 24.75 | 24.83 |
| CCF-0196 | 2022;12 | 24.75 | 35.25 |
| CCF-0494 | 2022;15 | 24.83 | 28.63 |
| CCF-0510 | 2022;15 | 24.84 | 24.96 |
| CCF-0721 | 2022;13 | 24.85 | 24.45 |
| CCF-0072 | 2022;11 | 24.87 | 25.22 |
| CCF-0473 | 2022;15 | 24.9 | 25.81 |
| CCF-0394 | 2022;15 | 24.92 | 25.62 |
| CCF-0387 | 2022;15 | 24.94 | 25.07 |
| CCF-0799 | 2022;11 | 24.96 | 24.59 |
| CCF-0694 | 2022;14 | 25 | 24.51 |
| CCF-0135 | 2022;11 | 25.02 | 25 |
| CCF-0545 | 2022;15 | 25.03 | 25.47 |
| CCF-0134 | 2022;11 | 25.11 | 25.08 |
| CCF-0611 | 2022;15 | 25.18 | 24.84 |
| CCF-0507 | 2022;15 | 25.18 | 25.18 |
| CCF-0705 | 2022;14 | 25.2 | 24.62 |
| CCF-0712 | 2022;14 | 25.23 | 24.58 |
| CCF-0014 | 2022;10 | 25.27 | 25.66 |

| | | | |
|---|---|---|---|
| CCF-0616 | 2022;15 | 25.29 | 24.73 |
| CCF-0539 | 2022;15 | 25.3 | 30.03 |
| CCF-0136 | 2022;11 | 25.36 | 25.48 |
| CCF-0171 | 2022;12 | 25.37 | 25.27 |
| CCF-0154 | 2022;12 | 25.39 | 25.48 |
| CCF-0667 | 2022;14 | 25.4 | 24.98 |
| CCF-0592 | 2022;15 | 25.51 | 28.97 |
| CCF-0367 | 2022;15 | 25.6 | 25.88 |
| CCF-0458 | 2022;15 | 25.7 | 25.91 |
| CCF-0359 | 2022;14 | 25.91 | 25.71 |
| CCF-0344 | 2022;14 | 26 | 26.18 |
| CCF-0277 | 2022;14 | 26.01 | 25.81 |
| CCF-0405 | 2022;15 | 26.04 | 25.72 |
| CCF-0569 | 2022;15 | 26.08 | 25.57 |
| CCF-0556 | 2022;15 | 26.09 | 25.98 |
| CCF-0098 | 2022;11 | 26.13 | 26.19 |
| CCF-0129 | 2022;11 | 26.14 | 26.28 |
| CCF-0400 | 2022;15 | 26.17 | 26.02 |
| CCF-0356 | 2022;14 | 26.18 | 26.45 |
| CCF-0584 | 2022;15 | 26.32 | 29.12 |
| CCF-0419 | 2022;15 | 26.32 | 30.47 |
| CCF-0653 | 2022;14 | 26.35 | 25.9 |
| CCF-0701 | 2022;14 | 26.4 | 26.5 |
| CCF-0562 | 2022;15 | 26.52 | 26.05 |
| CCF-0595 | 2022;15 | 26.58 | 26.29 |
| CCF-0859 | 2022;10 | 26.6 | 25.94 |
| CCF-0698 | 2022;14 | 26.63 | 26.19 |
| CCF-0558 | 2022;15 | 26.69 | 26.88 |
| CCF-0001 | 2022;10 | 26.69 | 27.15 |
| CCF-0466 | 2022;15 | 26.76 | 27.61 |
| CCF-0664 | 2022;14 | 26.78 | 26.56 |
| CCF-0011 | 2022;10 | 26.8 | 26.92 |
| CCF-0302 | 2022;14 | 26.8 | 26.98 |
| CCF-0502 | 2022;15 | 26.92 | 26.73 |
| CCF-0276 | 2022;14 | 27.18 | 27.19 |
| CCF-0388 | 2022;15 | 27.22 | 27.19 |
| CCF-0483 | 2022;15 | 27.31 | 19.74 |
| CCF-0032 | 2022;10 | 27.38 | 27.56 |
| CCF-0438 | 2022;15 | 27.42 | 27.52 |
| CCF-0554 | 2022;15 | 27.43 | 27.59 |
| CCF-0751 | 2022;13 | 27.45 | 26.84 |
| CCF-0561 | 2022;15 | 27.48 | 27.1 |
| CCF-0529 | 2022;15 | 27.5 | 31.52 |

| | | | |
|---|---|---|---|
| CCF-0094 | 2022;11 | 27.63 | 27.69 |
| CCF-0009 | 2022;10 | 27.73 | 27.61 |
| CCF-0216 | 2022;13 | 27.73 | 27.82 |
| CCF-0756 | 2022;13 | 27.78 | 27.13 |
| CCF-0289 | 2022;14 | 27.78 | 28.66 |
| CCF-0336 | 2022;14 | 27.8 | 27.68 |
| CCF-0644 | 2022;15 | 28.04 | 27.6 |
| CCF-0280 | 2022;14 | 28.06 | 28.03 |
| CCF-0669 | 2022;14 | 28.07 | 27.5 |
| CCF-0229 | 2022;13 | 28.1 | 28.26 |
| CCF-0028 | 2022;10 | 28.18 | 28.64 |
| CCF-0781 | 2022;12 | 28.22 | 27.67 |
| CCF-0533 | 2022;15 | 28.22 | 28.6 |
| CCF-0113 | 2022;11 | 28.23 | 28.23 |
| CCF-0716 | 2022;14 | 28.41 | 27.63 |
| CCF-0078 | 2022;12 | 28.46 | 28.39 |
| CCF-0467 | 2022;15 | 28.58 | 28.87 |
| CCF-0469 | 2022;15 | 28.62 | 28.94 |
| CCF-0390 | 2022;15 | 28.64 | 32.12 |
| CCF-0177 | 2022;12 | 28.69 | 28.64 |
| CCF-0056 | 2022;10 | 28.78 | 29.03 |
| CCF-0792 | 2022;12 | 28.82 | 28.33 |
| CCF-0293 | 2022;14 | 28.84 | 29.76 |
| CCF-0087 | 2022;11 | 28.89 | 29.08 |
| CCF-0521 | 2022;15 | 28.9 | 32.67 |
| CCF-0784 | 2022;12 | 28.92 | 28.26 |
| CCF-0740 | 2022;13 | 28.95 | 28.35 |
| CCF-0838 | 2022;10 | 28.95 | 28.45 |
| CCF-0447 | 2022;15 | 28.96 | 29.2 |
| CCF-0267 | 2022;14 | 28.97 | 29.2 |
| CCF-0274 | 2022;14 | 28.97 | 29.41 |
| CCF-0525 | 2022;15 | 29.03 | 29.44 |
| CCF-0035 | 2022;10 | 29.03 | 29.54 |
| CCF-0652 | 2022;14 | 29.06 | 28.52 |
| CCF-0418 | 2022;15 | 29.06 | 29.57 |
| CCF-0165 | 2022;12 | 29.19 | 29.11 |
| CCF-0175 | 2022;13 | 29.2 | 29.41 |
| CCF-0061 | 2022;10 | 29.38 | 29.71 |
| CCF-0214 | 2022;13 | 29.44 | 29.43 |
| CCF-0567 | 2022;15 | 29.49 | 31.75 |
| CCF-0185 | 2022;12 | 29.5 | 29.7 |
| CCF-0092 | 2022;11 | 29.5 | 29.96 |
| CCF-0399 | 2022;15 | 29.52 | 29.97 |

| | | | |
|---|---|---|---|
| CCF-0018 | 2022;10 | 29.53 | 29.85 |
| CCF-0444 | 2022;15 | 29.54 | 29.85 |
| CCF-0409 | 2022;15 | 29.56 | 33.64 |
| CCF-0647 | 2022;15 | 29.57 | 29.01 |
| CCF-0201 | 2022;13 | 29.6 | 29.61 |
| CCF-0151 | 2022;12 | 29.72 | 30.04 |
| CCF-0197 | 2022;12 | 29.79 | 30.27 |
| CCF-0809 | 2022;11 | 29.8 | 28.85 |
| CCF-0086 | 2022;11 | 29.81 | 29.88 |
| CCF-0125 | 2022;11 | 29.81 | 30.05 |
| CCF-0157 | 2022;12 | 29.83 | 30.62 |
| CCF-0815 | 2022;11 | 29.89 | 29.24 |
| CCF-0026 | 2022;10 | 29.94 | 30.14 |
| CCF-0371 | 2022;15 | 29.94 | 33.19 |
| CCF-0106 | 2022;11 | 29.99 | 30.67 |
| CCF-0848 | 2022;10 | 30.05 | 28.73 |
| CCF-0476 | 2022;15 | 30.1 | 31.52 |
| CCF-0170 | 2022;12 | 30.21 | 33.7 |
| CCF-0840 | 2022;10 | 30.34 | 29.69 |
| CCF-0131 | 2022;11 | 30.41 | 31.42 |
| CCF-0615 | 2022;15 | 30.45 | 29.56 |
| CCF-0308 | 2022;14 | 30.72 | 31.26 |
| CCF-0744 | 2022;13 | 30.78 | 29.52 |
| CCF-0687 | 2022;14 | 30.81 | 30.11 |
| CCF-0073 | 2022;11 | 30.85 | 31.25 |
| CCF-0262 | 2022;14 | 30.91 | 31.51 |
| CCF-0162 | 2022;12 | 30.94 | 31.34 |
| CCF-0002 | 2022;10 | 30.96 | 31.42 |
| CCF-0321 | 2022;14 | 30.97 | 31.63 |
| CCF-0391 | 2022;15 | 31.17 | 31.59 |
| CCF-0393 | 2022;15 | 31.24 | 32.2 |
| CCF-0847 | 2022;10 | 31.26 | 30.3 |
| CCF-0096 | 2022;11 | 31.26 | 31.51 |
| CCF-0563 | 2022;15 | 31.27 | 30.05 |
| CCF-0706 | 2022;14 | 31.34 | 30.27 |
| CCF-0240 | 2022;13 | 31.35 | 32.54 |
| CCF-0518 | 2022;15 | 31.4 | 31.98 |
| CCF-0285 | 2022;14 | 31.45 | 32.62 |
| CCF-0451 | 2022;15 | 31.56 | 32.75 |
| CCF-0816 | 2022;11 | 31.67 | 30.78 |
| CCF-0182 | 2022;12 | 31.67 | 32.05 |
| CCF-0849 | 2022;10 | 31.74 | 29.02 |
| CCF-0178 | 2022;12 | 31.74 | 32.2 |

| | | | |
|---|---|---|---|
| CCF-0589 | 2022;15 | 31.76 | 32.86 |
| CCF-0614 | 2022;15 | 31.77 | 30.3 |
| CCF-0347 | 2022;14 | 31.85 | 32.92 |
| CCF-0498 | 2022;15 | 31.94 | 32.89 |
| CCF-0211 | 2022;13 | 32.05 | 32.66 |
| CCF-0656 | 2022;14 | 32.07 | 31.08 |
| CCF-0585 | 2022;15 | 32.12 | 31.04 |
| CCF-0457 | 2022;15 | 32.15 | 35.1 |
| CCF-0789 | 2022;12 | 32.19 | 31.18 |
| CCF-0362 | 2022;14 | 32.24 | 33.16 |
| CCF-0340 | 2022;14 | 32.28 | 33.33 |
| CCF-0770 | 2022;12 | 32.31 | 31.73 |
| CCF-0475 | 2022;15 | 32.32 | 32.89 |
| CCF-0100 | 2022;11 | 32.36 | 32.88 |
| CCF-0017 | 2022;10 | 32.44 | 33.53 |
| CCF-0290 | 2022;14 | 32.45 | 36.18 |
| CCF-0449 | 2022;15 | 32.65 | 37.27 |
| CCF-0450 | 2022;15 | 32.65 | 37.27 |
| CCF-0235 | 2022;13 | 32.71 | 33.73 |
| CCF-0571 | 2022;15 | 32.75 | 31.84 |
| CCF-0546 | 2022;15 | 32.86 | 33.83 |
| CCF-0636 | 2022;15 | 32.91 | 31.85 |
| CCF-0237 | 2022;13 | 32.97 | 33.82 |
| CCF-0434 | 2022;15 | 33 | 34.08 |
| CCF-0836 | 2022;10 | 33.06 | 31.55 |
| CCF-0433 | 2022;15 | 33.07 | 34.28 |
| CCF-0818 | 2022;11 | 33.11 | 31.3 |
| CCF-0639 | 2022;15 | 33.17 | 31.09 |
| CCF-0217 | 2022;13 | 33.18 | 37.64 |
| CCF-0200 | 2022;13 | 33.21 | 33.92 |
| CCF-0046 | 2022;10 | 33.25 | 34.79 |
| CCF-0676 | 2022;14 | 33.29 | 31.29 |
| CCF-0755 | 2022;13 | 33.49 | 30.89 |
| CCF-0187 | 2022;12 | 33.49 | 35.2 |
| CCF-0329 | 2022;14 | 33.55 | 34.54 |
| CCF-0535 | 2022;15 | 33.55 | 36.79 |
| CCF-0097 | 2022;11 | 33.68 | 34.45 |
| CCF-0617 | 2022;15 | 33.73 | 32.54 |
| CCF-0295 | 2022;14 | 33.73 | 37.34 |
| CCF-0297 | 2022;14 | 33.73 | 37.34 |
| CCF-0040 | 2022;10 | 33.74 | 34.57 |
| CCF-0045 | 2022;10 | 33.75 | 35.09 |
| CCF-0103 | 2022;11 | 33.8 | 34.46 |

| | | | |
|---|---|---|---|
| CCF-0492 | 2022;15 | 33.86 | 34.97 |
| CCF-0432 | 2022;15 | 33.91 | 34.77 |
| CCF-0180 | 2022;12 | 33.96 | 35.04 |
| CCF-0834 | 2022;10 | 34 | 31.68 |
| CCF-0108 | 2022;11 | 34.07 | 34.82 |
| CCF-0704 | 2022;14 | 34.09 | 31.83 |
| CCF-0729 | 2022;13 | 34.09 | 32.9 |
| CCF-0238 | 2022;13 | 34.11 | 34.72 |
| CCF-0801 | 2022;11 | 34.2 | 31.68 |
| CCF-0468 | 2022;15 | 34.2 | 34.97 |
| CCF-0831 | 2022;10 | 34.22 | 32.95 |
| CCF-0796 | 2022;12 | 34.24 | 32.02 |
| CCF-0327 | 2022;14 | 34.24 | 35.1 |
| CCF-0779 | 2022;12 | 34.27 | 32.81 |
| CCF-0101 | 2022;11 | 34.27 | 35.2 |
| CCF-0169 | 2022;12 | 34.28 | 35.24 |
| CCF-0764 | 2022;12 | 34.29 | 32.46 |
| CCF-0576 | 2022;15 | 34.4 | 32.75 |
| CCF-0823 | 2022;11 | 34.43 | 32.79 |
| CCF-0515 | 2022;15 | 34.46 | 37.9 |
| CCF-0734 | 2022;13 | 34.47 | 32.17 |
| CCF-0069 | 2022;11 | 34.47 | 35.38 |
| CCF-0683 | 2022;14 | 34.49 | 32.76 |
| CCF-0788 | 2022;12 | 34.5 | 32.88 |
| CCF-0213 | 2022;13 | 34.52 | 35.79 |
| CCF-0728 | 2022;13 | 34.67 | 32.03 |
| CCF-0754 | 2022;13 | 34.77 | 32.67 |
| CCF-0557 | 2022;15 | 34.78 | 33.38 |
| CCF-0612 | 2022;15 | 34.88 | 32.4 |
| CCF-0427 | 2022;15 | 34.88 | 35.99 |
| CCF-0665 | 2022;14 | 34.98 | 33.04 |
| CCF-0780 | 2022;12 | 35.05 | 31.9 |
| CCF-0825 | 2022;11 | 35.06 | 33.66 |
| CCF-0249 | 2022;13 | 35.07 | 35.2 |
| CCF-0110 | 2022;11 | 35.12 | 35.49 |
| CCF-0310 | 2022;14 | 35.22 | 35.65 |
| CCF-0524 | 2022;15 | 35.29 | 35.92 |
| CCF-0067 | 2022;11 | 35.34 | 36.34 |
| CCF-0718 | 2022;13 | 35.43 | 34.05 |
| CCF-0172 | 2022;12 | 35.84 | 35.12 |
| CCF-0854 | 2022;10 | 36.03 | 34.36 |
| CCF-0120 | 2022;11 | 36.06 | 37.22 |
| CCF-0553 | 2022;15 | 36.22 | 35.4 |

| | | | |
|---|---|---|---|
| CCF-0422 | 2022;15 | 36.23 | 35.81 |
| CCF-0058 | 2022;10 | 36.36 | 35.96 |
| CCF-0594 | 2022;15 | 36.71 | 34.7 |
| CCF-0055 | 2022;10 | 37.41 | 36.4 |
| CCF-0810 | 2022;11 | 37.74 | 35.03 |
| Columbia 40 | 2022;11 | 15.6 | 16.1 |
| Columbia 122 | 2022;14 | 16.1 | 16.2 |
| Columbia 35 | 2022;11 | 16.3 | 20.9 |
| Columbia 103 | 2022;13 | 16.6 | 16.5 |
| Columbia 205 | 2022;15 | 17.1 | 16.8 |
| Columbia 24 | 2022;11 | 17.1 | 17 |
| Columbia 64 | 2022;12 | 17.4 | 17.3 |
| Columbia 102 | 2022;13 | 17.6 | 17.4 |
| Columbia 115 | 2022;14 | 17.7 | 17.6 |
| Columbia 136 | 2022;14 | 18 | 17.8 |
| Columbia 49 | 2022;12 | 18 | 18.2 |
| Columbia 197 | 2022;15 | 18.1 | 23 |
| Columbia 19 | 2022;10 | 18.3 | 18.3 |
| Columbia 42 | 2022;12 | 18.4 | 17.8 |
| Columbia 131 | 2022;14 | 18.6 | 18.6 |
| Columbia 96 | 2022;13 | 18.7 | 18.4 |
| Columbia 143 | 2022;14 | 18.9 | 18.6 |
| Columbia 147 | 2022;14 | 19.2 | 18.8 |
| Columbia 106 | 2022;13 | 19.2 | 18.9 |
| Columbia 130 | 2022;14 | 19.3 | 19.2 |
| Columbia 152 | 2022;14 | 19.3 | 19.6 |
| Columbia 177 | 2022;15 | 19.4 | 19.5 |
| Columbia 128 | 2022;14 | 19.5 | 24 |
| Columbia 29 | 2022;11 | 19.7 | 19.4 |
| Columbia 121 | 2022;14 | 19.8 | 19.4 |
| Columbia 94 | 2022;13 | 19.8 | 19.8 |
| Columbia 192 | 2022;15 | 20.2 | 20.1 |
| Columbia 110 | 2022;14 | 20.4 | 20.1 |
| Columbia 23 | 2022;11 | 20.6 | 19.9 |
| Columbia 190 | 2022;15 | 20.6 | 21.1 |
| Columbia 200 | 2022;15 | 20.7 | 25.6 |
| Columbia 98 | 2022;13 | 20.9 | 20.5 |
| Columbia 12 | 2022;10 | 21.2 | 21 |
| Columbia 86 | 2022;13 | 21.2 | 21.1 |
| Columbia 107 | 2022;13 | 21.3 | 21.3 |
| Columbia 1 | 2022;10 | 21.4 | 21 |
| Columbia 90 | 2022;13 | 21.5 | 21 |
| Columbia 120 | 2022;14 | 21.5 | 21.4 |

| | | | |
|---|---|---|---|
| Columbia 88 | 2022;13 | 21.8 | 21.6 |
| Columbia 125 | 2022;14 | 21.9 | 21.6 |
| Columbia 199 | 2022;15 | 21.9 | 22.1 |
| Columbia 209 | 2022;15 | 22.5 | 22.4 |
| Columbia 25 | 2022;11 | 22.5 | 22.7 |
| Columbia 81 | 2022;13 | 22.6 | 22.4 |
| Columbia 58 | 2022;12 | 22.6 | 27 |
| Columbia 45 | 2022;12 | 22.8 | 22.5 |
| Columbia 183 | 2022;15 | 22.8 | 23.4 |
| Columbia 85 | 2022;13 | 23 | 22.8 |
| Columbia 175 | 2022;15 | 23.1 | 23.1 |
| Columbia 28 | 2022;11 | 23.2 | 23.4 |
| Columbia 196 | 2022;15 | 23.3 | 23.2 |
| Columbia 99 | 2022;13 | 23.4 | 22.4 |
| Columbia 17 | 2022;10 | 23.4 | 23.1 |
| Columbia 211 | 2022;15 | 23.4 | 23.3 |
| Columbia 39 | 2022;11 | 23.6 | 23.6 |
| Columbia 186 | 2022;15 | 23.8 | 23.8 |
| Columbia 34 | 2022;11 | 24 | 24 |
| Columbia 149 | 2022;14 | 24 | 24.2 |
| Columbia 116 | 2022;14 | 24.2 | 24 |
| Columbia 166 | 2022;15 | 24.2 | 24.1 |
| Columbia 201 | 2022;15 | 24.3 | 24.4 |
| Columbia 154 | 2022;14 | 24.4 | 24.4 |
| Columbia 161 | 2022;15 | 24.5 | 24 |
| Columbia 109 | 2022;14 | 24.5 | 24.4 |
| Columbia 65 | 2022;12 | 24.5 | 24.6 |
| Columbia 87 | 2022;13 | 24.6 | 24.4 |
| Columbia 213 | 2022;15 | 24.6 | 27.8 |
| Columbia 44 | 2022;12 | 25 | 24.4 |
| Columbia 173 | 2022;15 | 25.2 | 25.2 |
| Columbia 15 | 2022;10 | 25.6 | 24.9 |
| Columbia 178 | 2022;15 | 25.6 | 25.3 |
| Columbia 30 | 2022;11 | 25.9 | 26.1 |
| Columbia 50 | 2022;12 | 26.2 | 26.8 |
| Columbia 119 | 2022;14 | 26.3 | 26 |
| Columbia 80 | 2022;13 | 26.4 | 26.1 |
| Columbia 118 | 2022;14 | 26.7 | 26.4 |
| Columbia 117 | 2022;14 | 27.1 | 26.5 |
| Columbia 124 | 2022;14 | 27.5 | 27.2 |
| Columbia 4 | 2022;10 | 27.7 | 27.7 |
| Columbia 206 | 2022;15 | 28.1 | 28 |
| Columbia 76 | 2022;13 | 28.6 | 28 |

| | | | |
|---|---|---|---|
| Columbia 168 | 2022;15 | 28.8 | 28.9 |
| Columbia 137 | 2022;14 | 29 | 28.8 |
| Columbia 146 | 2022;14 | 29 | 29.2 |
| Columbia 112 | 2022;14 | 29.1 | 28.3 |
| Columbia 38 | 2022;11 | 29.1 | 29.1 |
| Columbia 77 | 2022;13 | 29.3 | 28.9 |
| Columbia 203 | 2022;15 | 29.3 | 28.9 |
| Columbia 105 | 2022;13 | 29.5 | 33.5 |
| Columbia 37 | 2022;11 | 29.6 | 30.3 |
| Columbia 151 | 2022;14 | 30.4 | 29.7 |
| Columbia 170 | 2022;15 | 30.5 | 31.8 |
| Columbia 95 | 2022;13 | 30.5 | 34.2 |
| Columbia 70 | 2022;12 | 30.6 | 30 |
| Columbia 210 | 2022;15 | 30.6 | 31 |
| Columbia 172 | 2022;15 | 30.8 | 30.3 |
| Columbia 164 | 2022;15 | 30.9 | 32.6 |
| Columbia 100 | 2022;13 | 31 | 29.9 |
| Columbia 202 | 2022;15 | 31.2 | 33.5 |
| Columbia 111 | 2022;14 | 31.2 | 34.7 |
| Columbia 33 | 2022;11 | 31.5 | 30.7 |
| Columbia 83 | 2022;13 | 31.7 | 30.8 |
| Columbia 78 | 2022;13 | 31.7 | 30.9 |
| Columbia 158 | 2022;14 | 31.7 | 32.6 |
| Columbia 92 | 2022;13 | 31.8 | 31 |
| Columbia 148 | 2022;14 | 31.8 | 32.6 |
| Columbia 60 | 2022;12 | 32 | 30.9 |
| Columbia 3 | 2022;10 | 32.1 | 32.8 |
| Columbia 174 | 2022;15 | 32.1 | 35.3 |
| Columbia 184 | 2022;15 | 32.3 | 31.1 |
| Columbia 13 | 2022;10 | 32.4 | 33.5 |
| Columbia 160 | 2022;15 | 32.5 | 31.2 |
| Columbia 55 | 2022;12 | 32.5 | 31.7 |
| Columbia 113 | 2022;14 | 32.6 | 30.9 |
| Columbia 46 | 2022;12 | 32.6 | 31.9 |
| Columbia 123 | 2022;14 | 32.6 | 33.5 |
| Columbia 67 | 2022;12 | 32.7 | 32 |
| Columbia 159 | 2022;14 | 32.7 | 36 |
| Columbia 18 | 2022;10 | 32.8 | 31.5 |
| Columbia 52 | 2022;12 | 33 | 33.4 |
| Columbia 56 | 2022;12 | 33 | 33.8 |
| Columbia 188 | 2022;15 | 33.3 | 32.1 |
| Columbia 145 | 2022;14 | 33.4 | 32.3 |
| Columbia 57 | 2022;12 | 33.4 | 34.1 |

| | | | |
|---|---|---|---|
| Columbia 97 | 2022;13 | 33.5 | 34.4 |
| Columbia 198 | 2022;15 | 33.7 | 33 |
| Columbia 21 | 2022;10 | 33.8 | 32.8 |
| Columbia 153 | 2022;14 | 33.9 | 34.8 |
| Columbia 108 | 2022;13 | 34.1 | 33.4 |
| Columbia 194 | 2022;15 | 34.4 | 34.2 |
| Columbia 150 | 2022;14 | 34.7 | 33.4 |
| Columbia 10 | 2022;10 | 34.8 | 34.2 |
| Columbia 126 | 2022;14 | 34.9 | 32.6 |
| Columbia 2 | 2022;10 | 34.9 | 37.7 |
| Columbia 169 | 2022;15 | 35 | 33.9 |
| Columbia 36 | 2022;11 | 35 | 36.8 |
| Columbia 16 | 2022;10 | 35.2 | 35.2 |
| Columbia 155 | 2022;14 | 35.2 | 35.7 |
| Columbia 133 | 2022;14 | 35.3 | 34 |
| Columbia 47 | 2022;12 | 35.3 | 34.5 |
| Columbia 212 | 2022;15 | 35.4 | 35.4 |
| Columbia 157 | 2022;14 | 35.5 | 36.2 |
| Columbia 93 | 2022;13 | 35.6 | 35 |
| Columbia 6 | 2022;10 | 35.6 | 37.7 |
| Columbia 135 | 2022;14 | 35.8 | 33.8 |
| Columbia 7 | 2022;10 | 35.8 | 36.1 |
| Columbia 89 | 2022;13 | 36 | 34.1 |
| Columbia 31 | 2022;11 | 36 | 34.7 |
| Columbia 5 | 2022;10 | 36 | 36.9 |
| Columbia 142 | 2022;14 | 36.1 | 35 |
| Columbia 180 | 2022;15 | 36.2 | 37 |
| Columbia 63 | 2022;12 | 36.4 | 35.3 |
| Columbia 74 | 2022;13 | 36.5 | 33.4 |
| Columbia 127 | 2022;14 | 37.2 | 37.3 |
| WCMC-0036 | 2022;12 | 15.5 | 15.5 |
| WCMC-0069 | 2022;13 | 16.3 | 16.3 |
| WCMC-0031 | 2022;11 | 16.4 | 16.9 |
| WCMC-0073 | 2022;13 | 17 | 16.9 |
| WCMC-0090 | 2022;14 | 17.2 | 17.6 |
| WCMC-0026 | 2022;11 | 17.9 | 17.8 |
| WCMC-0054 | 2022;12 | 18.6 | 18.2 |
| WCMC-0041 | 2022;12 | 19.2 | 18.8 |
| WCMC-0114 | 2022;14 | 19.4 | 19.2 |
| WCMC-0108 | 2022;14 | 19.6 | 19.2 |
| WCMC-0080 | 2022;13 | 19.8 | 19.5 |
| WCMC-0003 | 2022;10 | 20 | 19.9 |
| WCMC-0021 | 2022;11 | 20 | 19.9 |

| | | | |
|---|---|---|---|
| WCMC-0079 | 2022;13 | 20 | 19.9 |
| WCMC-0082 | 2022;13 | 20 | 19.9 |
| WCMC-0101 | 2022;14 | 20.1 | 19.7 |
| WCMC-0078 | 2022;13 | 20.3 | 20 |
| WCMC-0008 | 2022;10 | 20.3 | 20.2 |
| WCMC-0117 | 2022;15 | 20.5 | 20.4 |
| WCMC-0061 | 2022;13 | 20.8 | 20.5 |
| WCMC-0023 | 2022;11 | 21.4 | 21.5 |
| WCMC-0024 | 2022;11 | 21.8 | 21.6 |
| WCMC-0119 | 2022;15 | 21.9 | 21.6 |
| WCMC-0050 | 2022;12 | 22.2 | 21.9 |
| WCMC-0059 | 2022;13 | 22.2 | 22.4 |
| WCMC-0063 | 2022;13 | 22.5 | 22.4 |
| WCMC-0049 | 2022;12 | 23.3 | 23.2 |
| WCMC-0056 | 2022;12 | 23.5 | 23 |
| WCMC-0113 | 2022;14 | 23.5 | 23.2 |
| WCMC-0032 | 2022;11 | 23.5 | 23.5 |
| WCMC-0066 | 2022;13 | 23.6 | 23.4 |
| WCMC-0070 | 2022;13 | 24 | 24.1 |
| WCMC-0074 | 2022;13 | 24.2 | 23.9 |
| WCMC-0077 | 2022;13 | 24.3 | 23.9 |
| WCMC-0025 | 2022;11 | 24.4 | 24.2 |
| WCMC-0044 | 2022;12 | 24.8 | 24.4 |
| WCMC-0012 | 2022;10 | 25.1 | 24.6 |
| WCMC-0014 | 2022;10 | 25.1 | 24.9 |
| WCMC-0046 | 2022;12 | 25.2 | 25 |
| WCMC-0083 | 2022;13 | 25.4 | 24.9 |
| WCMC-0096 | 2022;14 | 25.4 | 25.2 |
| WCMC-0099 | 2022;14 | 26 | 25.7 |
| WCMC-0037 | 2022;12 | 26.3 | 25.7 |
| WCMC-0053 | 2022;12 | 26.3 | 26 |
| WCMC-0047 | 2022;12 | 26.5 | 26.3 |
| WCMC-0067 | 2022;13 | 26.6 | 26.2 |
| WCMC-0087 | 2022;14 | 26.8 | 29.9 |
| WCMC-0093 | 2022;14 | 26.8 | 29.9 |
| WCMC-0089 | 2022;14 | 27.3 | 27.1 |
| WCMC-0040 | 2022;12 | 27.5 | 27.1 |
| WCMC-0092 | 2022;14 | 27.5 | 27.2 |
| WCMC-0042 | 2022;12 | 27.7 | 27.3 |
| WCMC-0058 | 2022;13 | 27.7 | 27.3 |
| WCMC-0105 | 2022;14 | 27.7 | 27.5 |
| WCMC-0048 | 2022;12 | 27.8 | 27.3 |
| WCMC-0029 | 2022;11 | 28.2 | 27.7 |

| | | | |
|---|---|---|---|
| WCMC-0005 | 2022;10 | 28.5 | 28.2 |
| WCMC-0011 | 2022;10 | 29.7 | 29.3 |
| WCMC-0010 | 2022;10 | 29.8 | 29.3 |
| WCMC-0106 | 2022;14 | 29.9 | 29.4 |
| WCMC-0016 | 2022;10 | 30 | 29.6 |
| WCMC-0081 | 2022;13 | 30.5 | 29.8 |
| WCMC-0022 | 2022;11 | 31.2 | 30 |
| WCMC-0038 | 2022;12 | 31.4 | 30.2 |
| WCMC-0004 | 2022;10 | 31.6 | 30.6 |
| WCMC-0027 | 2022;11 | 31.6 | 30.6 |
| WCMC-0039 | 2022;12 | 31.7 | 31 |
| WCMC-0076 | 2022;13 | 31.8 | 30.7 |
| WCMC-0075 | 2022;13 | 31.9 | 30.7 |
| WCMC-0100 | 2022;14 | 32.1 | 31.2 |
| WCMC-0120 | 2022;15 | 32.1 | 34.2 |
| WCMC-0001 | 2022;10 | 32.2 | 30.7 |
| WCMC-0094 | 2022;14 | 32.2 | 30.8 |
| WCMC-0098 | 2022;14 | 32.4 | 31.2 |
| WCMC-0043 | 2022;12 | 32.6 | 31 |
| WCMC-0057 | 2022;13 | 32.6 | 31.7 |
| WCMC-0064 | 2022;13 | 32.9 | 31.2 |
| WCMC-0111 | 2022;14 | 33.3 | 31.9 |
| WCMC-0007 | 2022;10 | 33.5 | 32 |
| WCMC-0112 | 2022;14 | 33.6 | 32.8 |
| WCMC-0103 | 2022;14 | 33.7 | 31.9 |
| WCMC-0002 | 2022;10 | 33.9 | 31.9 |
| WCMC-0086 | 2022;13 | 34.3 | 31.9 |
| WCMC-0055 | 2022;12 | 34.3 | 32.7 |
| WCMC-0017 | 2022;10 | 34.3 | 33 |
| WCMC-0018 | 2022;11 | 34.4 | 33.2 |
| WCMC-0034 | 2022;12 | 34.5 | 32.1 |
| WCMC-0045 | 2022;12 | 34.6 | 33 |
| WCMC-0107 | 2022;14 | 34.7 | 33.7 |
| WCMC-0019 | 2022;11 | 34.8 | 32.6 |
| WCMC-0060 | 2022;13 | 34.8 | 33 |
| WCMC-0062 | 2022;13 | 34.8 | 33.7 |
| WCMC-0020 | 2022;11 | 35.1 | 33.5 |
| WCMC-0071 | 2022;13 | 35.4 | 33.7 |
| WCMC-0109 | 2022;14 | 35.6 | 33.5 |
| WCMC-0085 | 2022;13 | 35.7 | 33.8 |
| WCMC-0035 | 2022;12 | 35.7 | 34.5 |
| WCMC-0030 | 2022;11 | 36.3 | 35.1 |
| WCMC-0104 | 2022;14 | 36.6 | 33.3 |

| | | | |
|---|---|---|---|
| WCMC-0009 | 2022;10 | 37.5 | 35.3 |
| Penn296 | 2022;15 | 15 | 15.2 |
| Penn224 | 2022;14 | 15.3 | 15.2 |
| Penn299 | 2022;15 | 15.5 | 15.1 |
| Penn310 | 2022;15 | 15.5 | 19.1 |
| Penn232 | 2022;14 | 15.8 | 19.8 |
| Penn356 | 2022;15 | 16.1 | 20 |
| Penn135 | 2022;13 | 16.2 | 19.1 |
| Penn166 | 2022;13 | 16.2 | 19.9 |
| Penn270 | 2022;14 | 16.4 | 16.2 |
| Penn158 | 2022;13 | 16.4 | 16.3 |
| Penn140 | 2022;13 | 16.4 | 16.4 |
| Penn307 | 2022;15 | 16.7 | 16.2 |
| Penn177 | 2022;13 | 16.7 | 16.7 |
| Penn122 | 2022;12 | 16.8 | 16.5 |
| Penn319 | 2022;15 | 16.8 | 16.5 |
| Penn298 | 2022;15 | 16.9 | 16.6 |
| Penn217 | 2022;14 | 16.9 | 21 |
| Penn180 | 2022;13 | 17 | 20.4 |
| Penn117 | 2022;12 | 17.1 | 17 |
| Penn131 | 2022;13 | 17.1 | 20.3 |
| Penn283 | 2022;15 | 17.1 | 20.6 |
| Penn316 | 2022;15 | 17.1 | 20.9 |
| Penn66 | 2022;11 | 17.2 | 16.9 |
| Penn290 | 2022;15 | 17.2 | 21 |
| Penn313 | 2022;15 | 17.3 | 16.8 |
| Penn253 | 2022;14 | 17.3 | 17 |
| Penn241 | 2022;14 | 17.3 | 17.2 |
| Penn154 | 2022;13 | 17.3 | 20.9 |
| Penn252 | 2022;14 | 17.3 | 21.4 |
| Penn206 | 2022;14 | 17.4 | 17 |
| Penn99 | 2022;12 | 17.4 | 17.2 |
| Penn259 | 2022;14 | 17.4 | 21.2 |
| Penn244 | 2022;14 | 17.5 | 17.5 |
| Penn202 | 2022;14 | 17.5 | 21.3 |
| Penn208 | 2022;14 | 17.5 | 21.3 |
| Penn346 | 2022;15 | 17.5 | 21.4 |
| Penn182 | 2022;13 | 17.6 | 17.3 |
| Penn176 | 2022;13 | 17.6 | 21.4 |
| Penn173 | 2022;13 | 17.7 | 17.5 |
| Penn355 | 2022;15 | 17.7 | 22 |
| Penn352 | 2022;15 | 17.8 | 17.4 |
| Penn201 | 2022;14 | 17.8 | 21.7 |

| | | | |
|---|---|---|---|
| Penn225 | 2022;14 | 17.8 | 22 |
| Penn362 | 2022;15 | 17.9 | 22 |
| Penn139 | 2022;13 | 18 | 17.8 |
| Penn143 | 2022;13 | 18 | 17.8 |
| Penn86 | 2022;12 | 18 | 17.9 |
| Penn267 | 2022;14 | 18 | 17.9 |
| Penn141 | 2022;13 | 18 | 18.4 |
| Penn221 | 2022;14 | 18 | 18.9 |
| Penn304 | 2022;15 | 18 | 21.6 |
| Penn282 | 2022;15 | 18.1 | 17.8 |
| Penn306 | 2022;15 | 18.2 | 18.1 |
| Penn235 | 2022;14 | 18.2 | 22 |
| Penn210 | 2022;14 | 18.2 | 22.2 |
| Penn328 | 2022;15 | 18.2 | 22.4 |
| Penn226 | 2022;14 | 18.3 | 18.1 |
| Penn246 | 2022;14 | 18.3 | 18.4 |
| Penn183 | 2022;13 | 18.4 | 22 |
| Penn150 | 2022;13 | 18.5 | 21.6 |
| Penn245 | 2022;14 | 18.6 | 22.1 |
| Penn357 | 2022;15 | 18.7 | 22.3 |
| Penn311 | 2022;15 | 18.7 | 22.6 |
| Penn325 | 2022;15 | 18.8 | 18.4 |
| Penn231 | 2022;14 | 18.8 | 18.5 |
| Penn236 | 2022;14 | 18.8 | 22.8 |
| Penn193 | 2022;13 | 18.9 | 18.5 |
| Penn98 | 2022;12 | 18.9 | 18.7 |
| Penn268 | 2022;14 | 18.9 | 18.7 |
| Penn58 | 2022;11 | 18.9 | 18.8 |
| Penn130 | 2022;13 | 19.1 | 18.8 |
| Penn172 | 2022;13 | 19.1 | 18.9 |
| Penn336 | 2022;15 | 19.1 | 23.2 |
| Penn212 | 2022;14 | 19.1 | 23.5 |
| Penn308 | 2022;15 | 19.2 | 19.1 |
| Penn300 | 2022;15 | 19.2 | 23.2 |
| Penn179 | 2022;13 | 19.2 | 23.4 |
| Penn314 | 2022;15 | 19.3 | 22.8 |
| Penn309 | 2022;15 | 19.3 | 23.4 |
| Penn233 | 2022;14 | 19.4 | 23.2 |
| Penn312 | 2022;15 | 19.5 | 23.3 |
| Penn271 | 2022;14 | 19.6 | 19.2 |
| Penn255 | 2022;14 | 19.6 | 23.9 |
| Penn132 | 2022;13 | 19.8 | 19.4 |
| Penn264 | 2022;14 | 19.8 | 23.4 |

| | | | |
|---|---|---|---|
| Penn112 | 2022;12 | 19.8 | 23.7 |
| Penn289 | 2022;15 | 19.9 | 19.7 |
| Penn243 | 2022;14 | 19.9 | 20.1 |
| Penn293 | 2022;15 | 20 | 23.8 |
| Penn90 | 2022;12 | 20.1 | 20.1 |
| Penn204 | 2022;14 | 20.1 | 20.1 |
| Penn321 | 2022;15 | 20.1 | 24.1 |
| Penn138 | 2022;13 | 20.1 | 24.2 |
| Penn129 | 2022;13 | 20.2 | 20 |
| Penn170 | 2022;13 | 20.2 | 20.7 |
| Penn323 | 2022;15 | 20.2 | 24.3 |
| Penn157 | 2022;13 | 20.4 | 20.2 |
| Penn276 | 2022;14 | 20.4 | 20.2 |
| Penn242 | 2022;14 | 20.5 | 20.2 |
| Penn367 | 2022;15 | 20.5 | 24.4 |
| Penn317 | 2022;15 | 20.5 | 24.5 |
| Penn335 | 2022;15 | 20.5 | 24.5 |
| Penn133 | 2022;13 | 20.6 | 20.2 |
| Penn187 | 2022;13 | 20.6 | 20.5 |
| Penn199 | 2022;13 | 20.6 | 24.7 |
| Penn195 | 2022;13 | 20.7 | 20.4 |
| Penn91 | 2022;12 | 20.7 | 21.2 |
| Penn185 | 2022;13 | 20.7 | 24.5 |
| Penn111 | 2022;12 | 20.8 | 20.3 |
| Penn287 | 2022;15 | 20.8 | 20.7 |
| Penn164 | 2022;13 | 20.8 | 24.7 |
| Penn366 | 2022;15 | 20.8 | 24.9 |
| Penn163 | 2022;13 | 20.8 | 25 |
| Penn160 | 2022;13 | 20.9 | 24.4 |
| Penn234 | 2022;14 | 21 | 20.8 |
| Penn155 | 2022;13 | 21 | 21 |
| Penn361 | 2022;15 | 21 | 24.8 |
| Penn216 | 2022;14 | 21.1 | 20.7 |
| Penn301 | 2022;15 | 21.1 | 25 |
| Penn331 | 2022;15 | 21.2 | 21.1 |
| Penn100 | 2022;12 | 21.3 | 21 |
| Penn165 | 2022;13 | 21.3 | 21.4 |
| Penn192 | 2022;13 | 21.4 | 22 |
| Penn266 | 2022;14 | 21.4 | 25.4 |
| Penn104 | 2022;12 | 21.5 | 21.2 |
| Penn237 | 2022;14 | 21.5 | 24.9 |
| Penn152 | 2022;13 | 21.6 | 21.4 |
| Penn137 | 2022;13 | 21.7 | 21.5 |

| | | | |
|---|---|---|---|
| Penn126 | 2022;13 | 21.7 | 22.1 |
| Penn96 | 2022;12 | 21.8 | 21.6 |
| Penn239 | 2022;14 | 21.8 | 25.5 |
| Penn134 | 2022;13 | 22 | 21.5 |
| Penn105 | 2022;12 | 22.1 | 21.6 |
| Penn348 | 2022;15 | 22.1 | 26.1 |
| Penn349 | 2022;15 | 22.2 | 26.3 |
| Penn169 | 2022;13 | 22.5 | 26.2 |
| Penn186 | 2022;13 | 22.6 | 22.4 |
| Penn360 | 2022;15 | 22.6 | 22.4 |
| Penn248 | 2022;14 | 22.6 | 25.9 |
| Penn110 | 2022;12 | 22.6 | 26.3 |
| Penn343 | 2022;15 | 22.6 | 26.3 |
| Penn272 | 2022;14 | 22.7 | 22.5 |
| Penn263 | 2022;14 | 23 | 22.8 |
| Penn119 | 2022;12 | 23 | 23 |
| Penn175 | 2022;13 | 23 | 26.7 |
| Penn190 | 2022;13 | 23.1 | 22.8 |
| Penn238 | 2022;14 | 23.1 | 22.8 |
| Penn363 | 2022;15 | 23.1 | 22.8 |
| Penn219 | 2022;14 | 23.2 | 23 |
| Penn114 | 2022;12 | 23.4 | 23.1 |
| Penn341 | 2022;15 | 23.4 | 27 |
| Penn258 | 2022;14 | 23.6 | 27.3 |
| Penn162 | 2022;13 | 23.7 | 23 |
| Penn284 | 2022;15 | 23.7 | 23.4 |
| Penn369 | 2022;15 | 23.7 | 23.5 |
| Penn151 | 2022;13 | 23.8 | 23.4 |
| Penn136 | 2022;13 | 23.9 | 23.8 |
| Penn344 | 2022;15 | 23.9 | 26.8 |
| Penn167 | 2022;13 | 24.1 | 24 |
| Penn174 | 2022;13 | 24.2 | 23.6 |
| Penn372 | 2022;15 | 24.2 | 23.7 |
| Penn115 | 2022;12 | 24.2 | 23.8 |
| Penn303 | 2022;15 | 24.2 | 24 |
| Penn334 | 2022;15 | 24.4 | 23.9 |
| Penn256 | 2022;14 | 24.4 | 24.2 |
| Penn342 | 2022;15 | 24.4 | 27.9 |
| Penn275 | 2022;14 | 24.8 | 24.3 |
| Penn6 | 2022;10 | 24.8 | 24.4 |
| Penn200 | 2022;13 | 24.8 | 24.4 |
| Penn147 | 2022;13 | 24.9 | 24.9 |
| Penn214 | 2022;14 | 24.9 | 25 |

| | | | |
|---|---|---|---|
| Penn222 | 2022;14 | 25 | 24.7 |
| Penn327 | 2022;15 | 25 | 24.7 |
| Penn305 | 2022;15 | 25 | 24.9 |
| Penn277 | 2022;14 | 25.1 | 24.5 |
| Penn288 | 2022;15 | 25.1 | 24.8 |
| Penn332 | 2022;15 | 25.4 | 28.5 |
| Penn159 | 2022;13 | 25.5 | 25.3 |
| Penn274 | 2022;14 | 25.6 | 29.1 |
| Penn125 | 2022;12 | 25.7 | 25.3 |
| Penn44 | 2022;11 | 25.8 | 25.6 |
| Penn353 | 2022;15 | 25.8 | 25.8 |
| Penn359 | 2022;15 | 25.8 | 28.7 |
| Penn351 | 2022;15 | 25.8 | 28.8 |
| Penn121 | 2022;12 | 26 | 25.9 |
| Penn197 | 2022;13 | 26 | 29.4 |
| Penn261 | 2022;14 | 26.6 | 25.9 |
| Penn92 | 2022;12 | 26.6 | 29.6 |
| Penn326 | 2022;15 | 26.7 | 26.3 |
| Penn113 | 2022;12 | 26.9 | 26.6 |
| Penn178 | 2022;13 | 27.2 | 30.1 |
| Penn324 | 2022;15 | 27.2 | 30.4 |
| Penn194 | 2022;13 | 27.4 | 27.1 |
| Penn142 | 2022;13 | 27.6 | 27.3 |
| Penn220 | 2022;14 | 27.7 | 27.6 |
| Penn161 | 2022;13 | 27.7 | 30.6 |
| Penn118 | 2022;12 | 27.8 | 27.6 |
| Penn358 | 2022;15 | 27.8 | 27.6 |
| Penn67 | 2022;11 | 27.8 | 27.7 |
| Penn109 | 2022;12 | 28 | 28.1 |
| Penn93 | 2022;12 | 28.1 | 27.4 |
| Penn116 | 2022;12 | 28.2 | 31.2 |
| Penn315 | 2022;15 | 28.3 | 28.9 |
| Penn257 | 2022;14 | 28.3 | 30.9 |
| Penn36 | 2022;10 | 28.9 | 28.1 |
| Penn188 | 2022;13 | 28.9 | 28.4 |
| Penn223 | 2022;14 | 28.9 | 31.1 |
| Penn87 | 2022;12 | 29.1 | 28.6 |
| Penn24 | 2022;10 | 29.1 | 29.2 |
| Penn333 | 2022;15 | 29.2 | 31.6 |
| Penn203 | 2022;14 | 29.4 | 29.6 |
| Penn123 | 2022;12 | 29.5 | 31.7 |
| Penn81 | 2022;12 | 29.6 | 28.9 |
| Penn27 | 2022;10 | 29.6 | 29.1 |

| | | | |
|---|---|---|---|
| Penn14 | 2022;10 | 29.6 | 29.2 |
| Penn74 | 2022;12 | 29.6 | 29.2 |
| Penn153 | 2022;13 | 29.6 | 29.4 |
| Penn230 | 2022;14 | 29.6 | 32 |
| Penn102 | 2022;12 | 29.7 | 29.3 |
| Penn89 | 2022;12 | 29.7 | 29.6 |
| Penn302 | 2022;15 | 30 | 31.9 |
| Penn205 | 2022;14 | 30 | 32.6 |
| Penn149 | 2022;13 | 30.4 | 29.9 |
| Penn292 | 2022;15 | 30.5 | 31.2 |
| Penn49 | 2022;11 | 30.6 | 29.6 |
| Penn229 | 2022;14 | 30.7 | 32.7 |
| Penn291 | 2022;15 | 30.8 | 30 |
| Penn228 | 2022;14 | 30.9 | 32.3 |
| Penn107 | 2022;12 | 31 | 30.4 |
| Penn145 | 2022;13 | 31 | 30.5 |
| Penn128 | 2022;13 | 31.1 | 30.2 |
| Penn144 | 2022;13 | 31.1 | 31.6 |
| Penn64 | 2022;11 | 31.3 | 31.2 |
| Penn273 | 2022;14 | 31.3 | 33 |
| Penn30 | 2022;10 | 31.4 | 30.2 |
| Penn350 | 2022;15 | 31.6 | 30.3 |
| Penn213 | 2022;14 | 31.6 | 31 |
| Penn240 | 2022;14 | 31.7 | 30.8 |
| Penn345 | 2022;15 | 31.8 | 33.6 |
| Penn7 | 2022;10 | 32 | 31.1 |
| Penn168 | 2022;13 | 32 | 33.2 |
| Penn171 | 2022;13 | 32.2 | 30.6 |
| Penn279 | 2022;14 | 32.2 | 31.1 |
| Penn260 | 2022;14 | 32.2 | 31.4 |
| Penn251 | 2022;14 | 32.2 | 31.7 |
| Penn347 | 2022;15 | 32.4 | 31.1 |
| Penn265 | 2022;14 | 32.6 | 31.3 |
| Penn181 | 2022;13 | 32.6 | 32 |
| Penn209 | 2022;14 | 32.8 | 32 |
| Penn262 | 2022;14 | 33.1 | 31.8 |
| Penn365 | 2022;15 | 33.1 | 32.2 |
| Penn337 | 2022;15 | 33.1 | 34.8 |
| Penn15 | 2022;10 | 33.2 | 32 |
| Penn26 | 2022;10 | 33.2 | 32 |
| Penn207 | 2022;14 | 33.2 | 32 |
| Penn368 | 2022;15 | 33.4 | 32.4 |
| Penn33 | 2022;10 | 33.4 | 32.7 |

| | | | |
|---|---|---|---|
| Penn3 | 2022;10 | 33.5 | 31.5 |
| Penn249 | 2022;14 | 33.5 | 32.2 |
| Penn83 | 2022;12 | 33.6 | 32.1 |
| Penn43 | 2022;11 | 33.6 | 33.1 |
| Penn59 | 2022;11 | 33.7 | 32.5 |
| Penn196 | 2022;13 | 33.7 | 32.8 |
| Penn156 | 2022;13 | 33.8 | 31.9 |
| Penn254 | 2022;14 | 33.9 | 33.2 |
| Penn52 | 2022;11 | 34 | 32.6 |
| Penn340 | 2022;15 | 34 | 32.7 |
| Penn371 | 2022;15 | 34.2 | 32.9 |
| Penn55 | 2022;11 | 34.2 | 33.2 |
| Penn191 | 2022;13 | 34.2 | 34.6 |
| Penn60 | 2022;11 | 34.3 | 32.8 |
| Penn108 | 2022;12 | 34.4 | 33.3 |
| Penn11 | 2022;10 | 34.6 | 33.1 |
| Penn95 | 2022;12 | 34.9 | 33.8 |
| Penn124 | 2022;12 | 35 | 33.8 |
| Penn184 | 2022;13 | 35 | 33.9 |
| Penn281 | 2022;15 | 35 | 34.6 |
| Penn218 | 2022;14 | 35.1 | 34.1 |
| Penn101 | 2022;12 | 35.2 | 34.3 |
| Penn370 | 2022;15 | 35.5 | 34.4 |
| Penn19 | 2022;10 | 35.6 | 33.3 |
| Penn278 | 2022;14 | 35.6 | 34.3 |
| Penn318 | 2022;15 | 35.6 | 34.4 |
| Penn364 | 2022;15 | 35.7 | 33.7 |
| Penn13 | 2022;10 | 35.7 | 34.5 |
| Penn45 | 2022;11 | 35.7 | 35.4 |
| Penn25 | 2022;10 | 35.7 | 36 |
| Penn286 | 2022;15 | 35.8 | 34.4 |
| Penn54 | 2022;11 | 35.9 | 33.7 |
| Penn211 | 2022;14 | 36.3 | 34.3 |
| Penn148 | 2022;13 | 36.6 | 35.3 |
| Penn28 | 2022;10 | 37.5 | 33.2 |
| Penn297 | 2022;15 | 38.1 | 34.7 |
| MSK 46 | 2022;13 | 30.05 | 29.5 |
| MSK 56 | 2022;13 | 33 | 30.45 |
| MSK 53 | 2022;13 | 33.33 | 31.86 |
| MSK 47 | 2022;13 | 33.42 | 31.29 |
| MSK 42 | 2022;12 | 34.1 | 32.84 |
| MSK 44 | 2022;13 | 35.65 | 34.15 |
| MSK 74 | 2022;14 | 36.79 | 35.98 |

| | | | |
|---|---|---|---|
| MSK 36 | 2022;11 | 37.3 | 34.88 |
| MSK 63 | 2022;14 | 37.64 | 36.27 |
| WU328 | 2022;15 | 15.71 | 15.59 |
| WU104 | 2022;12 | 15.78 | 15.93 |
| WU311 | 2022;14 | 15.81 | 15.88 |
| WU143 | 2022;12 | 15.87 | 16.21 |
| WU160 | 2022;13 | 15.92 | 16.3 |
| WU308 | 2022;14 | 15.94 | 16.34 |
| WU272 | 2022;14 | 15.95 | 16.26 |
| WU221 | 2022;13 | 16.04 | 16.64 |
| WU233 | 2022;13 | 16.05 | 16.18 |
| WU126 | 2022;12 | 16.08 | 16.35 |
| WU101 | 2022;12 | 16.28 | 16.1 |
| WU291 | 2022;14 | 16.47 | 16.34 |
| WU140 | 2022;12 | 16.66 | 17.03 |
| WU240 | 2022;13 | 16.67 | 16.82 |
| WU177 | 2022;13 | 16.82 | 22.8 |
| WU307 | 2022;14 | 16.92 | 17.21 |
| WU318 | 2022;14 | 16.94 | 17.21 |
| WU334 | 2022;15 | 16.95 | 17.28 |
| WU222 | 2022;13 | 16.98 | 17.59 |
| WU170 | 2022;13 | 17.1 | 23.03 |
| WU310 | 2022;14 | 17.11 | 22.29 |
| WU108 | 2022;12 | 17.15 | 17.5 |
| WU168 | 2022;13 | 17.2 | 17.47 |
| WU274 | 2022;14 | 17.24 | 21.33 |
| WU79 | 2022;11 | 17.28 | 17.15 |
| WU292 | 2022;14 | 17.38 | 22.28 |
| WU329 | 2022;15 | 17.4 | 17.3 |
| WU257 | 2022;13 | 17.55 | 17.92 |
| WU209 | 2022;13 | 17.58 | 17.94 |
| WU335 | 2022;15 | 17.61 | 21.42 |
| WU40 | 2022;10 | 17.63 | 18.04 |
| WU173 | 2022;13 | 17.64 | 18 |
| WU62 | 2022;10 | 17.66 | 16.9 |
| WU275 | 2022;14 | 17.67 | 18 |
| WU304 | 2022;14 | 17.69 | 17.98 |
| WU251 | 2022;13 | 17.78 | 17.95 |
| WU188 | 2022;13 | 17.79 | 23.19 |
| WU244 | 2022;13 | 17.87 | 18.08 |
| WU303 | 2022;14 | 17.87 | 18.24 |
| WU161 | 2022;13 | 17.91 | 23.52 |
| WU121 | 2022;12 | 17.97 | 17.53 |

| | | | |
|---|---|---|---|
| WU131 | 2022;12 | 17.99 | 18.32 |
| WU137 | 2022;12 | 17.99 | 18.76 |
| WU184 | 2022;13 | 18.08 | 18.78 |
| WU255 | 2022;13 | 18.08 | 23.05 |
| WU135 | 2022;12 | 18.09 | 18.54 |
| WU106 | 2022;12 | 18.12 | 18.64 |
| WU158 | 2022;13 | 18.16 | 18.59 |
| WU39 | 2022;10 | 18.17 | 18.62 |
| WU242 | 2022;13 | 18.18 | 18.44 |
| WU283 | 2022;14 | 18.19 | 23.61 |
| WU288 | 2022;14 | 18.21 | 18.35 |
| WU256 | 2022;13 | 18.21 | 18.47 |
| WU208 | 2022;13 | 18.27 | 18.81 |
| WU336 | 2022;15 | 18.28 | 21.98 |
| WU204 | 2022;13 | 18.28 | 24.17 |
| WU293 | 2022;14 | 18.41 | 18.64 |
| WU290 | 2022;14 | 18.43 | 23.7 |
| WU166 | 2022;13 | 18.46 | 18.81 |
| WU128 | 2022;12 | 18.49 | 18.75 |
| WU174 | 2022;13 | 18.5 | 18.68 |
| WU314 | 2022;14 | 18.58 | 18.22 |
| WU109 | 2022;12 | 18.58 | 19.09 |
| WU241 | 2022;13 | 18.69 | 18.96 |
| WU205 | 2022;13 | 18.7 | 19.42 |
| WU289 | 2022;14 | 18.75 | 23.85 |
| WU116 | 2022;12 | 18.78 | 19.12 |
| WU146 | 2022;12 | 18.84 | 19.45 |
| WU96 | 2022;12 | 18.94 | 18.88 |
| WU309 | 2022;14 | 19.01 | 19.35 |
| WU191 | 2022;13 | 19.04 | 19.65 |
| WU119 | 2022;12 | 19.06 | 18.79 |
| WU189 | 2022;13 | 19.13 | 19.98 |
| WU172 | 2022;13 | 19.24 | 19.8 |
| WU294 | 2022;14 | 19.27 | 19.29 |
| WU254 | 2022;13 | 19.33 | 19.4 |
| WU35 | 2022;10 | 19.39 | 20.22 |
| WU136 | 2022;12 | 19.43 | 20.05 |
| WU207 | 2022;13 | 19.5 | 20.07 |
| WU100 | 2022;12 | 19.52 | 19.3 |
| WU107 | 2022;12 | 19.52 | 20.18 |
| WU95 | 2022;12 | 19.6 | 19.26 |
| WU164 | 2022;13 | 19.62 | 19.67 |
| WU142 | 2022;12 | 19.62 | 19.73 |

| | | | |
|---|---|---|---|
| WU159 | 2022;13 | 19.7 | 20.05 |
| WU333 | 2022;15 | 19.82 | 19.91 |
| WU111 | 2022;12 | 19.84 | 20.1 |
| WU200 | 2022;13 | 19.9 | 24.02 |
| WU129 | 2022;12 | 19.93 | 20.24 |
| WU10 | 2022;10 | 19.94 | 20.11 |
| WU130 | 2022;12 | 20.03 | 20.32 |
| WU132 | 2022;12 | 20.05 | 20.06 |
| WU124 | 2022;12 | 20.06 | 20.45 |
| WU259 | 2022;13 | 20.11 | 20.17 |
| WU9 | 2022;10 | 20.32 | 20.56 |
| WU26 | 2022;10 | 20.32 | 20.79 |
| WU211 | 2022;13 | 20.42 | 26.04 |
| WU155 | 2022;13 | 20.43 | 20.06 |
| WU185 | 2022;13 | 20.43 | 20.89 |
| WU202 | 2022;13 | 20.49 | 20.92 |
| WU134 | 2022;12 | 20.64 | 20.66 |
| WU231 | 2022;13 | 20.69 | 21.44 |
| WU43 | 2022;10 | 20.8 | 21.06 |
| WU84 | 2022;11 | 20.84 | 21.48 |
| WU286 | 2022;14 | 20.85 | 26.25 |
| WU192 | 2022;13 | 20.94 | 26.37 |
| WU178 | 2022;13 | 21.09 | 21.5 |
| WU316 | 2022;14 | 21.12 | 25.4 |
| WU93 | 2022;11 | 21.2 | 22.15 |
| WU65 | 2022;11 | 21.32 | 22.13 |
| WU105 | 2022;12 | 21.42 | 22.27 |
| WU150 | 2022;13 | 21.46 | 21.78 |
| WU162 | 2022;13 | 21.48 | 21.97 |
| WU56 | 2022;10 | 21.5 | 22 |
| WU268 | 2022;13 | 21.51 | 20.95 |
| WU203 | 2022;13 | 21.55 | 21.9 |
| WU180 | 2022;13 | 21.56 | 21.69 |
| WU112 | 2022;12 | 21.79 | 22.17 |
| WU144 | 2022;12 | 21.87 | 21.92 |
| WU245 | 2022;13 | 21.98 | 21.87 |
| WU88 | 2022;11 | 21.98 | 22.24 |
| WU37 | 2022;10 | 21.98 | 22.62 |
| WU213 | 2022;13 | 22.09 | 27.7 |
| WU49 | 2022;10 | 22.15 | 21.52 |
| WU36 | 2022;10 | 22.18 | 22.56 |
| WU19 | 2022;10 | 22.21 | 22.75 |
| WU47 | 2022;10 | 22.23 | 22.62 |

| | | | |
|---|---|---|---|
| WU25 | 2022;10 | 22.26 | 22.76 |
| WU223 | 2022;13 | 22.4 | 22.56 |
| WU276 | 2022;14 | 22.41 | 22.24 |
| WU301 | 2022;14 | 22.49 | 22.5 |
| WU77 | 2022;11 | 22.5 | 23.07 |
| WU11 | 2022;10 | 22.51 | 22.86 |
| WU157 | 2022;13 | 22.53 | 22.63 |
| WU305 | 2022;14 | 22.53 | 22.78 |
| WU21 | 2022;10 | 22.55 | 22.81 |
| WU28 | 2022;10 | 22.61 | 22.6 |
| WU90 | 2022;11 | 22.62 | 22.28 |
| WU83 | 2022;11 | 22.67 | 22.37 |
| WU210 | 2022;13 | 22.83 | 23.34 |
| WU277 | 2022;14 | 22.84 | 23.2 |
| WU153 | 2022;13 | 22.85 | 23.11 |
| WU298 | 2022;14 | 22.9 | 27.83 |
| WU287 | 2022;14 | 22.95 | 23 |
| WU113 | 2022;12 | 22.96 | 23.31 |
| WU267 | 2022;13 | 22.98 | 23.33 |
| WU23 | 2022;10 | 22.99 | 23.46 |
| WU31 | 2022;10 | 23.09 | 22.59 |
| WU187 | 2022;13 | 23.19 | 24.07 |
| WU281 | 2022;14 | 23.26 | 23.4 |
| WU260 | 2022;13 | 23.43 | 27.82 |
| WU16 | 2022;10 | 23.45 | 23.01 |
| WU133 | 2022;12 | 23.49 | 23.5 |
| WU332 | 2022;15 | 23.57 | 23.34 |
| WU271 | 2022;13 | 23.59 | 23.95 |
| WU312 | 2022;14 | 23.73 | 23.12 |
| WU325 | 2022;15 | 23.76 | 23.71 |
| WU176 | 2022;13 | 23.8 | 24.18 |
| WU186 | 2022;13 | 23.8 | 24.95 |
| WU215 | 2022;13 | 23.85 | 24.16 |
| WU284 | 2022;14 | 23.92 | 28.66 |
| WU42 | 2022;10 | 23.96 | 24.59 |
| WU85 | 2022;11 | 24 | 23.87 |
| WU139 | 2022;12 | 24.02 | 24.33 |
| WU238 | 2022;13 | 24.07 | 28.17 |
| WU66 | 2022;11 | 24.18 | 24.36 |
| WU145 | 2022;12 | 24.18 | 24.45 |
| WU248 | 2022;13 | 24.22 | 24.28 |
| WU225 | 2022;13 | 24.29 | 24.63 |
| WU127 | 2022;12 | 24.51 | 24.28 |

| | | | |
|---|---|---|---|
| WU69 | 2022;11 | 24.53 | 24.8 |
| WU149 | 2022;13 | 24.54 | 24.74 |
| WU190 | 2022;13 | 24.89 | 25.74 |
| WU147 | 2022;13 | 24.91 | 24.68 |
| WU97 | 2022;12 | 24.94 | 24.72 |
| WU273 | 2022;14 | 24.95 | 25.13 |
| WU123 | 2022;12 | 25.08 | 25.31 |
| WU317 | 2022;14 | 25.17 | 25.35 |
| WU229 | 2022;13 | 25.24 | 25.23 |
| WU262 | 2022;13 | 25.3 | 24.67 |
| WU297 | 2022;14 | 25.3 | 24.85 |
| WU70 | 2022;11 | 25.36 | 26.42 |
| WU169 | 2022;13 | 25.44 | 25.08 |
| WU12 | 2022;10 | 25.52 | 25.48 |
| WU250 | 2022;13 | 25.55 | 25.73 |
| WU270 | 2022;13 | 25.55 | 25.92 |
| WU72 | 2022;11 | 25.66 | 25.9 |
| WU125 | 2022;12 | 25.68 | 25.75 |
| WU181 | 2022;13 | 25.69 | 25.85 |
| WU141 | 2022;12 | 25.74 | 25.78 |
| WU330 | 2022;15 | 25.89 | 25.46 |
| WU234 | 2022;13 | 25.97 | 26.19 |
| WU1 | 2022;10 | 26.02 | 25.71 |
| WU27 | 2022;10 | 26.22 | 26.63 |
| WU151 | 2022;13 | 26.24 | 26.12 |
| WU179 | 2022;13 | 26.32 | 26.45 |
| WU263 | 2022;13 | 26.35 | 25.8 |
| WU264 | 2022;13 | 26.59 | 26.32 |
| WU80 | 2022;11 | 26.79 | 26.55 |
| WU114 | 2022;12 | 26.8 | 26.92 |
| WU302 | 2022;14 | 26.87 | 26.78 |
| WU232 | 2022;13 | 26.93 | 26.88 |
| WU86 | 2022;11 | 27.18 | 27.79 |
| WU247 | 2022;13 | 27.26 | 27.66 |
| WU282 | 2022;14 | 27.29 | 27.53 |
| WU246 | 2022;13 | 27.34 | 27.7 |
| WU94 | 2022;12 | 27.35 | 28.01 |
| WU249 | 2022;13 | 27.36 | 27.48 |
| WU82 | 2022;11 | 27.37 | 26.65 |
| WU285 | 2022;14 | 27.58 | 32.4 |
| WU206 | 2022;13 | 27.59 | 28.48 |
| WU22 | 2022;10 | 27.68 | 28.34 |
| WU224 | 2022;13 | 27.95 | 28.5 |

| | | | |
|---|---|---|---|
| WU154 | 2022;13 | 28.16 | 28.29 |
| WU269 | 2022;13 | 28.34 | 27.74 |
| WU323 | 2022;15 | 28.61 | 28.31 |
| WU217 | 2022;13 | 28.63 | 29.2 |
| WU102 | 2022;12 | 28.72 | 29.26 |
| WU98 | 2022;12 | 28.78 | 28.24 |
| WU280 | 2022;14 | 28.82 | 29.01 |
| WU41 | 2022;10 | 29.01 | 29.48 |
| WU163 | 2022;13 | 29.05 | 29.48 |
| WU50 | 2022;10 | 29.08 | 29.66 |
| WU194 | 2022;13 | 29.44 | 30.68 |
| WU258 | 2022;13 | 29.46 | 29.96 |
| WU52 | 2022;10 | 29.69 | 27.92 |
| WU5 | 2022;10 | 29.78 | 28.76 |
| WU218 | 2022;13 | 29.91 | 33.62 |
| WU193 | 2022;13 | 29.95 | 30.99 |
| WU89 | 2022;11 | 29.96 | 29.47 |
| WU54 | 2022;10 | 30.2 | 29.3 |
| WU138 | 2022;12 | 30.29 | 31.1 |
| WU324 | 2022;15 | 30.35 | 29.55 |
| WU253 | 2022;13 | 30.54 | 30.47 |
| WU117 | 2022;12 | 30.56 | 31.09 |
| WU57 | 2022;10 | 30.66 | 30.05 |
| WU326 | 2022;15 | 31.02 | 30.73 |
| WU199 | 2022;13 | 31.12 | 30.88 |
| WU152 | 2022;13 | 31.18 | 31.88 |
| WU236 | 2022;13 | 31.26 | 32.07 |
| WU212 | 2022;13 | 31.28 | 35.6 |
| WU300 | 2022;14 | 31.68 | 32.62 |
| WU76 | 2022;11 | 31.68 | 32.69 |
| WU115 | 2022;12 | 31.74 | 32.47 |
| WU331 | 2022;15 | 31.88 | 30.62 |
| WU110 | 2022;12 | 31.9 | 32.63 |
| WU148 | 2022;13 | 32.44 | 31.44 |
| WU118 | 2022;12 | 32.6 | 33.51 |
| WU235 | 2022;13 | 32.6 | 33.92 |
| WU87 | 2022;11 | 32.91 | 34.2 |
| WU295 | 2022;14 | 32.95 | 34.1 |
| WU279 | 2022;14 | 33.13 | 34.59 |
| WU296 | 2022;14 | 33.17 | 32.6 |
| WU175 | 2022;13 | 33.36 | 35.16 |
| WU227 | 2022;13 | 33.39 | 34.83 |
| WU237 | 2022;13 | 33.44 | 34.78 |

| | | | |
|---|---|---|---|
| WU34 | 2022;10 | 33.5 | 31.8 |
| WU17 | 2022;10 | 33.57 | 31.72 |
| WU78 | 2022;11 | 33.61 | 34.84 |
| WU92 | 2022;11 | 33.64 | 34.74 |
| WU99 | 2022;12 | 33.77 | 32.51 |
| WU75 | 2022;11 | 33.85 | 34.94 |
| WU30 | 2022;10 | 33.96 | 32.52 |
| WU220 | 2022;13 | 34.03 | 33 |
| WU195 | 2022;13 | 34.03 | 35.37 |
| WU44 | 2022;10 | 34.06 | 32.62 |
| WU167 | 2022;13 | 34.08 | 35.59 |
| WU91 | 2022;11 | 34.15 | 33.29 |
| WU321 | 2022;15 | 34.2 | 32.99 |
| WU261 | 2022;13 | 34.2 | 35.87 |
| WU197 | 2022;13 | 34.21 | 36.66 |
| WU120 | 2022;12 | 34.26 | 35.15 |
| WU196 | 2022;13 | 34.35 | 36.01 |
| WU7 | 2022;10 | 34.84 | 32.98 |
| WU278 | 2022;14 | 34.87 | 37.88 |
| WU3 | 2022;10 | 35.03 | 32.38 |
| WU156 | 2022;13 | 35.11 | 36.3 |
| WU68 | 2022;11 | 35.21 | 35.1 |
| WU73 | 2022;11 | 35.22 | 32.46 |
| WU320 | 2022;15 | 35.33 | 33.82 |
| WU306 | 2022;14 | 35.33 | 38.1 |
| WU2 | 2022;10 | 35.39 | 32.83 |
| WU230 | 2022;13 | 35.51 | 33.3 |
| WU228 | 2022;13 | 35.55 | 37.83 |
| WU327 | 2022;15 | 35.62 | 33.77 |
| WU20 | 2022;10 | 35.66 | 38.74 |
| WU198 | 2022;13 | 35.77 | 34.81 |
| WU319 | 2022;15 | 35.88 | 33.97 |
| WU6 | 2022;10 | 36 | 33.1 |
| WU183 | 2022;13 | 36.03 | 35.82 |
| WU55 | 2022;10 | 36.1 | 33.94 |
| WU58 | 2022;10 | 36.21 | 34.19 |
| WU74 | 2022;11 | 36.71 | 37.55 |
| WU8 | 2022;10 | 37.09 | 34.17 |
| WU59 | 2022;10 | 38.14 | 33.68 |
| YNHH-1237 | 2022;15 | 14.5 | 19.2 |
| YNHH-0975 | 2022;14 | 15.2 | 15 |
| YNHH-0998 | 2022;14 | 15.4 | 20.1 |
| YNHH-0970 | 2022;14 | 15.6 | 15.7 |

| | | | |
|---|---|---|---|
| YNHH-1737 | 2022;15 | 15.7 | 15.7 |
| YNHH-0764 | 2022;14 | 15.7 | 20 |
| YNHH-1526 | 2022;15 | 15.8 | 15.5 |
| YNHH-1653 | 2022;15 | 15.9 | 15.6 |
| YNHH-0819 | 2022;14 | 15.9 | 16 |
| YNHH-1107 | 2022;14 | 16.3 | 16 |
| YNHH-1018 | 2022;14 | 16.3 | 16.5 |
| YNHH-0974 | 2022;14 | 16.4 | 16.1 |
| YNHH-1320 | 2022;15 | 16.5 | 16.2 |
| YNHH-1314 | 2022;15 | 16.5 | 16.6 |
| YNHH-1159 | 2022;14 | 16.6 | 16.5 |
| YNHH-1492 | 2022;15 | 16.6 | 21 |
| YNHH-0018 | 2022;10 | 16.7 | 16.5 |
| YNHH-1097 | 2022;14 | 16.7 | 16.6 |
| YNHH-1311 | 2022;15 | 16.7 | 16.7 |
| YNHH-1246 | 2022;15 | 16.7 | 21.1 |
| YNHH-1135 | 2022;14 | 16.8 | 16.8 |
| YNHH-1360 | 2022;15 | 16.8 | 16.8 |
| YNHH-1290 | 2022;15 | 16.8 | 16.9 |
| YNHH-1250 | 2022;15 | 16.9 | 16.2 |
| YNHH-0788 | 2022;14 | 16.9 | 16.6 |
| YNHH-1009 | 2022;14 | 16.9 | 16.7 |
| YNHH-1329 | 2022;15 | 16.9 | 16.8 |
| YNHH-1403 | 2022;15 | 16.99 | 20.61 |
| YNHH-0792 | 2022;14 | 17 | 16.7 |
| YNHH-1209 | 2022;15 | 17 | 17 |
| YNHH-1694 | 2022;15 | 17 | 17 |
| YNHH-0340 | 2022;12 | 17.1 | 17 |
| YNHH-1498 | 2022;15 | 17.1 | 17 |
| YNHH-0510 | 2022;13 | 17.1 | 17.1 |
| YNHH-1052 | 2022;14 | 17.1 | 17.1 |
| YNHH-1276 | 2022;15 | 17.1 | 17.3 |
| YNHH-1679 | 2022;15 | 17.1 | 21.1 |
| YNHH-1540 | 2022;15 | 17.1 | 21.2 |
| YNHH-1054 | 2022;14 | 17.1 | 21.8 |
| YNHH-1181 | 2022;15 | 17.2 | 17 |
| YNHH-1383 | 2022;15 | 17.2 | 17.3 |
| YNHH-0999 | 2022;14 | 17.2 | 17.5 |
| YNHH-0984 | 2022;14 | 17.2 | 21.9 |
| YNHH-1435 | 2022;15 | 17.21 | 16.99 |
| YNHH-0451 | 2022;13 | 17.3 | 16.9 |
| YNHH-0315 | 2022;12 | 17.3 | 17 |
| YNHH-1056 | 2022;14 | 17.3 | 17.1 |

| | | | |
|---|---|---|---|
| YNHH-1462 | 2022;15 | 17.3 | 17.1 |
| YNHH-1652 | 2022;15 | 17.3 | 17.3 |
| YNHH-1426 | 2022;15 | 17.3 | 21.8 |
| YNHH-0891 | 2022;14 | 17.4 | 17 |
| YNHH-0331 | 2022;12 | 17.4 | 17.3 |
| YNHH-1075 | 2022;14 | 17.4 | 17.3 |
| YNHH-1519 | 2022;15 | 17.4 | 17.6 |
| YNHH-1444 | 2022;15 | 17.5 | 17.1 |
| YNHH-0548 | 2022;13 | 17.5 | 17.2 |
| YNHH-1014 | 2022;14 | 17.5 | 17.2 |
| YNHH-1277 | 2022;15 | 17.5 | 17.3 |
| YNHH-1117 | 2022;14 | 17.5 | 17.4 |
| YNHH-1560 | 2022;15 | 17.5 | 17.4 |
| YNHH-1509 | 2022;15 | 17.5 | 17.5 |
| YNHH-1263 | 2022;15 | 17.5 | 17.6 |
| YNHH-1235 | 2022;15 | 17.5 | 22.2 |
| YNHH-1389 | 2022;15 | 17.5 | 22.4 |
| YNHH-1291 | 2022;15 | 17.6 | 17.3 |
| YNHH-0855 | 2022;14 | 17.6 | 17.4 |
| YNHH-1481 | 2022;15 | 17.6 | 17.7 |
| YNHH-1385 | 2022;15 | 17.6 | 22.5 |
| YNHH-1076 | 2022;14 | 17.7 | 17.5 |
| YNHH-0943 | 2022;14 | 17.7 | 17.7 |
| YNHH-1200 | 2022;15 | 17.7 | 21.1 |
| YNHH-1398 | 2022;15 | 17.7 | 22.4 |
| YNHH-1434 | 2022;15 | 17.8 | 17.4 |
| YNHH-1085 | 2022;14 | 17.9 | 17.6 |
| YNHH-0884 | 2022;14 | 17.9 | 17.8 |
| YNHH-1244 | 2022;15 | 17.9 | 17.8 |
| YNHH-1109 | 2022;14 | 17.9 | 22.6 |
| YNHH-1174 | 2022;15 | 17.9 | 22.8 |
| YNHH-1373 | 2022;15 | 17.97 | 18.01 |
| YNHH-0515 | 2022;13 | 18 | 17.8 |
| YNHH-0967 | 2022;14 | 18 | 17.8 |
| YNHH-1272 | 2022;15 | 18 | 17.8 |
| YNHH-1275 | 2022;15 | 18 | 17.8 |
| YNHH-0821 | 2022;14 | 18 | 17.9 |
| YNHH-0951 | 2022;14 | 18 | 17.9 |
| YNHH-1577 | 2022;15 | 18 | 18.2 |
| YNHH-1415 | 2022;15 | 18.05 | 22.27 |
| YNHH-0003 | 2022;12 | 18.1 | 17.6 |
| YNHH-0329 | 2022;12 | 18.1 | 18 |
| YNHH-1094 | 2022;14 | 18.1 | 18 |

| | | | |
|---|---|---|---|
| YNHH-1179 | 2022;15 | 18.1 | 18 |
| YNHH-1365 | 2022;15 | 18.1 | 18.1 |
| YNHH-1622 | 2022;15 | 18.1 | 21.9 |
| YNHH-1626 | 2022;15 | 18.2 | 17.7 |
| YNHH-0980 | 2022;14 | 18.2 | 18 |
| YNHH-1057 | 2022;14 | 18.2 | 18 |
| YNHH-1025 | 2022;14 | 18.2 | 18.2 |
| YNHH-1264 | 2022;15 | 18.2 | 18.2 |
| YNHH-1596 | 2022;15 | 18.2 | 18.2 |
| YNHH-0323 | 2022;12 | 18.2 | 22.5 |
| YNHH-1470 | 2022;15 | 18.2 | 22.5 |
| YNHH-0962 | 2022;14 | 18.3 | 18.1 |
| YNHH-1271 | 2022;15 | 18.3 | 18.1 |
| YNHH-1671 | 2022;15 | 18.3 | 18.1 |
| YNHH-0901 | 2022;14 | 18.3 | 18.2 |
| YNHH-0981 | 2022;14 | 18.3 | 18.2 |
| YNHH-1388 | 2022;15 | 18.3 | 18.3 |
| YNHH-1309 | 2022;15 | 18.3 | 18.4 |
| YNHH-0898 | 2022;14 | 18.4 | 18 |
| YNHH-1571 | 2022;15 | 18.4 | 18.3 |
| YNHH-0994 | 2022;14 | 18.4 | 18.4 |
| YNHH-1128 | 2022;14 | 18.4 | 18.5 |
| YNHH-1659 | 2022;15 | 18.4 | 18.5 |
| YNHH-1108 | 2022;14 | 18.4 | 22.2 |
| YNHH-1650 | 2022;15 | 18.4 | 22.5 |
| YNHH-1132 | 2022;14 | 18.5 | 18.4 |
| YNHH-1457 | 2022;15 | 18.5 | 18.4 |
| YNHH-1422 | 2022;15 | 18.5 | 18.5 |
| YNHH-1399 | 2022;15 | 18.5 | 22.5 |
| YNHH-1024 | 2022;14 | 18.5 | 23.3 |
| YNHH-1003 | 2022;14 | 18.6 | 18.4 |
| YNHH-1040 | 2022;14 | 18.6 | 18.5 |
| YNHH-1224 | 2022;15 | 18.6 | 19.1 |
| YNHH-1657 | 2022;15 | 18.6 | 22.8 |
| YNHH-1039 | 2022;14 | 18.6 | 23.2 |
| YNHH-1062 | 2022;14 | 18.7 | 18.4 |
| YNHH-0509 | 2022;13 | 18.7 | 18.5 |
| YNHH-1238 | 2022;15 | 18.7 | 18.7 |
| YNHH-0601 | 2022;13 | 18.7 | 18.8 |
| YNHH-1078 | 2022;14 | 18.7 | 22.8 |
| YNHH-1705 | 2022;15 | 18.8 | 18.5 |
| YNHH-1038 | 2022;14 | 18.8 | 18.6 |
| YNHH-1064 | 2022;14 | 18.8 | 18.6 |

| | | | |
|---|---|---|---|
| YNHH-1476 | 2022;15 | 18.8 | 18.6 |
| YNHH-1578 | 2022;15 | 18.8 | 18.6 |
| YNHH-1603 | 2022;15 | 18.8 | 18.6 |
| YNHH-1086 | 2022;14 | 18.8 | 18.7 |
| YNHH-1105 | 2022;14 | 18.8 | 18.7 |
| YNHH-1212 | 2022;15 | 18.8 | 18.7 |
| YNHH-1579 | 2022;15 | 18.8 | 18.7 |
| YNHH-1714 | 2022;15 | 18.8 | 18.7 |
| YNHH-1742 | 2022;15 | 18.8 | 18.8 |
| YNHH-1193 | 2022;15 | 18.8 | 22.3 |
| YNHH-1304 | 2022;15 | 18.8 | 22.5 |
| YNHH-1050 | 2022;14 | 18.8 | 23.1 |
| YNHH-0799 | 2022;14 | 18.9 | 18.5 |
| YNHH-0903 | 2022;14 | 18.9 | 18.6 |
| YNHH-1497 | 2022;15 | 18.9 | 18.7 |
| YNHH-1557 | 2022;15 | 18.9 | 18.7 |
| YNHH-0937 | 2022;14 | 18.9 | 18.8 |
| YNHH-1332 | 2022;15 | 18.9 | 18.8 |
| YNHH-0867 | 2022;14 | 19 | 18.6 |
| YNHH-1297 | 2022;15 | 19 | 18.6 |
| YNHH-0987 | 2022;14 | 19 | 18.7 |
| YNHH-1144 | 2022;14 | 19 | 18.7 |
| YNHH-1157 | 2022;14 | 19 | 18.9 |
| YNHH-1493 | 2022;15 | 19 | 18.9 |
| YNHH-1611 | 2022;15 | 19 | 19 |
| YNHH-1223 | 2022;15 | 19 | 19.1 |
| YNHH-1525 | 2022;15 | 19 | 19.1 |
| YNHH-1573 | 2022;15 | 19 | 23 |
| YNHH-1419 | 2022;15 | 19.01 | 18.88 |
| YNHH-1701 | 2022;15 | 19.1 | 18.8 |
| YNHH-0852 | 2022;14 | 19.1 | 18.9 |
| YNHH-1019 | 2022;14 | 19.1 | 18.9 |
| YNHH-0849 | 2022;14 | 19.1 | 19 |
| YNHH-1416 | 2022;15 | 19.1 | 21.5 |
| YNHH-0972 | 2022;14 | 19.2 | 18.8 |
| YNHH-1177 | 2022;15 | 19.2 | 19 |
| YNHH-1068 | 2022;14 | 19.2 | 19.1 |
| YNHH-1319 | 2022;15 | 19.2 | 23.1 |
| YNHH-1341 | 2022;15 | 19.2 | 23.9 |
| YNHH-1592 | 2022;15 | 19.3 | 19 |
| YNHH-1654 | 2022;15 | 19.3 | 19 |
| YNHH-1111 | 2022;14 | 19.3 | 19.2 |
| YNHH-0569 | 2022;13 | 19.3 | 19.3 |

| | | | |
|---|---|---|---|
| YNHH-1127 | 2022;14 | 19.3 | 19.3 |
| YNHH-1080 | 2022;14 | 19.3 | 19.4 |
| YNHH-1321 | 2022;15 | 19.3 | 23.2 |
| YNHH-1621 | 2022;15 | 19.3 | 23.4 |
| YNHH-1026 | 2022;14 | 19.3 | 24.1 |
| YNHH-1055 | 2022;14 | 19.3 | 24.2 |
| YNHH-1357 | 2022;15 | 19.4 | 18.93 |
| YNHH-1255 | 2022;15 | 19.4 | 19 |
| YNHH-0888 | 2022;14 | 19.4 | 19.1 |
| YNHH-0911 | 2022;14 | 19.4 | 19.2 |
| YNHH-1351 | 2022;15 | 19.4 | 19.3 |
| YNHH-1554 | 2022;15 | 19.4 | 19.3 |
| YNHH-0594 | 2022;13 | 19.4 | 19.6 |
| YNHH-0804 | 2022;14 | 19.4 | 23.2 |
| YNHH-1523 | 2022;15 | 19.5 | 19.1 |
| YNHH-0751 | 2022;14 | 19.5 | 19.2 |
| YNHH-0912 | 2022;14 | 19.5 | 19.3 |
| YNHH-0941 | 2022;14 | 19.5 | 19.3 |
| YNHH-1139 | 2022;14 | 19.5 | 19.3 |
| YNHH-1496 | 2022;15 | 19.5 | 19.3 |
| YNHH-0567 | 2022;13 | 19.5 | 19.4 |
| YNHH-0955 | 2022;14 | 19.5 | 19.4 |
| YNHH-1013 | 2022;14 | 19.5 | 19.4 |
| YNHH-1196 | 2022;15 | 19.5 | 19.4 |
| YNHH-1570 | 2022;15 | 19.5 | 19.5 |
| YNHH-0991 | 2022;14 | 19.5 | 19.6 |
| YNHH-1335 | 2022;15 | 19.5 | 23.3 |
| YNHH-1555 | 2022;15 | 19.5 | 24 |
| YNHH-0014 | 2022;10 | 19.6 | 19.3 |
| YNHH-0921 | 2022;14 | 19.6 | 19.3 |
| YNHH-1093 | 2022;14 | 19.6 | 19.5 |
| YNHH-1289 | 2022;15 | 19.6 | 19.7 |
| YNHH-1623 | 2022;15 | 19.6 | 23.8 |
| YNHH-0904 | 2022;14 | 19.7 | 19.2 |
| YNHH-1353 | 2022;15 | 19.7 | 19.4 |
| YNHH-1230 | 2022;15 | 19.7 | 19.5 |
| YNHH-0917 | 2022;14 | 19.7 | 19.6 |
| YNHH-1000 | 2022;14 | 19.7 | 19.6 |
| YNHH-1722 | 2022;15 | 19.7 | 19.6 |
| YNHH-1002 | 2022;14 | 19.7 | 19.7 |
| YNHH-1569 | 2022;15 | 19.7 | 24 |
| YNHH-1689 | 2022;15 | 19.7 | 24 |
| YNHH-0996 | 2022;14 | 19.8 | 19.6 |

| | | | |
|---|---|---|---|
| YNHH-1220 | 2022;15 | 19.8 | 19.6 |
| YNHH-1137 | 2022;14 | 19.8 | 19.7 |
| YNHH-1675 | 2022;15 | 19.8 | 19.7 |
| YNHH-1475 | 2022;15 | 19.8 | 19.8 |
| YNHH-1455 | 2022;15 | 19.8 | 19.9 |
| YNHH-1342 | 2022;15 | 19.8 | 20.2 |
| YNHH-1740 | 2022;15 | 19.8 | 22.9 |
| YNHH-1084 | 2022;14 | 19.8 | 24.2 |
| YNHH-1605 | 2022;15 | 19.9 | 19.5 |
| YNHH-0551 | 2022;13 | 19.9 | 19.6 |
| YNHH-0940 | 2022;14 | 19.9 | 19.8 |
| YNHH-1668 | 2022;15 | 19.9 | 19.8 |
| YNHH-1120 | 2022;14 | 19.9 | 19.9 |
| YNHH-1547 | 2022;15 | 19.9 | 19.9 |
| YNHH-1393 | 2022;15 | 19.9 | 20.1 |
| YNHH-1098 | 2022;14 | 19.9 | 24 |
| YNHH-1377 | 2022;15 | 19.9 | 24.5 |
| YNHH-1713 | 2022;15 | 20 | 19.4 |
| YNHH-1292 | 2022;15 | 20 | 19.5 |
| YNHH-0324 | 2022;12 | 20 | 19.6 |
| YNHH-0924 | 2022;14 | 20 | 19.8 |
| YNHH-0978 | 2022;14 | 20 | 19.8 |
| YNHH-1047 | 2022;14 | 20 | 19.9 |
| YNHH-1171 | 2022;15 | 20 | 20 |
| YNHH-1696 | 2022;15 | 20 | 23.9 |
| YNHH-1591 | 2022;15 | 20 | 24.1 |
| YNHH-1515 | 2022;15 | 20 | 24.6 |
| YNHH-0816 | 2022;14 | 20.1 | 19.8 |
| YNHH-1649 | 2022;15 | 20.1 | 19.9 |
| YNHH-1472 | 2022;15 | 20.1 | 20.3 |
| YNHH-0011 | 2022;15 | 20.1 | 24.2 |
| YNHH-0997 | 2022;14 | 20.1 | 24.2 |
| YNHH-1715 | 2022;15 | 20.1 | 24.4 |
| YNHH-1354 | 2022;15 | 20.11 | 23.9 |
| YNHH-0918 | 2022;14 | 20.2 | 19.8 |
| YNHH-0925 | 2022;14 | 20.2 | 19.8 |
| YNHH-1138 | 2022;14 | 20.2 | 19.9 |
| YNHH-0929 | 2022;14 | 20.2 | 20 |
| YNHH-1663 | 2022;15 | 20.2 | 20 |
| YNHH-0863 | 2022;14 | 20.2 | 20.1 |
| YNHH-1150 | 2022;14 | 20.2 | 20.3 |
| YNHH-1152 | 2022;14 | 20.2 | 20.3 |
| YNHH-0801 | 2022;14 | 20.2 | 24.4 |

| | | | |
|---|---|---|---|
| YNHH-1189 | 2022;15 | 20.3 | 20.1 |
| YNHH-1678 | 2022;15 | 20.3 | 20.1 |
| YNHH-1428 | 2022;15 | 20.3 | 20.2 |
| YNHH-1741 | 2022;15 | 20.3 | 20.2 |
| YNHH-1363 | 2022;15 | 20.3 | 20.3 |
| YNHH-0890 | 2022;14 | 20.3 | 23.8 |
| YNHH-1638 | 2022;15 | 20.3 | 24.2 |
| YNHH-0843 | 2022;14 | 20.3 | 24.8 |
| YNHH-0506 | 2022;13 | 20.4 | 19.9 |
| YNHH-1628 | 2022;15 | 20.4 | 20.2 |
| YNHH-1624 | 2022;15 | 20.4 | 20.3 |
| YNHH-1412 | 2022;15 | 20.4 | 20.5 |
| YNHH-0961 | 2022;14 | 20.4 | 20.6 |
| YNHH-1401 | 2022;15 | 20.4 | 24.5 |
| YNHH-1273 | 2022;15 | 20.4 | 25.2 |
| YNHH-0790 | 2022;14 | 20.5 | 20.1 |
| YNHH-1733 | 2022;15 | 20.5 | 20.1 |
| YNHH-0820 | 2022;14 | 20.5 | 20.3 |
| YNHH-1197 | 2022;15 | 20.5 | 20.3 |
| YNHH-1698 | 2022;15 | 20.5 | 20.3 |
| YNHH-1060 | 2022;14 | 20.5 | 20.4 |
| YNHH-1326 | 2022;15 | 20.5 | 20.4 |
| YNHH-1710 | 2022;15 | 20.5 | 20.4 |
| YNHH-1051 | 2022;14 | 20.5 | 20.5 |
| YNHH-1708 | 2022;15 | 20.5 | 24.6 |
| YNHH-0541 | 2022;13 | 20.6 | 20.2 |
| YNHH-1114 | 2022;14 | 20.6 | 20.3 |
| YNHH-1527 | 2022;15 | 20.6 | 20.4 |
| YNHH-1133 | 2022;14 | 20.6 | 20.5 |
| YNHH-1684 | 2022;15 | 20.6 | 20.5 |
| YNHH-1637 | 2022;15 | 20.6 | 20.7 |
| YNHH-1344 | 2022;15 | 20.68 | 20.37 |
| YNHH-0632 | 2022;13 | 20.7 | 20.3 |
| YNHH-1065 | 2022;14 | 20.7 | 20.5 |
| YNHH-1217 | 2022;15 | 20.7 | 20.7 |
| YNHH-0558 | 2022;13 | 20.7 | 20.8 |
| YNHH-1656 | 2022;15 | 20.7 | 21 |
| YNHH-1693 | 2022;15 | 20.7 | 25.1 |
| YNHH-0325 | 2022;12 | 20.8 | 20.4 |
| YNHH-1423 | 2022;15 | 20.8 | 20.5 |
| YNHH-1556 | 2022;15 | 20.8 | 20.5 |
| YNHH-1732 | 2022;15 | 20.8 | 20.6 |
| YNHH-1118 | 2022;14 | 20.8 | 20.7 |

| | | | |
|---|---|---|---|
| YNHH-1474 | 2022;15 | 20.8 | 20.7 |
| YNHH-1278 | 2022;15 | 20.8 | 20.9 |
| YNHH-1306 | 2022;15 | 20.8 | 21.2 |
| YNHH-0963 | 2022;14 | 20.8 | 25 |
| YNHH-0969 | 2022;14 | 20.8 | 25.2 |
| YNHH-0883 | 2022;14 | 20.9 | 20.3 |
| YNHH-1697 | 2022;15 | 20.9 | 20.6 |
| YNHH-0561 | 2022;13 | 20.9 | 20.7 |
| YNHH-1242 | 2022;15 | 20.9 | 20.7 |
| YNHH-1441 | 2022;15 | 20.9 | 20.7 |
| YNHH-1074 | 2022;14 | 20.9 | 20.8 |
| YNHH-1735 | 2022;15 | 20.9 | 24.5 |
| YNHH-1655 | 2022;15 | 20.9 | 25.1 |
| YNHH-1046 | 2022;14 | 20.9 | 25.3 |
| YNHH-0877 | 2022;14 | 21 | 20.8 |
| YNHH-0960 | 2022;14 | 21 | 20.9 |
| YNHH-1012 | 2022;14 | 21 | 20.9 |
| YNHH-1149 | 2022;14 | 21 | 20.9 |
| YNHH-0973 | 2022;14 | 21 | 25.3 |
| YNHH-0334 | 2022;12 | 21.1 | 20.6 |
| YNHH-0544 | 2022;13 | 21.1 | 20.6 |
| YNHH-0668 | 2022;13 | 21.1 | 20.6 |
| YNHH-1213 | 2022;15 | 21.1 | 20.8 |
| YNHH-1648 | 2022;15 | 21.1 | 20.8 |
| YNHH-0806 | 2022;14 | 21.1 | 21 |
| YNHH-1723 | 2022;15 | 21.1 | 21.3 |
| YNHH-0833 | 2022;14 | 21.1 | 21.4 |
| YNHH-1467 | 2022;15 | 21.1 | 25.6 |
| YNHH-1349 | 2022;15 | 21.19 | 20.59 |
| YNHH-0966 | 2022;14 | 21.2 | 20.9 |
| YNHH-1307 | 2022;15 | 21.2 | 21.2 |
| YNHH-0892 | 2022;14 | 21.2 | 25.4 |
| YNHH-1728 | 2022;15 | 21.2 | 25.6 |
| YNHH-1418 | 2022;15 | 21.26 | 25.55 |
| YNHH-1207 | 2022;15 | 21.3 | 21.1 |
| YNHH-1395 | 2022;15 | 21.3 | 21.1 |
| YNHH-1647 | 2022;15 | 21.3 | 21.1 |
| YNHH-1294 | 2022;15 | 21.3 | 25.8 |
| YNHH-0319 | 2022;12 | 21.4 | 21 |
| YNHH-1079 | 2022;14 | 21.4 | 21.1 |
| YNHH-0866 | 2022;14 | 21.4 | 21.2 |
| YNHH-1143 | 2022;14 | 21.4 | 21.2 |
| YNHH-1586 | 2022;15 | 21.4 | 21.2 |

| | | | |
|---|---|---|---|
| YNHH-0367 | 2022;12 | 21.4 | 21.3 |
| YNHH-0886 | 2022;14 | 21.4 | 21.3 |
| YNHH-1088 | 2022;14 | 21.4 | 21.4 |
| YNHH-0807 | 2022;14 | 21.4 | 21.6 |
| YNHH-1028 | 2022;14 | 21.4 | 25.9 |
| YNHH-1087 | 2022;14 | 21.4 | 25.9 |
| YNHH-0879 | 2022;14 | 21.5 | 21.1 |
| YNHH-1173 | 2022;15 | 21.5 | 21.2 |
| YNHH-1191 | 2022;15 | 21.5 | 21.2 |
| YNHH-0957 | 2022;14 | 21.5 | 21.3 |
| YNHH-1205 | 2022;15 | 21.5 | 21.3 |
| YNHH-1267 | 2022;15 | 21.5 | 21.6 |
| YNHH-0811 | 2022;14 | 21.5 | 25.9 |
| YNHH-1518 | 2022;15 | 21.6 | 21.1 |
| YNHH-0947 | 2022;14 | 21.6 | 21.2 |
| YNHH-0783 | 2022;14 | 21.6 | 21.4 |
| YNHH-1145 | 2022;14 | 21.6 | 21.5 |
| YNHH-1146 | 2022;14 | 21.6 | 21.5 |
| YNHH-1687 | 2022;15 | 21.6 | 21.5 |
| YNHH-1629 | 2022;15 | 21.6 | 21.6 |
| YNHH-1350 | 2022;15 | 21.6 | 25.3 |
| YNHH-1503 | 2022;15 | 21.6 | 25.4 |
| YNHH-1544 | 2022;15 | 21.6 | 25.6 |
| YNHH-1218 | 2022;15 | 21.6 | 25.7 |
| YNHH-1588 | 2022;15 | 21.6 | 26 |
| YNHH-1511 | 2022;15 | 21.7 | 21.4 |
| YNHH-1390 | 2022;15 | 21.7 | 21.7 |
| YNHH-1280 | 2022;15 | 21.7 | 21.8 |
| YNHH-1616 | 2022;15 | 21.7 | 22.3 |
| YNHH-0893 | 2022;14 | 21.7 | 25.7 |
| YNHH-0881 | 2022;14 | 21.7 | 25.9 |
| YNHH-1413 | 2022;15 | 21.76 | 21.74 |
| YNHH-1343 | 2022;15 | 21.76 | 25.73 |
| YNHH-1359 | 2022;15 | 21.8 | 21.5 |
| YNHH-1545 | 2022;15 | 21.8 | 21.5 |
| YNHH-0578 | 2022;13 | 21.8 | 21.6 |
| YNHH-0844 | 2022;14 | 21.8 | 21.6 |
| YNHH-1001 | 2022;14 | 21.8 | 21.6 |
| YNHH-1366 | 2022;15 | 21.8 | 21.7 |
| YNHH-0948 | 2022;14 | 21.8 | 21.8 |
| YNHH-1167 | 2022;15 | 21.8 | 22.3 |
| YNHH-1730 | 2022;15 | 21.9 | 21.4 |
| YNHH-1676 | 2022;15 | 21.9 | 21.6 |

| | | | |
|---|---|---|---|
| YNHH-0829 | 2022;14 | 21.9 | 21.7 |
| YNHH-0986 | 2022;14 | 21.9 | 21.7 |
| YNHH-1147 | 2022;14 | 21.9 | 21.7 |
| YNHH-1688 | 2022;15 | 21.9 | 21.7 |
| YNHH-1008 | 2022;14 | 21.9 | 21.8 |
| YNHH-1175 | 2022;15 | 21.9 | 21.8 |
| YNHH-0846 | 2022;14 | 21.9 | 21.9 |
| YNHH-0592 | 2022;13 | 21.9 | 25.5 |
| YNHH-1130 | 2022;14 | 21.9 | 25.7 |
| YNHH-0791 | 2022;14 | 21.9 | 26.8 |
| YNHH-1186 | 2022;15 | 22 | 21.5 |
| YNHH-0854 | 2022;14 | 22 | 21.7 |
| YNHH-1699 | 2022;15 | 22 | 21.7 |
| YNHH-1214 | 2022;15 | 22 | 21.8 |
| YNHH-1017 | 2022;14 | 22.1 | 21.6 |
| YNHH-1208 | 2022;15 | 22.1 | 21.8 |
| YNHH-1005 | 2022;14 | 22.1 | 21.9 |
| YNHH-1299 | 2022;15 | 22.1 | 21.9 |
| YNHH-0798 | 2022;14 | 22.1 | 22 |
| YNHH-1006 | 2022;14 | 22.1 | 22 |
| YNHH-1627 | 2022;15 | 22.1 | 22 |
| YNHH-1116 | 2022;14 | 22.1 | 22.3 |
| YNHH-1543 | 2022;15 | 22.1 | 26.5 |
| YNHH-0559 | 2022;13 | 22.2 | 21.9 |
| YNHH-0737 | 2022;13 | 22.2 | 21.9 |
| YNHH-0875 | 2022;14 | 22.2 | 21.9 |
| YNHH-0950 | 2022;14 | 22.2 | 21.9 |
| YNHH-0926 | 2022;14 | 22.2 | 22.1 |
| YNHH-1148 | 2022;14 | 22.3 | 22.1 |
| YNHH-1362 | 2022;15 | 22.3 | 22.2 |
| YNHH-1131 | 2022;14 | 22.3 | 22.3 |
| YNHH-1615 | 2022;15 | 22.3 | 25.8 |
| YNHH-0566 | 2022;13 | 22.3 | 26.3 |
| YNHH-1437 | 2022;15 | 22.3 | 26.4 |
| YNHH-1021 | 2022;14 | 22.4 | 22.3 |
| YNHH-1331 | 2022;15 | 22.4 | 26.4 |
| YNHH-1546 | 2022;15 | 22.4 | 26.4 |
| YNHH-0337 | 2022;12 | 22.5 | 22.1 |
| YNHH-1364 | 2022;15 | 22.5 | 22.1 |
| YNHH-1376 | 2022;15 | 22.5 | 22.3 |
| YNHH-1463 | 2022;15 | 22.5 | 22.4 |
| YNHH-1682 | 2022;15 | 22.5 | 26.5 |
| YNHH-0410 | 2022;12 | 22.6 | 22.2 |

| | | | |
|---|---|---|---|
| YNHH-0514 | 2022;13 | 22.6 | 22.2 |
| YNHH-1254 | 2022;15 | 22.6 | 22.8 |
| YNHH-1216 | 2022;15 | 22.6 | 26.9 |
| YNHH-1729 | 2022;15 | 22.6 | 26.9 |
| YNHH-1394 | 2022;15 | 22.6 | 27.1 |
| YNHH-1409 | 2022;15 | 22.6 | 27.1 |
| YNHH-1429 | 2022;15 | 22.69 | 22.54 |
| YNHH-1007 | 2022;14 | 22.7 | 22.4 |
| YNHH-1015 | 2022;14 | 22.7 | 22.4 |
| YNHH-0850 | 2022;14 | 22.7 | 22.5 |
| YNHH-1257 | 2022;15 | 22.7 | 22.5 |
| YNHH-0954 | 2022;14 | 22.7 | 22.6 |
| YNHH-0923 | 2022;14 | 22.7 | 26.6 |
| YNHH-1069 | 2022;14 | 22.7 | 26.7 |
| YNHH-0894 | 2022;14 | 22.8 | 22.6 |
| YNHH-1180 | 2022;15 | 22.8 | 26 |
| YNHH-1151 | 2022;14 | 22.8 | 26.2 |
| YNHH-1229 | 2022;15 | 22.9 | 22.4 |
| YNHH-0241 | 2022;11 | 22.9 | 22.6 |
| YNHH-0865 | 2022;14 | 22.9 | 22.6 |
| YNHH-1066 | 2022;14 | 22.9 | 22.7 |
| YNHH-1558 | 2022;15 | 22.9 | 22.8 |
| YNHH-1268 | 2022;15 | 22.9 | 22.9 |
| YNHH-1664 | 2022;15 | 22.9 | 22.9 |
| YNHH-1325 | 2022;15 | 22.9 | 26.7 |
| YNHH-1530 | 2022;15 | 23 | 22.7 |
| YNHH-1576 | 2022;15 | 23 | 22.7 |
| YNHH-1691 | 2022;15 | 23 | 22.7 |
| YNHH-1282 | 2022;15 | 23 | 22.8 |
| YNHH-1617 | 2022;15 | 23 | 22.8 |
| YNHH-1731 | 2022;15 | 23 | 22.9 |
| YNHH-1563 | 2022;15 | 23 | 23 |
| YNHH-1198 | 2022;15 | 23 | 23.1 |
| YNHH-1506 | 2022;15 | 23 | 27.3 |
| YNHH-1721 | 2022;15 | 23.1 | 22.6 |
| YNHH-1270 | 2022;15 | 23.1 | 22.8 |
| YNHH-1704 | 2022;15 | 23.1 | 23.3 |
| YNHH-1700 | 2022;15 | 23.1 | 27.2 |
| YNHH-1371 | 2022;15 | 23.18 | 22.91 |
| YNHH-0549 | 2022;13 | 23.2 | 22.8 |
| YNHH-1548 | 2022;15 | 23.2 | 22.9 |
| YNHH-1142 | 2022;14 | 23.2 | 23.2 |
| YNHH-1718 | 2022;15 | 23.2 | 26.8 |

| | | | |
|---|---|---|---|
| YNHH-1397 | 2022;15 | 23.21 | 22.87 |
| YNHH-1438 | 2022;15 | 23.22 | 22.77 |
| YNHH-1156 | 2022;14 | 23.3 | 23 |
| YNHH-0245 | 2022;11 | 23.3 | 23.2 |
| YNHH-1479 | 2022;15 | 23.3 | 23.2 |
| YNHH-1379 | 2022;15 | 23.3 | 23.3 |
| YNHH-1636 | 2022;15 | 23.3 | 27.3 |
| YNHH-0936 | 2022;14 | 23.3 | 27.5 |
| YNHH-0995 | 2022;14 | 23.4 | 23.1 |
| YNHH-1265 | 2022;15 | 23.4 | 23.2 |
| YNHH-1261 | 2022;15 | 23.4 | 23.3 |
| YNHH-1425 | 2022;15 | 23.49 | 23.2 |
| YNHH-1646 | 2022;15 | 23.5 | 22.9 |
| YNHH-0016 | 2022;10 | 23.5 | 23 |
| YNHH-0870 | 2022;14 | 23.5 | 23.2 |
| YNHH-1204 | 2022;15 | 23.5 | 23.3 |
| YNHH-1233 | 2022;15 | 23.5 | 23.4 |
| YNHH-1711 | 2022;15 | 23.5 | 23.4 |
| YNHH-0553 | 2022;13 | 23.5 | 23.7 |
| YNHH-1317 | 2022;15 | 23.5 | 26.9 |
| YNHH-0328 | 2022;12 | 23.6 | 23.3 |
| YNHH-1095 | 2022;14 | 23.6 | 23.3 |
| YNHH-1172 | 2022;15 | 23.6 | 23.3 |
| YNHH-0795 | 2022;14 | 23.6 | 23.4 |
| YNHH-1210 | 2022;15 | 23.6 | 23.5 |
| YNHH-1471 | 2022;15 | 23.6 | 23.6 |
| YNHH-1507 | 2022;15 | 23.6 | 23.6 |
| YNHH-1378 | 2022;15 | 23.6 | 27.9 |
| YNHH-1301 | 2022;15 | 23.6 | 28 |
| YNHH-1100 | 2022;14 | 23.7 | 23.5 |
| YNHH-1092 | 2022;14 | 23.7 | 23.6 |
| YNHH-1141 | 2022;14 | 23.7 | 23.6 |
| YNHH-1482 | 2022;15 | 23.7 | 23.6 |
| YNHH-1374 | 2022;15 | 23.8 | 23.7 |
| YNHH-1380 | 2022;15 | 23.8 | 27.9 |
| YNHH-1347 | 2022;15 | 23.82 | 27.73 |
| YNHH-1288 | 2022;15 | 23.9 | 23.7 |
| YNHH-1310 | 2022;15 | 23.9 | 23.8 |
| YNHH-1420 | 2022;15 | 23.9 | 28.1 |
| YNHH-1607 | 2022;15 | 24 | 23.6 |
| YNHH-1717 | 2022;15 | 24 | 23.7 |
| YNHH-1487 | 2022;15 | 24 | 23.8 |
| YNHH-1036 | 2022;14 | 24 | 24.3 |

| | | | |
|---|---|---|---|
| YNHH-0922 | 2022;14 | 24 | 27.6 |
| YNHH-1285 | 2022;15 | 24 | 27.9 |
| YNHH-1339 | 2022;15 | 24.1 | 23.9 |
| YNHH-1396 | 2022;15 | 24.1 | 23.9 |
| YNHH-0780 | 2022;14 | 24.2 | 23.9 |
| YNHH-0857 | 2022;14 | 24.2 | 23.9 |
| YNHH-1361 | 2022;15 | 24.2 | 24.1 |
| YNHH-0781 | 2022;14 | 24.3 | 24 |
| YNHH-1454 | 2022;15 | 24.3 | 24 |
| YNHH-1583 | 2022;15 | 24.3 | 24 |
| YNHH-0571 | 2022;13 | 24.3 | 24.1 |
| YNHH-1165 | 2022;15 | 24.3 | 24.1 |
| YNHH-0869 | 2022;14 | 24.3 | 24.2 |
| YNHH-1119 | 2022;14 | 24.3 | 24.2 |
| YNHH-0317 | 2022;12 | 24.4 | 24 |
| YNHH-1600 | 2022;15 | 24.4 | 24.2 |
| YNHH-1283 | 2022;15 | 24.4 | 24.3 |
| YNHH-0101 | 2022;10 | 24.5 | 24.1 |
| YNHH-1077 | 2022;14 | 24.5 | 24.1 |
| YNHH-0897 | 2022;14 | 24.5 | 24.2 |
| YNHH-1034 | 2022;14 | 24.5 | 24.3 |
| YNHH-1053 | 2022;14 | 24.5 | 24.3 |
| YNHH-1161 | 2022;15 | 24.5 | 24.4 |
| YNHH-1176 | 2022;15 | 24.5 | 24.4 |
| YNHH-1599 | 2022;15 | 24.5 | 28.4 |
| YNHH-1447 | 2022;15 | 24.52 | 24.34 |
| YNHH-0868 | 2022;14 | 24.6 | 24.3 |
| YNHH-1081 | 2022;14 | 24.6 | 24.4 |
| YNHH-1440 | 2022;15 | 24.6 | 24.4 |
| YNHH-1032 | 2022;14 | 24.6 | 24.5 |
| YNHH-1501 | 2022;15 | 24.6 | 24.6 |
| YNHH-1124 | 2022;14 | 24.6 | 28.2 |
| YNHH-0956 | 2022;14 | 24.7 | 24.3 |
| YNHH-1011 | 2022;14 | 24.7 | 24.7 |
| YNHH-1201 | 2022;15 | 24.7 | 24.7 |
| YNHH-1202 | 2022;15 | 24.7 | 24.7 |
| YNHH-1608 | 2022;15 | 24.7 | 27.7 |
| YNHH-0983 | 2022;14 | 24.7 | 28.5 |
| YNHH-1405 | 2022;15 | 24.7 | 28.65 |
| YNHH-1286 | 2022;15 | 24.7 | 28.7 |
| YNHH-1404 | 2022;15 | 24.75 | 28.59 |
| YNHH-1256 | 2022;15 | 24.8 | 24.2 |
| YNHH-1203 | 2022;15 | 24.8 | 24.5 |

| | | | |
|---|---|---|---|
| YNHH-1736 | 2022;15 | 24.8 | 27.8 |
| YNHH-1695 | 2022;15 | 24.8 | 28.4 |
| YNHH-1071 | 2022;14 | 24.8 | 28.6 |
| YNHH-0982 | 2022;14 | 24.8 | 28.7 |
| YNHH-1619 | 2022;15 | 24.9 | 24.5 |
| YNHH-1702 | 2022;15 | 24.9 | 24.6 |
| YNHH-1091 | 2022;14 | 24.9 | 24.7 |
| YNHH-1707 | 2022;15 | 24.9 | 24.7 |
| YNHH-1185 | 2022;15 | 25 | 24.6 |
| YNHH-1618 | 2022;15 | 25 | 24.8 |
| YNHH-1016 | 2022;14 | 25 | 25 |
| YNHH-1247 | 2022;15 | 25 | 25 |
| YNHH-1660 | 2022;15 | 25 | 25 |
| YNHH-1670 | 2022;15 | 25 | 28.7 |
| YNHH-1031 | 2022;14 | 25.1 | 24.5 |
| YNHH-0019 | 2022;10 | 25.1 | 24.8 |
| YNHH-1328 | 2022;15 | 25.1 | 24.8 |
| YNHH-0528 | 2022;13 | 25.1 | 24.9 |
| YNHH-0934 | 2022;14 | 25.1 | 25 |
| YNHH-1666 | 2022;15 | 25.1 | 25.4 |
| YNHH-1680 | 2022;15 | 25.1 | 28.3 |
| YNHH-1345 | 2022;15 | 25.1 | 29 |
| YNHH-1370 | 2022;15 | 25.2 | 24.9 |
| YNHH-1043 | 2022;14 | 25.2 | 25 |
| YNHH-1101 | 2022;14 | 25.2 | 25 |
| YNHH-1510 | 2022;15 | 25.2 | 25 |
| YNHH-1550 | 2022;15 | 25.2 | 25 |
| YNHH-1513 | 2022;15 | 25.2 | 25.1 |
| YNHH-1168 | 2022;15 | 25.2 | 25.3 |
| YNHH-1061 | 2022;14 | 25.2 | 29.1 |
| YNHH-0915 | 2022;14 | 25.3 | 25.1 |
| YNHH-0823 | 2022;14 | 25.3 | 25.2 |
| YNHH-1129 | 2022;14 | 25.4 | 25 |
| YNHH-0842 | 2022;14 | 25.4 | 25.2 |
| YNHH-1252 | 2022;15 | 25.4 | 25.3 |
| YNHH-0932 | 2022;14 | 25.4 | 25.4 |
| YNHH-1613 | 2022;15 | 25.4 | 29 |
| YNHH-0511 | 2022;13 | 25.5 | 25.3 |
| YNHH-1610 | 2022;15 | 25.5 | 25.3 |
| YNHH-1633 | 2022;15 | 25.5 | 25.6 |
| YNHH-0905 | 2022;14 | 25.5 | 29.3 |
| YNHH-0520 | 2022;13 | 25.6 | 25.2 |
| YNHH-0555 | 2022;13 | 25.6 | 25.4 |

| | | | |
|---|---|---|---|
| YNHH-0828 | 2022;14 | 25.6 | 25.4 |
| YNHH-1531 | 2022;15 | 25.6 | 25.4 |
| YNHH-1391 | 2022;15 | 25.6 | 25.5 |
| YNHH-1516 | 2022;15 | 25.7 | 25.4 |
| YNHH-1027 | 2022;14 | 25.7 | 25.6 |
| YNHH-1187 | 2022;15 | 25.7 | 25.6 |
| YNHH-1640 | 2022;15 | 25.8 | 25.3 |
| YNHH-0686 | 2022;13 | 25.8 | 25.4 |
| YNHH-0008 | 2022;14 | 25.8 | 25.5 |
| YNHH-1029 | 2022;14 | 25.8 | 25.5 |
| YNHH-0837 | 2022;14 | 25.8 | 25.6 |
| YNHH-1022 | 2022;14 | 25.8 | 25.6 |
| YNHH-1211 | 2022;15 | 25.8 | 25.6 |
| YNHH-1709 | 2022;15 | 25.8 | 25.6 |
| YNHH-1184 | 2022;15 | 25.9 | 25.5 |
| YNHH-1194 | 2022;15 | 25.9 | 25.6 |
| YNHH-0615 | 2022;13 | 25.9 | 25.7 |
| YNHH-1112 | 2022;14 | 25.9 | 25.7 |
| YNHH-1572 | 2022;15 | 25.9 | 25.7 |
| YNHH-1542 | 2022;15 | 25.9 | 29.5 |
| YNHH-1386 | 2022;15 | 25.92 | 25.26 |
| YNHH-1240 | 2022;15 | 26 | 25.7 |
| YNHH-1549 | 2022;15 | 26 | 25.7 |
| YNHH-1639 | 2022;15 | 26 | 25.7 |
| YNHH-0831 | 2022;14 | 26 | 25.8 |
| YNHH-0959 | 2022;14 | 26 | 25.8 |
| YNHH-1313 | 2022;15 | 26 | 25.8 |
| YNHH-1222 | 2022;15 | 26 | 25.9 |
| YNHH-1113 | 2022;14 | 26 | 29.4 |
| YNHH-1734 | 2022;15 | 26 | 29.8 |
| YNHH-0802 | 2022;14 | 26 | 29.9 |
| YNHH-0024 | 2022;10 | 26.1 | 25.6 |
| YNHH-0336 | 2022;12 | 26.1 | 25.8 |
| YNHH-1346 | 2022;15 | 26.1 | 25.9 |
| YNHH-0860 | 2022;14 | 26.1 | 29.8 |
| YNHH-1651 | 2022;15 | 26.1 | 30.1 |
| YNHH-1122 | 2022;14 | 26.2 | 25.9 |
| YNHH-1302 | 2022;15 | 26.2 | 29.4 |
| YNHH-0827 | 2022;14 | 26.3 | 25.8 |
| YNHH-1532 | 2022;15 | 26.3 | 26.1 |
| YNHH-0992 | 2022;14 | 26.4 | 26.1 |
| YNHH-1045 | 2022;14 | 26.4 | 26.1 |
| YNHH-1226 | 2022;15 | 26.4 | 26.1 |

| | | | |
|---|---|---|---|
| YNHH-1269 | 2022;15 | 26.4 | 26.2 |
| YNHH-0907 | 2022;14 | 26.4 | 26.3 |
| YNHH-1450 | 2022;15 | 26.4 | 26.4 |
| YNHH-1451 | 2022;15 | 26.5 | 26.3 |
| YNHH-1692 | 2022;15 | 26.5 | 30.2 |
| YNHH-0021 | 2022;10 | 26.6 | 26.2 |
| YNHH-0529 | 2022;13 | 26.6 | 26.2 |
| YNHH-0895 | 2022;14 | 26.6 | 26.4 |
| YNHH-1334 | 2022;15 | 26.6 | 30.3 |
| YNHH-1170 | 2022;15 | 26.7 | 26.4 |
| YNHH-1706 | 2022;15 | 26.7 | 26.4 |
| YNHH-1134 | 2022;14 | 26.7 | 26.5 |
| YNHH-0547 | 2022;13 | 26.8 | 26.4 |
| YNHH-1327 | 2022;15 | 26.8 | 26.4 |
| YNHH-0946 | 2022;14 | 26.8 | 26.5 |
| YNHH-1106 | 2022;14 | 26.8 | 26.5 |
| YNHH-0576 | 2022;13 | 26.8 | 26.6 |
| YNHH-1406 | 2022;15 | 26.88 | 30.26 |
| YNHH-1575 | 2022;15 | 26.9 | 26.5 |
| YNHH-1248 | 2022;15 | 26.9 | 26.6 |
| YNHH-1469 | 2022;15 | 26.9 | 30.7 |
| YNHH-0709 | 2022;13 | 27 | 26.7 |
| YNHH-0952 | 2022;14 | 27 | 26.8 |
| YNHH-1486 | 2022;15 | 27 | 26.8 |
| YNHH-1598 | 2022;15 | 27 | 26.9 |
| YNHH-1517 | 2022;15 | 27.1 | 26.8 |
| YNHH-0825 | 2022;14 | 27.1 | 26.9 |
| YNHH-0971 | 2022;14 | 27.1 | 26.9 |
| YNHH-0729 | 2022;13 | 27.1 | 27 |
| YNHH-1661 | 2022;15 | 27.2 | 26.7 |
| YNHH-0928 | 2022;14 | 27.2 | 26.9 |
| YNHH-1499 | 2022;15 | 27.2 | 26.9 |
| YNHH-1485 | 2022;15 | 27.2 | 27 |
| YNHH-1340 | 2022;15 | 27.24 | 26.82 |
| YNHH-0722 | 2022;13 | 27.3 | 27.1 |
| YNHH-1494 | 2022;15 | 27.3 | 27.1 |
| YNHH-1249 | 2022;15 | 27.3 | 31.1 |
| YNHH-0647 | 2022;13 | 27.4 | 27 |
| YNHH-1566 | 2022;15 | 27.4 | 27 |
| YNHH-0848 | 2022;14 | 27.4 | 27.2 |
| YNHH-0368 | 2022;12 | 27.4 | 30.7 |
| YNHH-1466 | 2022;15 | 27.4 | 30.7 |
| YNHH-1037 | 2022;14 | 27.4 | 30.9 |

| | | | |
|---|---|---|---|
| YNHH-0916 | 2022;14 | 27.5 | 26.8 |
| YNHH-1287 | 2022;15 | 27.5 | 27 |
| YNHH-1375 | 2022;15 | 27.5 | 27.2 |
| YNHH-1453 | 2022;15 | 27.5 | 27.3 |
| YNHH-0988 | 2022;14 | 27.5 | 27.5 |
| YNHH-1303 | 2022;15 | 27.5 | 27.9 |
| YNHH-1594 | 2022;15 | 27.6 | 27.1 |
| YNHH-1719 | 2022;15 | 27.6 | 27.1 |
| YNHH-1644 | 2022;15 | 27.6 | 27.3 |
| YNHH-0694 | 2022;13 | 27.7 | 27.1 |
| YNHH-0010 | 2022;15 | 27.7 | 27.3 |
| YNHH-1232 | 2022;15 | 27.7 | 27.4 |
| YNHH-1305 | 2022;15 | 27.8 | 27.3 |
| YNHH-0779 | 2022;14 | 27.8 | 30.9 |
| YNHH-1035 | 2022;14 | 27.9 | 27.5 |
| YNHH-1121 | 2022;14 | 27.9 | 27.7 |
| YNHH-1725 | 2022;15 | 27.9 | 27.7 |
| YNHH-0826 | 2022;14 | 28 | 27.7 |
| YNHH-1260 | 2022;15 | 28 | 27.7 |
| YNHH-0793 | 2022;14 | 28 | 27.8 |
| YNHH-0307 | 2022;12 | 28.1 | 27.7 |
| YNHH-0873 | 2022;14 | 28.1 | 31.3 |
| YNHH-0680 | 2022;13 | 28.2 | 27.8 |
| YNHH-1744 | 2022;15 | 28.2 | 27.9 |
| YNHH-0508 | 2022;13 | 28.3 | 27.6 |
| YNHH-0738 | 2022;13 | 28.3 | 27.9 |
| YNHH-1178 | 2022;15 | 28.3 | 27.9 |
| YNHH-1083 | 2022;14 | 28.3 | 28.1 |
| YNHH-1245 | 2022;15 | 28.3 | 28.4 |
| YNHH-1489 | 2022;15 | 28.3 | 31.9 |
| YNHH-0920 | 2022;14 | 28.4 | 27.8 |
| YNHH-1215 | 2022;15 | 28.4 | 28 |
| YNHH-0796 | 2022;14 | 28.5 | 28 |
| YNHH-1126 | 2022;14 | 28.5 | 28.2 |
| YNHH-1293 | 2022;15 | 28.5 | 28.2 |
| YNHH-1048 | 2022;14 | 28.5 | 28.4 |
| YNHH-1712 | 2022;15 | 28.6 | 28.3 |
| YNHH-0800 | 2022;14 | 28.6 | 28.4 |
| YNHH-1738 | 2022;15 | 28.6 | 28.4 |
| YNHH-1620 | 2022;15 | 28.7 | 27.9 |
| YNHH-0759 | 2022;14 | 28.7 | 28.1 |
| YNHH-0771 | 2022;14 | 28.7 | 28.2 |
| YNHH-1581 | 2022;15 | 28.7 | 28.4 |

| | | | |
|---|---|---|---|
| YNHH-1674 | 2022;15 | 28.7 | 31.9 |
| YNHH-1219 | 2022;15 | 28.7 | 32.3 |
| YNHH-1414 | 2022;15 | 28.77 | 28.38 |
| YNHH-1158 | 2022;14 | 28.8 | 28.6 |
| YNHH-1568 | 2022;15 | 28.8 | 32.2 |
| YNHH-1604 | 2022;15 | 28.9 | 28.6 |
| YNHH-1163 | 2022;15 | 28.9 | 28.8 |
| YNHH-0965 | 2022;14 | 28.9 | 32 |
| YNHH-1488 | 2022;15 | 29 | 28.3 |
| YNHH-0690 | 2022;13 | 29 | 28.5 |
| YNHH-0838 | 2022;14 | 29 | 28.7 |
| YNHH-1514 | 2022;15 | 29.1 | 28.5 |
| YNHH-1072 | 2022;14 | 29.1 | 31.8 |
| YNHH-1155 | 2022;14 | 29.1 | 32.4 |
| YNHH-1552 | 2022;15 | 29.2 | 28.6 |
| YNHH-0704 | 2022;13 | 29.2 | 28.7 |
| YNHH-1300 | 2022;15 | 29.2 | 29.1 |
| YNHH-1446 | 2022;15 | 29.3 | 28.6 |
| YNHH-1044 | 2022;14 | 29.3 | 28.9 |
| YNHH-0979 | 2022;14 | 29.4 | 28.9 |
| YNHH-1505 | 2022;15 | 29.4 | 32 |
| YNHH-0330 | 2022;12 | 29.5 | 28.7 |
| YNHH-0989 | 2022;14 | 29.5 | 29.5 |
| YNHH-0786 | 2022;14 | 29.6 | 29.3 |
| YNHH-0695 | 2022;13 | 29.7 | 29 |
| YNHH-1067 | 2022;14 | 29.7 | 29.4 |
| YNHH-1070 | 2022;14 | 29.7 | 29.4 |
| YNHH-1190 | 2022;15 | 29.7 | 29.5 |
| YNHH-1431 | 2022;15 | 29.7 | 32.7 |
| YNHH-0910 | 2022;14 | 29.8 | 29 |
| YNHH-1188 | 2022;15 | 29.8 | 29.4 |
| YNHH-1231 | 2022;15 | 29.8 | 29.4 |
| YNHH-0539 | 2022;13 | 29.9 | 29 |
| YNHH-1160 | 2022;14 | 29.9 | 29.4 |
| YNHH-1206 | 2022;15 | 29.9 | 29.4 |
| YNHH-1433 | 2022;15 | 29.98 | 33.27 |
| YNHH-1033 | 2022;14 | 30 | 29.6 |
| YNHH-1534 | 2022;15 | 30 | 29.7 |
| YNHH-1673 | 2022;15 | 30 | 33.1 |
| YNHH-1348 | 2022;15 | 30.1 | 29 |
| YNHH-1561 | 2022;15 | 30.1 | 29.9 |
| YNHH-0773 | 2022;14 | 30.2 | 29.6 |
| YNHH-0899 | 2022;14 | 30.2 | 32.8 |

| | | | |
|---|---|---|---|
| YNHH-1417 | 2022;15 | 30.27 | 29.09 |
| YNHH-1658 | 2022;15 | 30.3 | 29.7 |
| YNHH-1606 | 2022;15 | 30.3 | 30 |
| YNHH-0609 | 2022;13 | 30.3 | 32.4 |
| YNHH-1724 | 2022;15 | 30.3 | 32.7 |
| YNHH-0682 | 2022;13 | 30.4 | 29.8 |
| YNHH-0727 | 2022;13 | 30.4 | 29.8 |
| YNHH-1199 | 2022;15 | 30.4 | 29.9 |
| YNHH-1258 | 2022;15 | 30.4 | 29.9 |
| YNHH-1458 | 2022;15 | 30.4 | 30 |
| YNHH-0914 | 2022;14 | 30.4 | 33.1 |
| YNHH-0298 | 2022;12 | 30.5 | 29.5 |
| YNHH-0322 | 2022;12 | 30.5 | 29.6 |
| YNHH-0271 | 2022;12 | 30.5 | 30 |
| YNHH-0944 | 2022;14 | 30.5 | 30 |
| YNHH-0338 | 2022;12 | 30.5 | 30.2 |
| YNHH-0222 | 2022;11 | 30.5 | 32.8 |
| YNHH-0513 | 2022;13 | 30.6 | 29.6 |
| YNHH-0909 | 2022;14 | 30.6 | 29.9 |
| YNHH-1565 | 2022;15 | 30.6 | 29.9 |
| YNHH-1251 | 2022;15 | 30.7 | 30 |
| YNHH-1236 | 2022;15 | 30.7 | 30.4 |
| YNHH-0770 | 2022;14 | 30.8 | 30.3 |
| YNHH-1464 | 2022;15 | 30.8 | 30.6 |
| YNHH-0847 | 2022;14 | 30.9 | 30.4 |
| YNHH-0900 | 2022;14 | 30.9 | 30.4 |
| YNHH-1685 | 2022;15 | 30.9 | 30.6 |
| YNHH-0316 | 2022;12 | 30.9 | 30.7 |
| YNHH-0976 | 2022;14 | 30.9 | 33.4 |
| YNHH-1295 | 2022;15 | 30.9 | 33.7 |
| YNHH-1553 | 2022;15 | 30.9 | 34 |
| YNHH-0126 | 2022;11 | 31 | 30.1 |
| YNHH-0834 | 2022;14 | 31 | 30.3 |
| YNHH-0044 | 2022;10 | 31 | 30.5 |
| YNHH-0663 | 2022;13 | 31 | 30.5 |
| YNHH-0931 | 2022;14 | 31 | 30.6 |
| YNHH-1367 | 2022;15 | 31 | 30.6 |
| YNHH-0366 | 2022;12 | 31.1 | 30.5 |
| YNHH-0419 | 2022;12 | 31.1 | 30.5 |
| YNHH-0355 | 2022;12 | 31.1 | 30.7 |
| YNHH-0958 | 2022;14 | 31.1 | 33.7 |
| YNHH-0206 | 2022;11 | 31.2 | 30.3 |
| YNHH-0162 | 2022;11 | 31.2 | 30.5 |

| | | | |
|---|---|---|---|
| YNHH-0444 | 2022;13 | 31.2 | 30.5 |
| YNHH-0415 | 2022;12 | 31.2 | 30.6 |
| YNHH-0641 | 2022;13 | 31.2 | 30.6 |
| YNHH-0309 | 2022;12 | 31.2 | 30.8 |
| YNHH-1590 | 2022;15 | 31.2 | 31.5 |
| YNHH-1609 | 2022;15 | 31.2 | 33.8 |
| YNHH-0679 | 2022;13 | 31.3 | 30.3 |
| YNHH-0853 | 2022;14 | 31.3 | 30.4 |
| YNHH-0042 | 2022;10 | 31.3 | 30.5 |
| YNHH-0614 | 2022;13 | 31.3 | 30.5 |
| YNHH-0004 | 2022;13 | 31.3 | 30.7 |
| YNHH-0824 | 2022;14 | 31.3 | 31 |
| YNHH-1483 | 2022;15 | 31.3 | 31 |
| YNHH-0054 | 2022;10 | 31.3 | 31.8 |
| YNHH-0864 | 2022;14 | 31.3 | 33 |
| YNHH-1123 | 2022;14 | 31.3 | 33.3 |
| YNHH-0212 | 2022;11 | 31.4 | 30.7 |
| YNHH-0226 | 2022;11 | 31.4 | 30.7 |
| YNHH-0293 | 2022;12 | 31.4 | 30.7 |
| YNHH-0808 | 2022;14 | 31.4 | 30.9 |
| YNHH-1356 | 2022;15 | 31.4 | 30.9 |
| YNHH-0235 | 2022;11 | 31.5 | 30.3 |
| YNHH-0591 | 2022;13 | 31.5 | 30.5 |
| YNHH-0321 | 2022;12 | 31.5 | 30.6 |
| YNHH-0755 | 2022;14 | 31.5 | 31.7 |
| YNHH-1400 | 2022;15 | 31.54 | 33.2 |
| YNHH-0301 | 2022;12 | 31.6 | 30.8 |
| YNHH-0103 | 2022;10 | 31.6 | 30.9 |
| YNHH-1574 | 2022;15 | 31.6 | 31 |
| YNHH-0395 | 2022;12 | 31.6 | 31.1 |
| YNHH-1529 | 2022;15 | 31.6 | 31.1 |
| YNHH-1234 | 2022;15 | 31.6 | 33.9 |
| YNHH-0635 | 2022;13 | 31.7 | 30.9 |
| YNHH-0977 | 2022;14 | 31.7 | 30.9 |
| YNHH-0027 | 2022;10 | 31.7 | 31 |
| YNHH-0050 | 2022;10 | 31.7 | 31 |
| YNHH-1720 | 2022;15 | 31.7 | 31.4 |
| YNHH-0532 | 2022;13 | 31.7 | 31.6 |
| YNHH-1049 | 2022;14 | 31.7 | 34.1 |
| YNHH-1355 | 2022;15 | 31.77 | 30.89 |
| YNHH-1480 | 2022;15 | 31.8 | 30.4 |
| YNHH-0462 | 2022;13 | 31.8 | 30.9 |
| YNHH-0769 | 2022;14 | 31.8 | 31 |

| | | | |
|---|---|---|---|
| YNHH-1461 | 2022;15 | 31.8 | 31.3 |
| YNHH-1082 | 2022;14 | 31.8 | 31.7 |
| YNHH-0441 | 2022;13 | 31.9 | 29.8 |
| YNHH-0041 | 2022;10 | 31.9 | 31.4 |
| YNHH-1585 | 2022;15 | 31.9 | 31.4 |
| YNHH-1042 | 2022;14 | 31.9 | 31.5 |
| YNHH-0139 | 2022;11 | 32 | 30.5 |
| YNHH-0257 | 2022;11 | 32 | 30.9 |
| YNHH-0093 | 2022;10 | 32 | 31.3 |
| YNHH-0830 | 2022;14 | 32 | 31.5 |
| YNHH-0213 | 2022;11 | 32.1 | 31 |
| YNHH-1115 | 2022;14 | 32.1 | 31.2 |
| YNHH-0938 | 2022;14 | 32.1 | 31.6 |
| YNHH-1490 | 2022;15 | 32.2 | 31 |
| YNHH-0990 | 2022;14 | 32.2 | 31.3 |
| YNHH-0968 | 2022;14 | 32.2 | 31.7 |
| YNHH-0184 | 2022;11 | 32.3 | 31.3 |
| YNHH-0469 | 2022;13 | 32.3 | 31.4 |
| YNHH-0030 | 2022;10 | 32.3 | 31.5 |
| YNHH-0038 | 2022;10 | 32.3 | 31.5 |
| YNHH-0599 | 2022;13 | 32.3 | 31.5 |
| YNHH-0064 | 2022;10 | 32.3 | 31.6 |
| YNHH-0637 | 2022;13 | 32.3 | 31.6 |
| YNHH-0993 | 2022;14 | 32.3 | 31.6 |
| YNHH-1392 | 2022;15 | 32.3 | 31.8 |
| YNHH-0091 | 2022;10 | 32.4 | 30.9 |
| YNHH-0255 | 2022;11 | 32.4 | 31 |
| YNHH-0203 | 2022;11 | 32.4 | 31.3 |
| YNHH-0153 | 2022;11 | 32.4 | 31.5 |
| YNHH-0434 | 2022;13 | 32.4 | 31.5 |
| YNHH-0642 | 2022;13 | 32.4 | 31.7 |
| YNHH-0832 | 2022;14 | 32.4 | 31.7 |
| YNHH-0654 | 2022;13 | 32.5 | 31.2 |
| YNHH-1484 | 2022;15 | 32.5 | 31.3 |
| YNHH-1683 | 2022;15 | 32.5 | 31.8 |
| YNHH-0068 | 2022;10 | 32.5 | 31.9 |
| YNHH-1004 | 2022;14 | 32.5 | 32 |
| YNHH-1225 | 2022;15 | 32.5 | 32.2 |
| YNHH-0198 | 2022;11 | 32.6 | 31.8 |
| YNHH-0353 | 2022;12 | 32.6 | 31.8 |
| YNHH-1104 | 2022;14 | 32.6 | 31.8 |
| YNHH-0949 | 2022;14 | 32.6 | 31.9 |
| YNHH-1318 | 2022;15 | 32.6 | 31.9 |

| | | | |
|---|---|---|---|
| YNHH-1452 | 2022;15 | 32.6 | 32.2 |
| YNHH-1243 | 2022;15 | 32.6 | 34.6 |
| YNHH-0345 | 2022;12 | 32.7 | 31.6 |
| YNHH-0401 | 2022;12 | 32.7 | 31.8 |
| YNHH-1634 | 2022;15 | 32.7 | 32 |
| YNHH-1528 | 2022;15 | 32.7 | 32.1 |
| YNHH-0067 | 2022;10 | 32.7 | 32.2 |
| YNHH-0597 | 2022;13 | 32.7 | 32.2 |
| YNHH-1478 | 2022;15 | 32.7 | 32.6 |
| YNHH-1227 | 2022;15 | 32.7 | 34.3 |
| YNHH-0280 | 2022;12 | 32.8 | 31.6 |
| YNHH-0785 | 2022;14 | 32.8 | 31.7 |
| YNHH-0564 | 2022;13 | 32.8 | 31.9 |
| YNHH-0896 | 2022;14 | 32.8 | 32 |
| YNHH-0721 | 2022;13 | 32.8 | 32.1 |
| YNHH-1630 | 2022;15 | 32.8 | 32.5 |
| YNHH-0233 | 2022;11 | 32.9 | 31.1 |
| YNHH-0197 | 2022;11 | 32.9 | 31.2 |
| YNHH-1330 | 2022;15 | 32.9 | 31.4 |
| YNHH-0196 | 2022;11 | 32.9 | 31.6 |
| YNHH-0002 | 2022;12 | 32.9 | 32.1 |
| YNHH-0504 | 2022;13 | 32.9 | 32.2 |
| YNHH-0498 | 2022;13 | 33 | 31.6 |
| YNHH-0214 | 2022;11 | 33 | 32.1 |
| YNHH-0348 | 2022;12 | 33 | 32.1 |
| YNHH-0421 | 2022;12 | 33 | 32.1 |
| YNHH-0602 | 2022;13 | 33 | 32.1 |
| YNHH-1582 | 2022;15 | 33 | 32.9 |
| YNHH-1443 | 2022;15 | 33 | 33.8 |
| YNHH-0413 | 2022;12 | 33.1 | 31 |
| YNHH-0479 | 2022;13 | 33.1 | 31.3 |
| YNHH-0221 | 2022;11 | 33.1 | 32.2 |
| YNHH-0108 | 2022;10 | 33.1 | 32.4 |
| YNHH-0073 | 2022;10 | 33.1 | 32.5 |
| YNHH-0438 | 2022;13 | 33.1 | 32.5 |
| YNHH-1274 | 2022;15 | 33.1 | 35.1 |
| YNHH-1504 | 2022;15 | 33.1 | 35.6 |
| YNHH-0113 | 2022;11 | 33.2 | 32.1 |
| YNHH-1551 | 2022;15 | 33.2 | 32.2 |
| YNHH-0572 | 2022;13 | 33.2 | 32.3 |
| YNHH-1369 | 2022;15 | 33.2 | 32.3 |
| YNHH-0406 | 2022;12 | 33.2 | 32.6 |
| YNHH-0287 | 2022;12 | 33.2 | 32.9 |

| | | | |
|---|---|---|---|
| YNHH-1432 | 2022;15 | 33.23 | 32.64 |
| YNHH-0136 | 2022;11 | 33.3 | 31.8 |
| YNHH-1427 | 2022;15 | 33.3 | 32.4 |
| YNHH-1182 | 2022;15 | 33.3 | 32.5 |
| YNHH-0065 | 2022;10 | 33.3 | 33.3 |
| YNHH-0933 | 2022;14 | 33.3 | 36.5 |
| YNHH-0595 | 2022;13 | 33.4 | 31.2 |
| YNHH-0703 | 2022;13 | 33.4 | 32 |
| YNHH-1456 | 2022;15 | 33.4 | 32.7 |
| YNHH-0610 | 2022;13 | 33.4 | 33 |
| YNHH-0349 | 2022;12 | 33.5 | 32.4 |
| YNHH-0012 | 2022;11 | 33.5 | 32.5 |
| YNHH-0732 | 2022;13 | 33.5 | 32.5 |
| YNHH-0209 | 2022;11 | 33.5 | 32.7 |
| YNHH-0392 | 2022;12 | 33.5 | 32.8 |
| YNHH-1631 | 2022;15 | 33.5 | 32.8 |
| YNHH-1228 | 2022;15 | 33.5 | 32.9 |
| YNHH-1445 | 2022;15 | 33.5 | 35.2 |
| YNHH-0858 | 2022;14 | 33.6 | 31.9 |
| YNHH-0581 | 2022;13 | 33.6 | 32.9 |
| YNHH-1645 | 2022;15 | 33.6 | 32.9 |
| YNHH-0521 | 2022;13 | 33.6 | 33 |
| YNHH-0643 | 2022;13 | 33.6 | 33 |
| YNHH-1096 | 2022;14 | 33.6 | 33.1 |
| YNHH-1125 | 2022;14 | 33.6 | 33.1 |
| YNHH-0020 | 2022;10 | 33.7 | 31.2 |
| YNHH-0449 | 2022;13 | 33.7 | 32.5 |
| YNHH-0138 | 2022;11 | 33.7 | 32.6 |
| YNHH-0887 | 2022;14 | 33.7 | 32.8 |
| YNHH-0433 | 2022;13 | 33.7 | 33 |
| YNHH-1266 | 2022;15 | 33.7 | 33.2 |
| YNHH-0660 | 2022;13 | 33.7 | 35.7 |
| YNHH-0058 | 2022;10 | 33.8 | 32.9 |
| YNHH-0350 | 2022;12 | 33.8 | 32.9 |
| YNHH-1023 | 2022;14 | 33.8 | 32.9 |
| YNHH-1262 | 2022;15 | 33.8 | 32.9 |
| YNHH-1491 | 2022;15 | 33.8 | 33.4 |
| YNHH-1164 | 2022;15 | 33.8 | 35.7 |
| YNHH-0927 | 2022;14 | 33.9 | 32.7 |
| YNHH-0178 | 2022;11 | 33.9 | 33.1 |
| YNHH-0268 | 2022;12 | 34 | 32.1 |
| YNHH-0313 | 2022;12 | 34 | 32.5 |
| YNHH-0570 | 2022;13 | 34 | 32.8 |

| | | | |
|---|---|---|---|
| YNHH-0784 | 2022;14 | 34 | 32.8 |
| YNHH-0874 | 2022;14 | 34 | 32.8 |
| YNHH-0889 | 2022;14 | 34 | 33 |
| YNHH-0063 | 2022;10 | 34 | 33.3 |
| YNHH-0177 | 2022;11 | 34 | 33.4 |
| YNHH-1192 | 2022;15 | 34 | 33.6 |
| YNHH-1690 | 2022;15 | 34.1 | 33 |
| YNHH-0554 | 2022;13 | 34.1 | 33.2 |
| YNHH-0813 | 2022;14 | 34.1 | 33.3 |
| YNHH-0862 | 2022;14 | 34.1 | 33.7 |
| YNHH-0405 | 2022;12 | 34.2 | 32.8 |
| YNHH-0267 | 2022;12 | 34.2 | 33 |
| YNHH-0753 | 2022;14 | 34.2 | 33.4 |
| YNHH-1460 | 2022;15 | 34.2 | 33.7 |
| YNHH-0748 | 2022;13 | 34.3 | 32.6 |
| YNHH-1281 | 2022;15 | 34.3 | 32.8 |
| YNHH-0538 | 2022;13 | 34.3 | 33.3 |
| YNHH-1559 | 2022;15 | 34.3 | 33.6 |
| YNHH-1681 | 2022;15 | 34.3 | 33.7 |
| YNHH-1614 | 2022;15 | 34.3 | 34 |
| YNHH-0151 | 2022;11 | 34.4 | 32.9 |
| YNHH-0621 | 2022;13 | 34.4 | 33.1 |
| YNHH-0341 | 2022;12 | 34.4 | 33.2 |
| YNHH-0535 | 2022;13 | 34.4 | 33.4 |
| YNHH-0741 | 2022;13 | 34.4 | 33.5 |
| YNHH-0005 | 2022;13 | 34.4 | 33.6 |
| YNHH-0560 | 2022;13 | 34.4 | 33.6 |
| YNHH-1407 | 2022;15 | 34.49 | 34.3 |
| YNHH-0733 | 2022;13 | 34.5 | 33.1 |
| YNHH-0765 | 2022;14 | 34.5 | 33.1 |
| YNHH-0053 | 2022;10 | 34.5 | 33.6 |
| YNHH-1324 | 2022;15 | 34.5 | 33.8 |
| YNHH-0652 | 2022;13 | 34.5 | 34 |
| YNHH-1162 | 2022;15 | 34.5 | 35.8 |
| YNHH-0211 | 2022;11 | 34.6 | 33.2 |
| YNHH-0431 | 2022;12 | 34.6 | 33.6 |
| YNHH-0633 | 2022;13 | 34.6 | 33.6 |
| YNHH-1667 | 2022;15 | 34.6 | 33.6 |
| YNHH-0356 | 2022;12 | 34.6 | 34.1 |
| YNHH-1495 | 2022;15 | 34.7 | 33.2 |
| YNHH-1538 | 2022;15 | 34.7 | 33.2 |
| YNHH-1602 | 2022;15 | 34.7 | 33.4 |
| YNHH-0507 | 2022;13 | 34.7 | 33.6 |

| | | | |
|---|---|---|---|
| YNHH-0124 | 2022;11 | 34.7 | 33.7 |
| YNHH-1241 | 2022;15 | 34.7 | 33.7 |
| YNHH-1102 | 2022;14 | 34.7 | 33.9 |
| YNHH-1703 | 2022;15 | 34.7 | 34 |
| YNHH-1743 | 2022;15 | 34.7 | 34 |
| YNHH-0359 | 2022;12 | 34.7 | 34.1 |
| YNHH-0179 | 2022;11 | 34.8 | 33.2 |
| YNHH-0141 | 2022;11 | 34.8 | 33.4 |
| YNHH-1448 | 2022;15 | 34.8 | 33.4 |
| YNHH-0134 | 2022;11 | 34.8 | 33.5 |
| YNHH-0199 | 2022;11 | 34.8 | 33.5 |
| YNHH-0930 | 2022;14 | 34.8 | 33.5 |
| YNHH-0032 | 2022;10 | 34.8 | 33.8 |
| YNHH-0787 | 2022;14 | 34.8 | 34.1 |
| YNHH-1336 | 2022;15 | 34.9 | 33.5 |
| YNHH-0661 | 2022;13 | 34.9 | 33.8 |
| YNHH-1520 | 2022;15 | 34.9 | 33.8 |
| YNHH-0047 | 2022;10 | 34.9 | 33.9 |
| YNHH-0708 | 2022;13 | 34.9 | 33.9 |
| YNHH-1239 | 2022;15 | 34.9 | 33.9 |
| YNHH-0048 | 2022;10 | 34.9 | 34.1 |
| YNHH-0092 | 2022;10 | 34.9 | 34.4 |
| YNHH-1410 | 2022;15 | 34.96 | 33.84 |
| YNHH-0478 | 2022;13 | 35 | 33.4 |
| YNHH-0039 | 2022;10 | 35 | 33.6 |
| YNHH-0318 | 2022;12 | 35 | 33.8 |
| YNHH-0617 | 2022;13 | 35 | 33.9 |
| YNHH-0939 | 2022;14 | 35 | 34 |
| YNHH-0743 | 2022;13 | 35 | 34.1 |
| YNHH-1580 | 2022;15 | 35 | 34.5 |
| YNHH-1333 | 2022;15 | 35 | 35 |
| YNHH-1439 | 2022;15 | 35.02 | 34.02 |
| YNHH-1449 | 2022;15 | 35.1 | 33.4 |
| YNHH-0885 | 2022;14 | 35.1 | 33.8 |
| YNHH-0075 | 2022;10 | 35.1 | 33.9 |
| YNHH-0935 | 2022;14 | 35.1 | 34.1 |
| YNHH-1508 | 2022;15 | 35.1 | 34.5 |
| YNHH-1323 | 2022;15 | 35.1 | 36.7 |
| YNHH-0530 | 2022;13 | 35.2 | 33.8 |
| YNHH-1408 | 2022;15 | 35.2 | 33.8 |
| YNHH-0017 | 2022;10 | 35.2 | 33.9 |
| YNHH-0074 | 2022;10 | 35.2 | 34 |
| YNHH-0533 | 2022;13 | 35.2 | 34 |

| | | | |
|---|---|---|---|
| YNHH-0906 | 2022;14 | 35.2 | 34.4 |
| YNHH-0942 | 2022;14 | 35.2 | 34.5 |
| YNHH-0945 | 2022;14 | 35.2 | 34.5 |
| YNHH-1284 | 2022;15 | 35.2 | 35.5 |
| YNHH-0299 | 2022;12 | 35.3 | 33.6 |
| YNHH-0080 | 2022;10 | 35.3 | 33.9 |
| YNHH-0676 | 2022;13 | 35.3 | 34 |
| YNHH-1562 | 2022;15 | 35.3 | 34.4 |
| YNHH-1662 | 2022;15 | 35.3 | 34.5 |
| YNHH-1502 | 2022;15 | 35.3 | 34.7 |
| YNHH-0861 | 2022;14 | 35.4 | 33.7 |
| YNHH-0499 | 2022;13 | 35.4 | 34 |
| YNHH-0455 | 2022;13 | 35.4 | 34.3 |
| YNHH-0851 | 2022;14 | 35.4 | 34.5 |
| YNHH-0062 | 2022;10 | 35.4 | 34.7 |
| YNHH-1442 | 2022;15 | 35.4 | 34.8 |
| YNHH-0603 | 2022;13 | 35.5 | 33.9 |
| YNHH-0750 | 2022;14 | 35.5 | 34.4 |
| YNHH-0953 | 2022;14 | 35.5 | 34.5 |
| YNHH-1593 | 2022;15 | 35.5 | 34.8 |
| YNHH-1727 | 2022;15 | 35.5 | 34.9 |
| YNHH-1669 | 2022;15 | 35.5 | 35 |
| YNHH-1625 | 2022;15 | 35.5 | 35.1 |
| YNHH-1136 | 2022;14 | 35.5 | 35.4 |
| YNHH-0354 | 2022;12 | 35.6 | 33.3 |
| YNHH-0814 | 2022;14 | 35.6 | 34.1 |
| YNHH-1584 | 2022;15 | 35.6 | 34.4 |
| YNHH-0756 | 2022;14 | 35.6 | 34.5 |
| YNHH-0292 | 2022;12 | 35.6 | 34.7 |
| YNHH-0882 | 2022;14 | 35.6 | 34.7 |
| YNHH-1183 | 2022;15 | 35.6 | 34.9 |
| YNHH-1154 | 2022;14 | 35.6 | 35.6 |
| YNHH-0144 | 2022;11 | 35.7 | 33.2 |
| YNHH-0723 | 2022;13 | 35.7 | 34.2 |
| YNHH-0026 | 2022;10 | 35.7 | 34.5 |
| YNHH-1337 | 2022;15 | 35.7 | 34.6 |
| YNHH-0036 | 2022;10 | 35.7 | 34.7 |
| YNHH-0669 | 2022;13 | 35.8 | 34.3 |
| YNHH-1382 | 2022;15 | 35.8 | 34.4 |
| YNHH-1524 | 2022;15 | 35.8 | 34.4 |
| YNHH-1010 | 2022;14 | 35.8 | 34.7 |
| YNHH-1589 | 2022;15 | 35.8 | 34.7 |
| YNHH-1512 | 2022;15 | 35.8 | 34.9 |

| | | | |
|---|---|---|---|
| YNHH-1166 | 2022;15 | 35.9 | 34.8 |
| YNHH-1195 | 2022;15 | 35.9 | 35.1 |
| YNHH-0022 | 2022;10 | 35.9 | 35.3 |
| YNHH-0396 | 2022;12 | 36 | 33.7 |
| YNHH-0167 | 2022;11 | 36 | 34.4 |
| YNHH-0164 | 2022;11 | 36 | 34.6 |
| YNHH-1041 | 2022;14 | 36 | 34.9 |
| YNHH-1073 | 2022;14 | 36 | 35 |
| YNHH-1140 | 2022;14 | 36 | 35 |
| YNHH-0501 | 2022;13 | 36 | 35.1 |
| YNHH-0372 | 2022;12 | 36.1 | 34.4 |
| YNHH-1316 | 2022;15 | 36.1 | 34.4 |
| YNHH-1739 | 2022;15 | 36.1 | 34.6 |
| YNHH-0201 | 2022;11 | 36.1 | 34.7 |
| YNHH-1541 | 2022;15 | 36.1 | 35.3 |
| YNHH-0568 | 2022;13 | 36.2 | 34.6 |
| YNHH-0081 | 2022;10 | 36.2 | 34.7 |
| YNHH-0645 | 2022;13 | 36.2 | 34.9 |
| YNHH-0191 | 2022;11 | 36.2 | 35.1 |
| YNHH-1090 | 2022;14 | 36.2 | 35.5 |
| YNHH-0107 | 2022;10 | 36.2 | 35.7 |
| YNHH-0249 | 2022;11 | 36.3 | 34.3 |
| YNHH-0015 | 2022;10 | 36.3 | 35.1 |
| YNHH-1465 | 2022;15 | 36.3 | 35.1 |
| YNHH-0775 | 2022;14 | 36.3 | 35.3 |
| YNHH-1020 | 2022;14 | 36.3 | 35.8 |
| YNHH-1103 | 2022;14 | 36.4 | 35.2 |
| YNHH-1726 | 2022;15 | 36.4 | 35.3 |
| YNHH-1595 | 2022;15 | 36.4 | 35.4 |
| YNHH-1430 | 2022;15 | 36.42 | 34.93 |
| YNHH-0142 | 2022;11 | 36.5 | 34.8 |
| YNHH-0619 | 2022;13 | 36.5 | 35.7 |
| YNHH-0859 | 2022;14 | 36.5 | 35.8 |
| YNHH-0051 | 2022;10 | 36.5 | 36.2 |
| YNHH-1169 | 2022;15 | 36.5 | 37 |
| YNHH-1110 | 2022;14 | 36.6 | 34.7 |
| YNHH-0227 | 2022;11 | 36.6 | 35.6 |
| YNHH-1686 | 2022;15 | 36.6 | 35.9 |
| YNHH-0486 | 2022;13 | 36.6 | 36.5 |
| YNHH-0473 | 2022;13 | 36.7 | 35.1 |
| YNHH-1315 | 2022;15 | 36.7 | 35.3 |
| YNHH-1597 | 2022;15 | 36.7 | 35.4 |
| YNHH-0667 | 2022;13 | 36.7 | 36 |

| | | | |
|---|---|---|---|
| YNHH-1153 | 2022;14 | 36.8 | 34.9 |
| YNHH-0394 | 2022;12 | 36.8 | 35.2 |
| YNHH-1322 | 2022;15 | 36.8 | 36.6 |
| YNHH-0616 | 2022;13 | 36.9 | 35.3 |
| YNHH-0172 | 2022;11 | 37 | 35.4 |
| YNHH-1381 | 2022;15 | 37 | 36.6 |
| YNHH-1308 | 2022;15 | 37.2 | 36.2 |
| YNHH-0006 | 2022;14 | 37.2 | 36.5 |
| YNHH-0412 | 2022;12 | 37.3 | 35.4 |
| YNHH-0698 | 2022;13 | 37.3 | 36.1 |
| YNHH-0118 | 2022;11 | 37.4 | 34.9 |
| YNHH-0071 | 2022;10 | 37.4 | 35.1 |
| YNHH-1521 | 2022;15 | 37.4 | 36.2 |
| YNHH-0332 | 2022;12 | 37.5 | 35.5 |
| YNHH-1587 | 2022;15 | 37.5 | 36.6 |
| YNHH-0009 | 2022;15 | 37.59 | 35.1 |
| YNHH-0094 | 2022;10 | 37.6 | 35.7 |
| YNHH-1411 | 2022;15 | 37.72 | 35.72 |
| YNHH-0045 | 2022;10 | 37.8 | 34.9 |
| YNHH-1424 | 2022;15 | 37.84 | 35.64 |
| YNHH-0523 | 2022;13 | 37.9 | 34.6 |
| YNHH-0658 | 2022;13 | 37.9 | 34.8 |
| YNHH-0057 | 2022;10 | 37.9 | 34.9 |
| YNHH-1384 | 2022;15 | 37.9 | 37.5 |
| YNHH-0497 | 2022;13 | 38 | 35.1 |
| YNHH-1612 | 2022;15 | 38 | 36.5 |
| YNHH-0023 | 2022;10 | 38.1 | 35.5 |
| YNHH-0841 | 2022;14 | 38.1 | 36.5 |
| YNHH-0007 | 2022;14 | 38.1 | 36.8 |
| YNHH-1030 | 2022;14 | 38.1 | 36.8 |
| YNHH-0037 | 2022;10 | 38.4 | 35.1 |
| YNHH-0120 | 2022;11 | 38.4 | 35.4 |

| Lineage |
| --- |
| BA.1.15 |
| BA.1.1 |
| BA.1.1 |
| BA.1.1 |
| BA.1.1 |
| BA.1.1 |
| BA.1.1 |
| BA.1.1 |
| BA.2 |
| BA.2.9 |
| BA.2.10 |
| B.1.596 |
| BA.2 |
| BA.2 |
| BA.2 |
| BA.2 |
| BA.2 |
| BA.2.9 |
| BA.2.9 |
| BA.2 |
| BA.2 |
| BA.2 |
| BA.2 |
| BA.2.3 |
| BA.2.3 |
| BA.2.3 |
| BA.1.1 |
| BA.1.1 |
| BA.1.1 |
| BA.1.1 |
| BA.1.1 |
| BA.1.1 |
| BA.1.1 |
| BA.1.1 |
| BA.1.1 |
| BA.1.1 |
| BA.1.1 |
| BA.1.1 |
| BA.1.1 |
| BA.1.1 |
| BA.1.1 |
| BA.1.1 |

| |
|---|
| BA.1.1 |
| BA.1.1 |
| BA.1.1 |
| BA.1.1 |
| BA.1.1 |
| BA.1.1 |
| BA.1.1 |
| BA.1.1 |
| BA.1.1 |
| BA.1.1 |
| BA.1.1 |
| BA.1.1 |
| BA.1.1 |
| BA.1.1 |
| BA.1.1 |
| BA.1.1 |
| BA.2.3 |
| BA.2 |
| BA.2.9 |
| BA.2.12 |
| BA.2.9 |
| BA.2 |
| BA.2 |
| BA.2.3 |
| BA.2 |
| BA.2 |
| BA.2 |
| BA.2 |
| BA.2.9 |
| BA.2 |
| BA.2 |
| BA.2.3 |
| BA.2.3 |
| BA.2 |
| BA.2.9 |
| BA.2 |
| BA.2.9 |
| BA.2 |
| BA.2 |
| BA.1.1 |
| BA.1.1 |
| BA.1.1 |
| BA.1.1 |

| |
|---|
| BA.1.1 |
| BA.1.1 |
| BA.1.1 |
| BA.1.1 |
| BA.1.1 |
| BA.1.1 |
| BA.1.1 |
| BA.1.1 |
| BA.1.1 |
| BA.1.1 |
| BA.1.1 |
| BA.1.1 |
| BA.1.1 |
| BA.1.1 |
| BA.1 |
| AY.103 |
| BA.2 |
| BA.1.1 |
| BA.2 |
| BA.1.1 |
| BA.2 |
| BA.1 |
| BA.2 |
| BA.1.1 |
| BA.2 |
| BA.2 |
| BA.2 |
| BA.2 |
| BA.2 |
| BA.2 |
| BA.1.1 |
| BA.2 |
| BA.1.1 |
| BA.1.1 |
| BA.2 |
| BA.2 |
| BA.2 |
| BA.2 |
| BA.1.1 |
| BA.2 |
| BA.2 |
| BA.1.1 |
| BA.2 |

| |
|---|
| BA.2 |
| BA.1.1 |
| BA.1 |
| BA.1 |
| BA.2 |
| BA.1.1 |
| BA.2 |
| BA.1.1 |
| BA.1.1 |
| BA.2 |
| BA.1.1 |
| BA.2 |
| BA.1.1 |
| BA.1.1 |
| BA.1.1 |
| BA.2 |
| BA.2 |
| BA.2 |
| BA.2 |
| BA.2 |
| BA.1.1 |
| BA.1.1 |
| BA.1.1 |
| BA.2 |
| BA.1.1 |
| BA.2 |
| BA.1.1 |
| BA.2 |
| BA.1 |
| BA.2 |
| BA.2 |
| BA.2 |
| BA.2 |
| BA.1.1 |
| BA.1.1 |
| BA.2 |
| BA.2 |
| BA.1.1 |
| BA.1.1 |
| BA.2 |
| BA.1.1 |
| BA.2 |
| BA.1.1 |

| |
|---|
| BA.2 |
| BA.2 |
| BA.2 |
| BA.2 |
| BA.2 |
| BA.2 |
| BA.2 |
| BA.1.1 |
| BA.2 |
| BA.2 |
| BA.1.1 |
| BA.1.1 |
| BA.1.1 |
| BA.1.1 |
| BA.2 |
| BA.2 |
| BA.2.9 |
| BA.2 |
| BA.2.9 |
| BA.2 |
| BA.2.10 |
| BA.2 |
| BA.2 |
| BA.2.3 |
| BA.2 |
| BA.2 |
| BA.2 |
| BA.2.3 |
| BA.2 |
| BA.2 |
| BA.2 |
| BA.2 |
| BA.2.9 |
| BA.2 |
| BA.2.3 |
| BA.2.12.1 |
| BA.2 |
| BA.2.9 |
| BA.2 |
| BA.2 |
| BA.2 |
| BA.2 |
| BA.2 |

| |
|---|
| BA.2.12 |
| BA.2 |
| BA.2 |
| BA.2 |
| BA.2 |
| BA.2 |
| BA.2.9 |
| BA.2.12 |
| BA.1.1 |
| BA.1.1 |
| BA.1.1 |
| BA.1.1 |
| BA.1.1 |
| BA.1.1 |
| BA.1.1 |
| BA.1.1 |
| BA.1.1 |
| BA.1.1 |
| BA.1.1 |
| BA.1.1 |
| BA.1.1 |
| BA.1.15 |
| BA.1.1 |
| BA.1.15 |
| BA.1.15 |
| BA.1 |
| BA.1 |
| BA.2 |
| BA.2 |
| BA.2 |
| BA.2 |
| BA.2 |
| BA.2 |
| BA.2 |
| BA.2 |
| BA.2 |
| BA.2 |
| BA.2 |
| BA.2 |
| BA.2 |
| BA.2 |
| BA.2 |
| BA.2 |

| |
|---|
| BA.2 |
| BA.2 |
| BA.2 |
| BA.2 |
| BA.2 |
| BA.2 |
| BA.2 |
| BA.2 |
| BA.2 |
| BA.2 |
| BA.2 |
| BA.2 |
| BA.2 |
| BA.2 |
| BA.2 |
| BA.2 |
| BA.2 |
| BA.2 |
| BA.2 |
| BA.2 |
| BA.2 |
| BA.2 |
| BA.2 |
| BA.2 |
| BA.2 |
| BA.2 |
| BA.2 |
| BA.1.1 |
| BA.1.1 |
| BA.1 |
| BA.1.1 |
| BA.1.1 |
| BA.1 |
| BA.1 |
| BA.1.1 |
| BA.1.1 |
| BA.1.1 |
| BA.1.1 |
| BA.1.1 |
| BA.1.1 |
| BA.1.1 |
| BA.1.1 |
| BA.1.1 |

| |
|---|
| BA.1.1 |
| BA.1 |
| BA.1.1 |
| BA.1.1 |
| BA.1.1 |
| BA.1 |
| BA.2 |
| BA.2 |
| BA.2 |
| BA.2 |
| BA.1.1 |
| BA.2 |
| BA.2 |
| BA.2 |
| BA.2 |
| BA.2 |
| BA.2 |
| BA.2 |
| BA.2 |
| BA.2 |
| BA.2 |
| BA.1.1 |
| BA.1.1 |
| BA.1.1 |
| BA.2 |
| BA.2 |
| BA.1.1 |
| BA.1.1 |
| BA.2 |
| BA.2 |
| BA.2 |
| BA.1.1 |
| BA.2 |
| BA.1.1 |
| BA.1.1 |
| BA.2 |
| BA.2.12 |
| BA.2 |
| BA.2.12.1 |
| BA.2 |
| BA.2.12.1 |
| BA.2 |
| BA.2.9 |

| |
|---|
| BA.2 |
| BA.2 |
| BA.2 |
| BA.2 |
| BA.2 |
| BA.2.12.1 |
| BA.2.12.1 |
| BA.2.3 |
| BA.2 |
| BA.2 |
| BA.2.12.1 |
| BA.2.12.1 |
| BA.2 |
| BA.2 |
| BA.2 |
| BA.2.3 |
| BA.2 |
| BA.2 |
| BA.2.9 |
| BA.2.9 |
| BA.2.12.1 |
| BA.2.3 |
| BA.2 |
| BA.2.12.1 |
| BA.2.3 |
| BA.2 |
| BA.2.9 |
| BA.2.3 |
| BA.2 |
| BA.2 |
| BA.2.12.1 |
| BA.2.9 |
| BA.2 |
| BA.2.12.1 |
| BA.2 |
| BA.2 |
| BA.2 |
| BA.2.9 |
| BA.2.9 |
| BA.2.12 |
| BA.2 |
| BA.2.9 |
| BA.2.3 |

| |
|---|
| BA.2 |
| BA.2.12 |
| BA.2 |
| BA.2.3 |
| BA.2 |
| BA.2.12.1 |
| BA.2.3 |
| BA.2.3 |
| BA.2.3 |
| BA.2.3 |
| BA.2.3 |
| BA.2.3 |
| BA.2 |
| BA.2 |
| BA.2 |
| BA.1.1 |
| BA.1.1 |
| BA.1.1 |
| BA.1.1 |
| BA.1.1 |
| BA.1.1 |
| BA.1.1 |
| BA.1.15 |
| BA.1.1 |
| BA.1.1 |
| BA.1.1 |
| BA.1.1 |
| BA.2.10 |
| BA.1.1 |
| BA.1.1 |
| BA.1.1 |
| AY.3 |
| BA.2 |
| BA.1.15 |
| BA.1.1 |
| BA.2 |
| BA.2 |
| BA.2 |
| BA.1.1 |
| BA.1.1 |
| BA.2.9 |
| BA.2.10 |
| BA.1.1 |

| |
|---|
| BA.2.9 |
| BA.1.1 |
| BA.2 |
| BA.2.3 |
| BA.1.1 |
| BA.2 |
| BA.1.1 |
| BA.2 |
| BA.2 |
| BA.2 |
| BA.1.1 |
| BA.2.12.1 |
| BA.2.12.1 |
| BA.2.3 |
| BA.2.12.1 |
| BA.2.9 |
| BA.2 |
| BA.2.12.1 |
| BA.1.1 |
| BA.2 |
| BA.1.1 |
| BA.2 |
| BA.2 |
| BA.2 |
| BA.2 |
| BA.2 |
| BA.2 |
| BA.2 |
| BA.2 |
| BA.1.1 |
| BA.2.9 |
| BA.2 |
| BA.1.1 |
| BA.2.9 |
| BA.1.1 |
| BA.2 |
| BA.2 |
| BA.1.1 |
| BA.1.1 |
| BA.2.9 |
| BA.2 |
| BA.2.3 |
| BA.2 |

| |
|---|
| BA.1.1 |
| BA.2.3 |
| BA.1.1 |
| BA.1.1 |
| BA.2 |
| BA.2 |
| BA.2 |
| BA.1.1 |
| BA.2.7 |
| BA.2 |
| BA.1.1 |
| BA.2 |
| BA.2 |
| BA.2 |
| BA.2.7 |
| BA.2.9 |
| BA.1.1 |
| BA.2 |
| BA.1.1 |
| BA.2 |
| BA.1.1 |
| BA.1.1 |
| BA.1.1 |
| BA.2 |
| BA.2 |
| BA.1.1 |
| BA.2.9 |
| BA.2 |
| BA.1.1 |
| BA.2 |
| BA.1.1 |
| BA.1.1 |
| BA.1.1 |
| BA.2 |
| BA.2.12.1 |
| BA.2 |
| BA.2.7 |
| BA.1.1 |
| BA.2.7 |
| BA.2 |
| BA.2 |
| BA.2.7 |
| BA.2.7 |

| |
|---|
| BA.2.7 |
| BA.2 |
| BA.2 |
| BA.2 |
| BA.2.7 |
| BA.2.7 |
| BA.2.7 |
| BA.2 |
| BA.2 |
| BA.2 |
| BA.2 |
| BA.2 |
| BA.1.1 |
| BA.2.7 |
| BA.2 |
| BA.2 |
| BA.2 |
| BA.2 |
| BA.2 |
| BA.2.9 |
| BA.2.9 |
| BA.2 |
| BA.2.12.1 |
| BA.1.1 |
| BA.1.1 |
| BA.2 |
| BA.1.1 |
| BA.1.1 |
| BA.2.7 |
| BA.1.1 |
| BA.2 |
| BA.2 |
| BA.2 |
| BA.1.1 |
| BA.1.1 |
| BA.2 |
| BA.2.3 |
| BA.2 |
| BA.2 |
| BA.2 |
| BA.2.9 |
| BA.1.1 |
| BA.2.9 |

| |
|---|
| BA.2 |
| BA.2 |
| BA.2 |
| BA.2.7 |
| BA.2.7 |
| BA.1.1 |
| BA.2 |
| BA.2 |
| BA.2 |
| BA.2 |
| BA.2 |
| BA.2.10.1 |
| BA.2.12.1 |
| BA.2.9 |
| BA.2 |
| BA.2 |
| BA.2 |
| BA.2 |
| BA.2.3 |
| BA.2.3 |
| BA.2 |
| BA.2 |
| BA.2.3 |
| BA.2 |
| BA.2.3 |
| BA.2 |
| BA.2.12.1 |
| BA.2.12.1 |
| BA.2.9 |
| BA.2.9 |
| BA.2.3 |
| BA.2 |
| BA.2.9 |
| BA.2 |
| BA.2 |
| BA.2 |
| BA.2.3 |
| BA.2 |
| BA.2 |
| BA.2.10 |
| BA.2 |
| BA.2.3 |
| BA.2.12.1 |

| |
|---|
| BA.2 |
| BA.2 |
| BA.2.9 |
| BA.2.12.1 |
| BA.2 |
| BA.2.12.1 |
| BA.2 |
| BA.2.9 |
| BA.2.12.1 |
| BA.2.9 |
| BA.2.3 |
| BA.2 |
| BA.2.3 |
| BA.2 |
| BA.2.12.1 |
| BA.2.12.1 |
| BA.2.10 |
| BA.2 |
| BA.2.12.1 |
| BA.2 |
| BA.2 |
| BA.2 |
| BA.2.9 |
| BA.2 |
| BA.2 |
| BA.2.12.1 |
| BA.2.12.1 |
| BA.2 |
| BA.2.3 |
| BA.2.3 |
| BA.2.12.1 |
| BA.2 |
| BA.2 |
| BA.2.7 |
| BA.2 |
| BA.2.9 |
| BA.2.7 |
| BA.2 |
| BA.2 |
| BA.1.1 |
| BA.2 |
| BA.2.12.1 |
| BA.2 |

| |
|---|
| BA.2.3 |
| BA.1.1 |
| BA.1.1 |
| BA.2 |
| BA.2.9 |
| BA.2.12.1 |
| BA.2 |
| BA.2 |
| BA.2.7 |
| BA.2.9 |
| BA.2 |
| BA.2 |
| BA.2 |
| BA.2.12.1 |
| BA.2.12.1 |
| BA.2 |
| BA.2 |
| BA.2 |
| BA.2 |
| BA.2 |
| BA.2 |
| BA.2.10 |
| BA.2 |
| BA.2 |
| BA.2 |
| BA.2 |
| BA.2.9 |
| BA.2 |
| BA.2.7 |
| BA.2 |
| BA.2 |
| BA.2 |
| BA.2 |
| BA.2 |
| BA.1.1 |
| BA.2.7 |
| BA.2.12.1 |
| BA.2.9 |
| BA.2 |
| BA.2 |
| BA.2.9 |
| BA.2 |
| BA.2.7 |

| |
|---|
| BA.2.12.1 |
| BA.2.7 |
| BA.2 |
| BA.2 |
| BA.2 |
| BA.1.1 |
| BA.2 |
| BA.2 |
| BA.2 |
| BA.1.1 |
| BA.2.12.1 |
| BA.2 |
| BA.2.12.1 |
| BA.2 |
| BA.2.9 |
| BA.3 |
| BA.1 |
| BA.2.3 |
| BA.2.7 |
| BA.2 |
| BA.2 |
| BA.2 |
| BA.2 |
| BA.2 |
| BA.2 |
| BA.1.1.14 |
| BA.2 |
| BA.2 |
| BA.2.10 |
| BA.2 |
| BA.2 |
| BA.2 |
| BA.2.9 |
| BA.2 |
| BA.2.7 |
| BA.2.10 |
| BA.2.7 |
| BA.2.7 |
| BA.1.1 |
| BA.2 |
| BA.2 |
| BA.2 |
| BA.2 |

| |
|---|
| BA.2.7 |
| BA.2 |
| BA.2 |
| BA.1.1 |
| BA.2.9 |
| BA.2 |
| BA.2 |
| BA.2 |
| BA.2 |
| BA.2 |
| BA.2 |
| BA.2.9 |
| BA.2 |
| BA.2 |
| BA.2.9 |
| BA.2 |
| BA.2 |
| BA.2.9 |
| BA.2 |
| BA.2 |
| BA.2 |
| BA.1.1 |
| BA.2 |
| BA.2 |
| BA.2 |
| BA.2 |
| BA.2 |
| BA.2.9 |
| BA.2 |
| BA.1.1.1 |
| BA.2.12.1 |
| BA.2.10 |
| BA.2 |
| BA.2.12.1 |
| BA.2 |
| BA.1.1 |
| BA.2.10.1 |
| BA.2 |
| BA.2 |
| BA.2 |
| BA.2 |
| BA.2.12.1 |
| BA.2 |

| |
|---|
| BA.2 |
| BA.2.12.1 |
| BA.2.9 |
| BA.2 |
| BA.2.3 |
| BA.2 |
| BA.2 |
| BA.2.12.1 |
| BA.2.12.1 |
| BA.2.12.1 |
| BA.2.12.1 |
| BA.2.12.1 |
| BA.2.12.1 |
| BA.2.12.1 |
| BA.2.12.1 |
| BA.2.12.1 |
| BA.2.12.1 |
| BA.2.12.1 |
| BA.2.12.1 |
| BA.2.12.1 |
| BA.2.12.1 |
| BA.2.12.1 |
| BA.2.12.1 |
| BA.2 |
| BA.2 |
| BA.2 |
| BA.2 |
| BA.2 |
| BA.2 |
| BA.2 |
| BA.2.9 |
| BA.2 |
| BA.2.12.1 |
| BA.2 |
| BA.2 |
| BA.2 |
| BA.2 |
| BA.2 |
| BA.2 |
| BA.1.1 |
| BA.2.9 |
| BA.2 |
| BA.2.12.1 |

| |
|---|
| BA.2.12.1 |
| BA.2.12.1 |
| BA.2.12.1 |
| BA.2.12.1 |
| BA.2.12.1 |
| BA.2.12.1 |
| BA.2.12.1 |
| BA.2.12.1 |
| BA.2.12.1 |
| BA.2.12.1 |
| BA.2.12.1 |
| BA.2.12.1 |
| BA.1 |
| BA.1 |
| BA.1 |
| BA.1.1 |
| BA.1.1 |
| BA.1.1 |
| BA.1.1 |
| BA.1.1 |
| BA.1.1 |
| BA.1.1 |
| BA.1.1 |
| BA.1.1 |
| BA.1.1 |
| BA.1.1 |
| BA.1.1 |
| BA.1.1 |
| BA.1.1 |
| BA.2 |
| BA.2 |
| BA.2 |
| BA.2 |
| BA.2 |
| BA.2 |
| BA.2 |
| BA.2 |
| BA.2 |
| BA.2 |
| BA.2 |
| BA.2 |
| BA.2 |
| BA.2 |

| |
|---|
| BA.2 |
| BA.2 |
| BA.2 |
| BA.2 |
| BA.2 |
| BA.2 |
| BA.2 |
| BA.2 |
| BA.2 |
| BA.2.3 |
| BA.2.3 |
| BA.2.9 |
| BA.2.9 |
